# A Time-to-Event Comparison of Immune and Endocrine Biomarkers and Latent Profiles in Hospitalisation: An Outcome-wide Approach

**DOI:** 10.64898/2026.01.09.26343800

**Authors:** Odessa S. Hamilton, Olesya Ajnakina, Philipp Frank, Shaun Scholes, Andrew Steptoe

## Abstract

Background. Early identification of risk for hospitalisation is crucial to reducing public health burden. Immune and endocrine-related markers are robust indicators of disease in epidemiological studies, but their value has not been consistently established with severe disorders requiring hospitalisation. Patterning of biomarker expression through latent profile analysis (LPA), may improve predictive accuracy for clinical outcomes above individual biomarkers alone. Method. Four biomarkers (C-reactive protein; fibrinogen; leukocytes; insulin growth-factor-1) measured in the English Longitudinal Study of Ageing (ELSA) in 2008 were linked to administrative data on hospitalisations obtained from Hospital Episode Statistics (HES). Hospitalisation for 12 disease classes was monitored from 2008-2018 (*n*=4,940). Analyses were adjusted for genetic predisposition and a wide set of confounders. Findings. There were 9,419 cases of hospitalisation over the 10-year follow-up period. LPA of the four biomarkers indicated a three-profile solution offered greatest parsimony, categorised as *low-risk* [52.43%]; *moderate-risk* [35.89%]; and *high-risk* [11.68%] inflammatory status. Profiles offered greater specificity than individual biomarkers with a risk gradient in hospitalisation for sleep, circulatory, endocrine, and genitourinary disorders, where the magnitude of associations was notably higher in the *high-risk* group. Profiles also identified risk for infection-related hospitalisation not identified by individual biomarkers alone (HR: *high*-versus-*low*-*risk*=1.38; 95%CI=1.08-1.78, *p*=0.011). No associations emerged in hospitalisation for digestive, nervous, or skin disorders. Interpretation. LPA enabled more precise risk-stratification and subgroup-specific analyses, with profiles better characterising health outcomes requiring hospitalisation than individual biomarkers. Funding. Biotechnology and Biological Sciences Research Council (BBSRC); Economic and Social Research Council (ESRC); National Institute on Aging.

## Introduction

An important role of the immune and endocrine systems is to facilitate the body’s return to its normal physiological state following challenge.^1^ Although these systems have often been studied separately, they work together in a complex, coordinated way.^1^ While the exact biological mechanisms behind this process are not fully understood,^2^ it is known to be triggered by common stressors,^3^ tissue damage,^4^ and pathogen insults.^5^ Genetic differences, particularly in genes related to inflammation, also play a role by influencing how immune and endocrine responses unfold and how diseases progress in different individuals.^6^ The systemic inflammatory response, combined with genetic vulnerability, is now recognised as a major factor in many serious mental and physical health conditions, which contribute significantly to global disease and mortality.^7^ It is unsurprising then that immune and endocrine markers are considered promising targets for treatment. However, results from large-scale studies and clinical trials often do not align, creating challenges for translating these findings into therapies.^8^ Here, we seek to improve this translation.

Biomarkers are measurable indicators of biological activity that aid in diagnosing and assessing the severity of inflammation. They offer insight into disease mechanisms, facilitating early diagnosis and tailored treatments.^4^ However, due to the complexity of biological systems, targeting a single biomarker may be insufficient.^2^ Examining biomarker patterns may better reveal multisystem dysfunction and guide new therapeutic approaches, emphasising the need for research that fully captures immune and endocrine disturbances in disease. Observational studies remain vital, offering cost-effective, real-world evidence for diagnosis and prognosis, while identifying potential targets for intervention.

This study aims to improve the translation of observational inflammation research into clinical applications by linking biomarkers with hospital admission data while incorporating genomic data to address genetic susceptibility. The advantages of this approach are twofold. First, the study outcomes do not rely on participants’ reports of their health status, which may not always be reliable particularly among older individuals. Second, the study focuses on more severe health outcomes that require hospitalisation, rather than milder health problems. We also used latent profile analysis (LPA) to identify patterns of inflammation and endocrine activation in a large sample of older adults living in private households, aiming to uncover subgroups with distinct risk profiles. This probabilistic modelling approach enhances relevance to clinical settings by improving parsimony and addressing statistical issues such as multicollinearity, nonlinearity, and unobserved heterogeneity, with a built-in penalty to prevent overfitting.^9^ The model is designed to provide a more nuanced understanding of immune and endocrine contributions to disease while reducing selection bias in biomarkers and participants.

Four genetically influenced immune and endocrine biomarkers (i.e., C-reactive protein [CRP], fibrinogen [Fb], white blood cell counts [WBCC], and insulin-like growth factor-1 [IGF-1]) were modelled individually, then analysed through LPA to estimate a 10-year hospitalisation risk across 12 exploratory ICD-10 disease groups, ranging from psychiatric disorders to respiratory system disorders. These biomarkers represent distinct inflammatory pathways:^10^ CRP and Fb are liver-derived acute-phase proteins; WBCC reflects immune cell lineages from bone marrow; and IGF-1, is a liver-secreted hormone related to ageing, regulated by growth hormone. Each biomarker also been linked to disease.^11^ This exploratory approach assesses whether latent profiles offered more comprehensive information on disease risk than independent biomarkers across all categories.

## METHODS

### Data

Fully anonymised data were drawn from the English Longitudinal Study of Ageing (ELSA), an ongoing multidisciplinary, observational study. Individual-level administration data from English National Health Service (NHS) hospitals, linked to ELSA, were drawn from the Hospital Episode Statistics (HES) database. Full details for both datasets can be found in Supplementary Materials (SM) 1.

### Study Design

Sample derivation is from the nurse visit at wave (W) 4 (2008/baseline) to censoring (2018). A total of 6,523 participants had complete measures and at least one biomarker at baseline. After exclusions of CRP values >20 mg/L (*n*=116 taken to reflect acute rather than chronic inflammation), the sample was 6,407. Of these, 1,467 had missing genetic data at W4, leaving an ELSA sample of 4,940 and 12 distinct analytic samples ranging 3,163-4,276 that can be seen in Supplementary (S) Table 1.

**Table 1.**
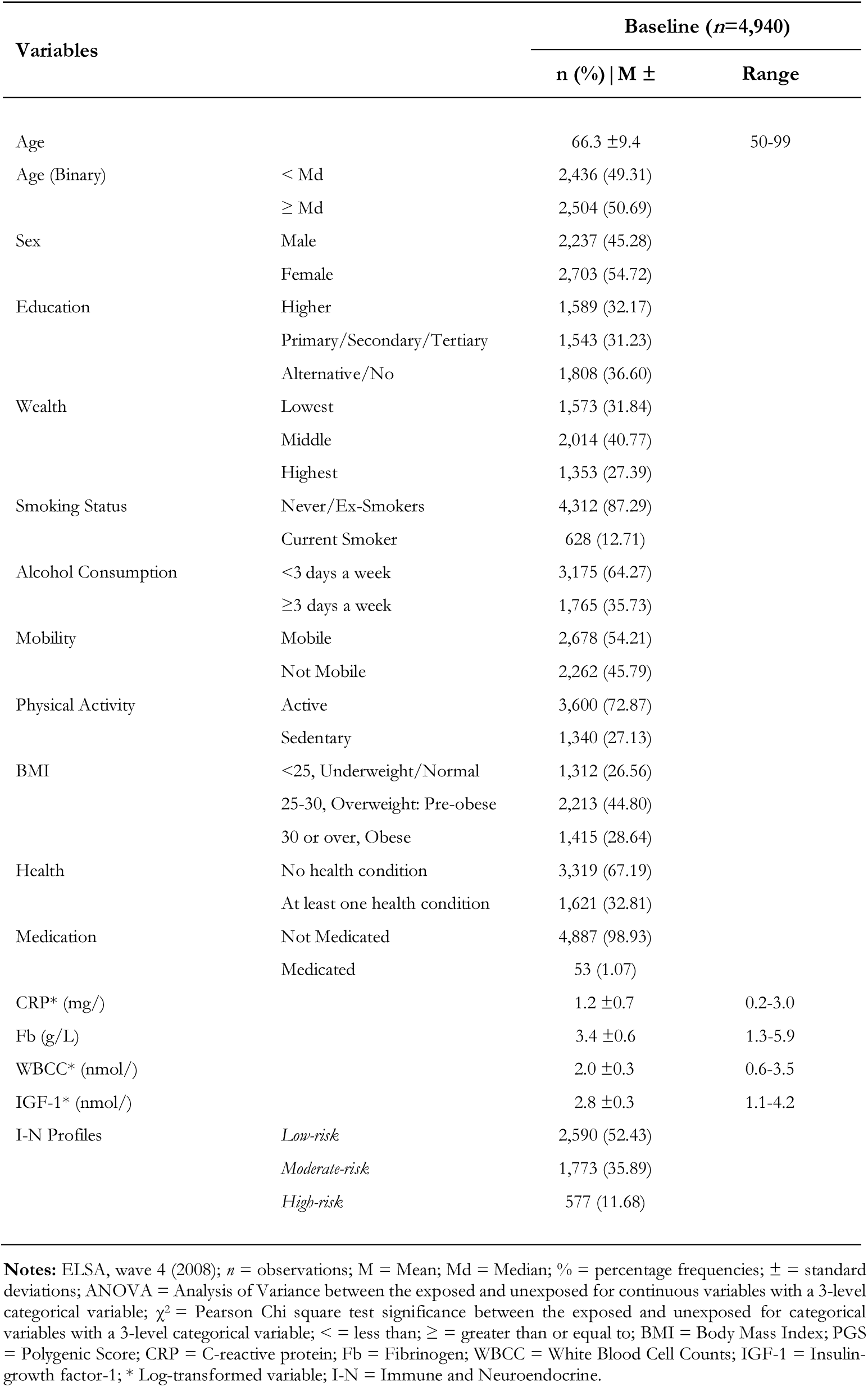
Sample characteristics

### Exposures

Immune and endocrine biomarkers were measured at baseline and included high-sensitivity plasma C-reactive protein (CRP; mg/L), plasma fibrinogen (Fb; g/L), plasma leukocytes/white blood cell counts (WBCC; 109/L), and serum insulin-like growth factor-1 (IGF-1; mmol/L). Selection was based on availability of immune and endocrine-related biomarkers. Fb was normally distributed, but due to an initially skewed distribution, logarithmic transformation was performed on CRP, WBCC, and IGF-1 values. As compared to the other biomarkers, IGF-1 values follow a counter-directional pattern. Exclusion criteria for obtaining bloods included coagulation issues, history of convulsions, haematological disorders, or being on anticoagulant medication. Blood samples were discarded if deemed insufficient or unsuitable (e.g., haemolysed; received at the laboratory >5 days post-collection; full details in SM2).

### Outcomes

Outcomes were based on the date of first hospital admission (i.e., incident cases) from one of 12 ICD-10 clinical chapters (Table S1):- Psychiatric disorders (F00-F50l, F52-F99); Sleep disorders (F51; G4); Infectious and parasitic disorders (A00-B99); Disorders of the blood, blood-forming organs, and related disorders of the immune mechanism (D50-D89); Endocrine, nutritional, and metabolic disorders (E00-E90); Nervous system disorders (G00-G99); Circulatory system disorders (I00-I99); Respiratory system disorders (J00-J99); Digestive system disorders (K00-K93); Skin and subcutaneous tissue disorders (L00-L99); Musculoskeletal system and connective tissue disorders (M00-M99); Genitourinary system disorders (N00-N99).

### Covariates

Confounding variables were identified *a priori* through a directed acyclic graph (DAG; Figure S1; SM2). All cases before baseline were excluded (i.e., hospital admissions prior to the date of ELSA participation at the W4 nurse visit [2008]). Covariates included: *demographic variables*: age (≥50 years); sex (male; female); *socioeconomic variables*: education (higher; primary/secondary/tertiary; or no formal education); wealth (tertials of household wealth, determined by the summation of property [minus mortgage], business assets, possessions, liquid assets; cash, savings, investments, artwork, and jewellery, net of debt, exclusive of pension); *lifestyle variables*: alcohol consumption (low [<3]; high [≥3] days weekly); smoking status (non-smokers/ex-smokers; smokers); *clinical variables*: mobility difficulties (≥1 difficulties [walking 100 yards; sitting 2-hours; rising from chairs after long periods sitting; climbing stairs; stooping, kneeling, crouching; extending arms above shoulders; shifting large objects; lifting/carrying objects >10lb; picking-up a 5p coin]); *genetic variables*: 10 principal components (PCs), polygenic scores (PGS) for inflammatory, endocrine, pain-related, psychiatric, and cardiometabolic traits relevant to the 12 disease categories of interest (SM4).

## Statistical Analyses

Polygenic Scores (PGS). PGS computation is detailed in SM4. To ensure PGSs with different distributions could be appropriately compared and to improve interpretability, Z-scores were used for standardisation.

Imputation. Missing data from attrition and item non-response was imputed using missForest, an iterative, machine learning imputation method, predicated on a Random Forests algorithm (SM5). Missing values were imputed in R (v.4.4.1) on exposures where at least one biomarker was available and on covariates except genetic data.

Latent profile analysis (LPA). An LPA was performed to uncover patterns of immune and endocrine activity.^12^ A stepwise approach was taken to identify the optimal number of latent profiles, informed by the Akaike (AIC), Bayesian (BIC), and adjusted Bayesian information criteria (aBIC), along with the likelihood ratio test, entropy, and posterior probabilities (complete specifications and benchmarks for the LPA are detailed in SM6).

Descriptive Statistics. Baseline characteristics were expressed as means and standard deviations for the continuous variables and counts and proportions for categorical and binary variables.

Survival Analyses. Cox proportional hazards (PH) regression models were used to estimate the Hazard Ratios (HR) and 95% confidence intervals (95%CIs) of associations between immune and endocrine latent profiles at baseline (W4; 2008) and hospitalisation incidence from 12 disease classes (described above) across a 10-year follow-up period (to January 2018, the last date of linkage to HES). Proportional hazards assumptions were met. A monotonic trend, where biomarker values consistently increase or decrease across observations, was also computed by treating each biomarker as a continuous variable rather than a categorical profile. To address multicollinearity, each biomarker was modelled independently. Prevalent cases of each outcome at baseline were excluded, leading to a variation in sample sizes across outcomes. Immune and endocrine profiles and biomarkers were computed per 1000 person-years, to assess incidence rates of hospitalisation across the follow-up period. Survival time was calculated from the date of the W4 nurse visit (2008) to the first hospital admission date or censorship (i.e., study end [2018]), whichever came first. Hospitalisation for each of the 12 disease classes was analysed separately, with three models used for different levels of covariate adjustment: model (M)1 was unadjusted; M2 adjusted for baseline *demographic* and *genetic* variables; M3 was fully adjusted. Association analyses were conducted in Stata 17.1.

Sensitivity Analyses. Three sensitivity analyses were performed. First, exploratory studies do not strictly require multiplicity adjustment,^13^ but multiple testing was addressed for sensitivity by controlling the false discovery rate (FDR) using the Benjamini-Hochberg procedure, which is robust to positive dependence among tests.^14^ The FDR correction was applied separately to profile-level analyses (24 tests; 2 exposures × 12 outcomes) and individual biomarker analyses (48 tests; 4 exposures × 12 outcomes). Benjamini-Hochberg-adjusted *q*-values were computed in R (v.4.4.1), according to:

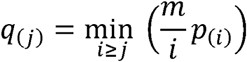

where *p*_(_*_i_*_)_ denotes the ordered *p*-values and *m* the total number of tests within a given analytic domain. Results with *q*<0.05 were considered statistically significant. Second, further self-reported diseases were adjusted for (viz., chronic lung disease; coronary heart disease; abnormal heart rhythm; heart murmur; congestive heart failure; angina; hypertension; diabetes; cancer; Parkinson’s disease; Alzheimer’s disease; dementia; asthma; arthritis; osteoporosis; psychiatric disorder) to ensure that their omission was not biasing results, despite earlier caseness exclusions. Third, some measures are likely mediators but are commonly treated as confounders, so they were modelled incrementally for sensitivity (viz. physical activity; body mass index [BMI; kg/m2]; and medications [anticholinergics; antidepressants; antimanics; antipsychotics; antithrombotics; benzodiazepines {BZDs}; calcium channel blockers {CCBs}; central nervous system {CNS} drugs; diuretics; muscle relaxants; NSAIDs; opioids; renin-angiotensin-aldosterone system {RAAS} inhibitors; steroids]).

## RESULTS

### Cohort characteristics

Summary descriptives of the overall sample (*n*=4,940) are shown in Table 1 (detailed further in Table S2). Observations, events, and time at risk are shown in Table S3. During the 10-year follow-up period, a total of 9,419 hospitalisation events were observed. Participants, male and female (49.3|50.7%), were aged ∼66.3±9.4 years (range [*rg*]: 50-99) at W4. Of these, most were healthy (67.2%), never or ex-smokers (87.3%), and who consumed alcohol less than three days a week. Over half had no reported mobility difficulties (54.2%) and were physically active (72.9%), with few taking medications (1.1%), and half were either underweight to normal weight (26.6%) or obese (28.6%). The highest (wealthiest) capital group was the smallest (27.4%), but there was a fairly equal educational divide (32.1/31.2/36.6%). CRP was positively correlated with Fb (*r*=0.567), and WBCC (*r*=0.301), but negatively correlated with IGF-1 (*r*=-0.164; all at *p*<0.001; detailed further in Table S4).

### LPA of immune and endocrine biomarkers

A three-profile model provided the most parsimonious fit to the underlying data structure; entropy was 0.7, the mean posterior probabilities did not exceed 0.7, each profile had ≥5% of participants, there were limited gains in information criterion value after this model (∼1%; Table S5; Figures S2-5a-g), and the profiles were theoretically meaningful. Profile 1 (*low-risk*), 52.4% of the sample, was characterised by individuals with low CRP, Fb, WBCC, and high IGF-1. Profile 2 (*moderate-risk*), 35.9% of the sample, was characterised by individuals with moderate CRP, Fb, WBCC, and IGF-1 levels. Profile 3 (*high-risk*), 11.7% of the sample, was characterised by individuals with high CRP, Fb, WBCC, and low IGF-1 (Figure S6-7). Of these, CRP had the greatest percentage-point change between profiles (Table S6).

Longitudinal associations between immune and endocrine profiles and hospitalisation Start this section with some descriptive material about the hospitalisation data. How many cases overall? Which conditions were most common, etc, As illustrated in Figure 1, as compared to the *low-risk* profile, the *moderate-risk* biomarker profile was significantly associated with hospitalisation for sleep, circulatory, endocrine, genitourinary, and respiratory disorders. A higher threshold was required for hospitalisation for psychiatric, blood, infectious, and musculoskeletal disorders, where the *high-risk* profile conferred risk. There was no hospitalisation risk for digestive, nervous, or skin disorders (Table S7).

**Figure 1.**
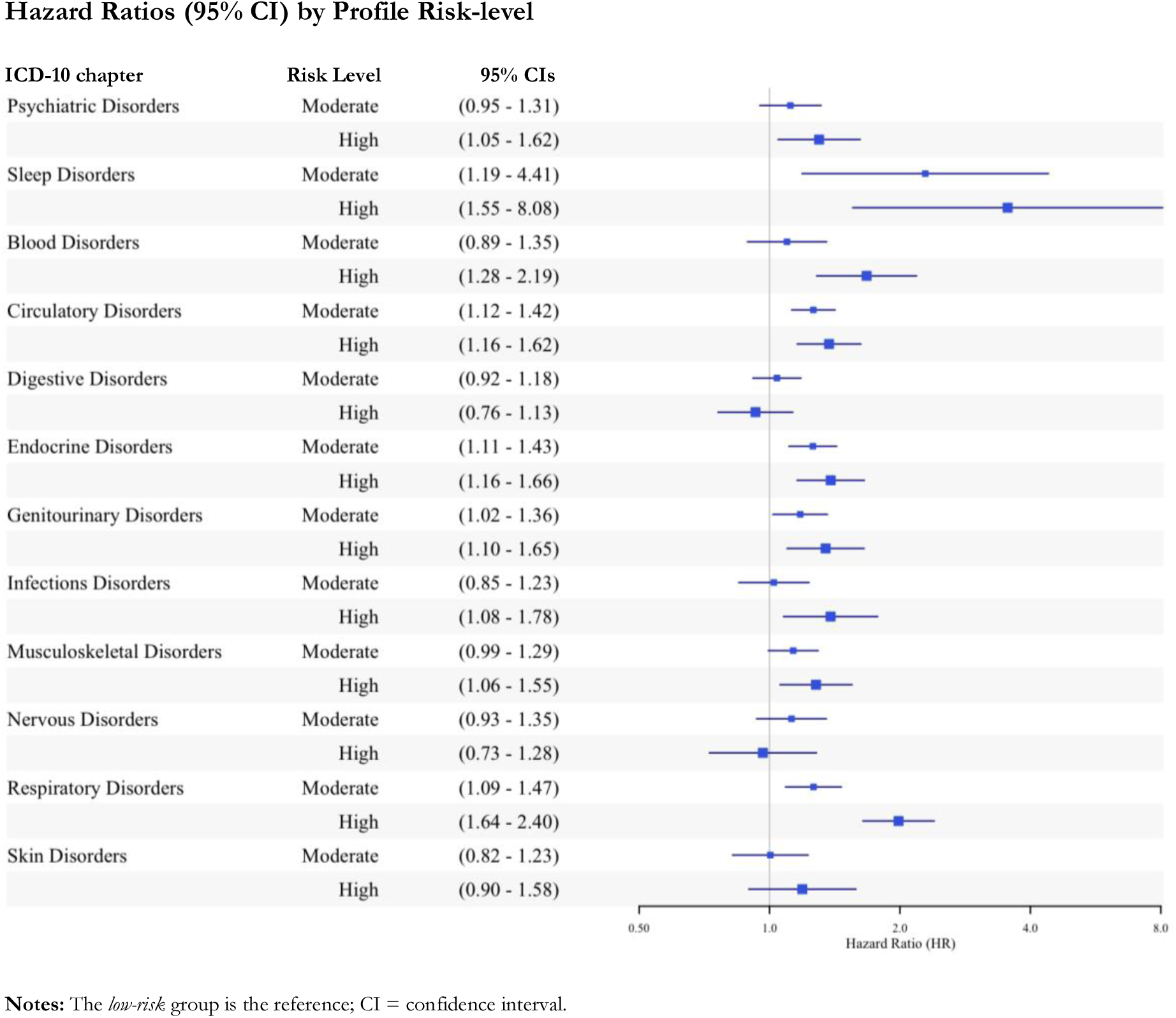
**Associations between latent profiles of immune and endocrine biomarkers and hospitalisation for 12 ICD-10 disease groups**

### Longitudinal associations of individual biomarkers with hospitalisation

Table 2, illustrated in Figure 2, shows CRP, then WBCC had the greatest independent relationships to hospitalisation risk. Fb was associated with hospitalisation for sleep, blood, circulatory, endocrine, genitourinary, and respiratory disorders, but not for disorders of psychiatry, digestion, infection, skin, or musculoskeletal and nervous systems. WBCC was additionally associated with psychiatric disorders and CRP with musculoskeletal disorders too.

**Figure 2.**
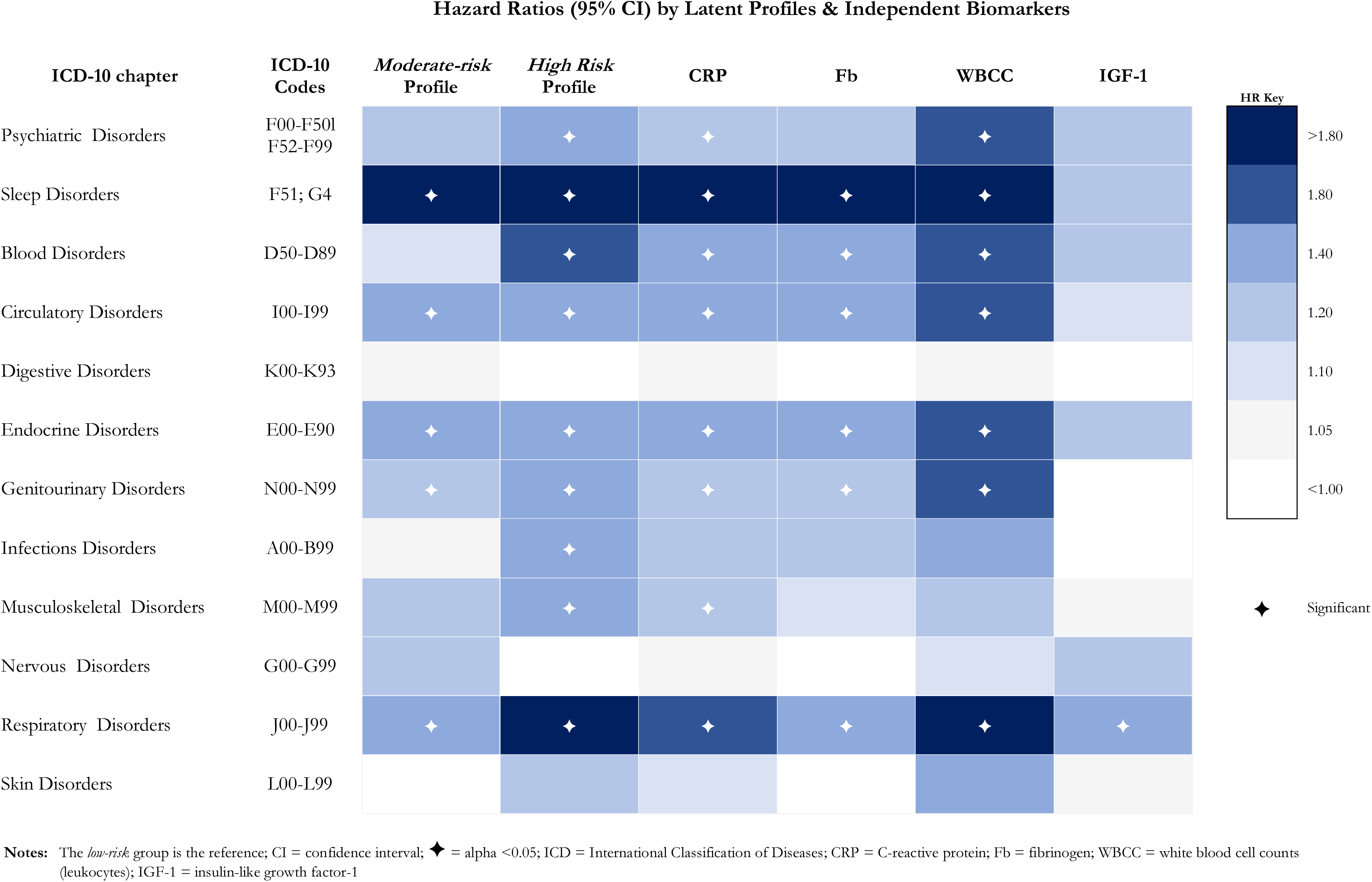
Associations between latent profiles of immune and endocrine biomarkers and independent biomarkers in hospitalisation for 12 ICD-10 disease groups

**Table 2.**
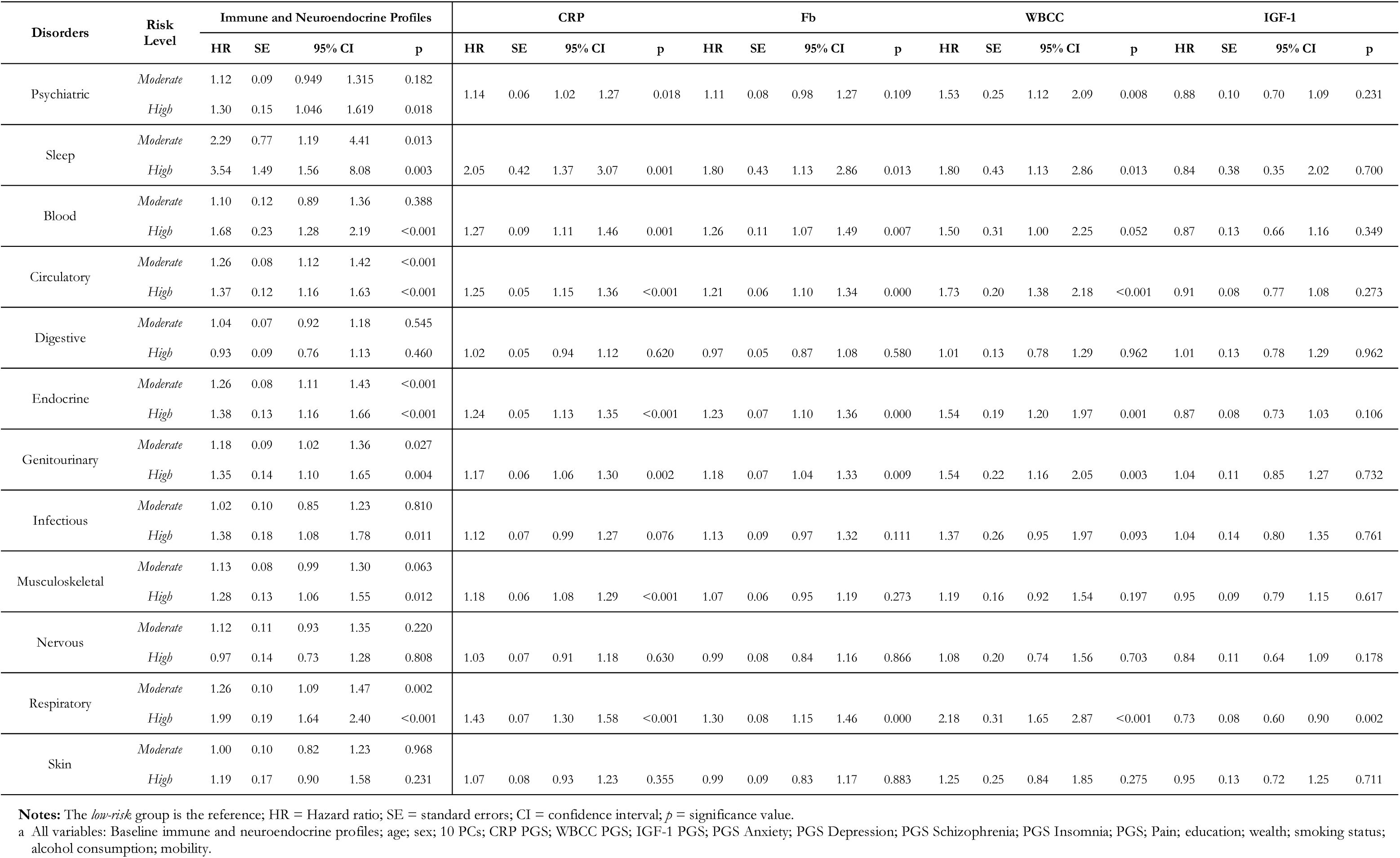
Fully adjusted longitudinal associations between immune-neuroendocrine biomarkers and all-cause hospitalisation

### Profiles and individual biomarkers comparison in hospitalisation

While there were consistent strong associations across all exposures for respiratory-related admissions, for infections, *high-risk* versus low-risk profile membership was associated with a 38% increased risk for hospitalisation (95%CI=1.08-1.78, *p*=0.011), unrealised by independent biomarkers alone. Still, there was an overlap in 95%CI between latent profile and independent biomarker analyses, with the former being larger in most cases. Profiles also revealed a gradient in risk in over half (55.6%) of the categories, with *high-risk* profiles explaining more of the association than the *moderate-risk* profiles, in addition to being statistically different from the *low-risk* profiles, but 95%CI overlapped in all cases except for respiratory-related hospitalisation (Table 3; Figure 2).

### Sensitivity Analyses

FDR adjustment did not attenuate any associations relative to nominal significance (Table S8). Finally, conditioning on additional disease categories did not bias results, and results were materially unchanged with the inclusion of physical activity, BMI, and medication (Tables S9a-bh).

## DISCUSSION

We sought to improve the translation of observational findings, by identifying empirically-validated subgroup differences among the population that stratified individuals into a trifecta of risk through LPA. This novel methodological approach was conducted in a large, prospective cohort study of older adults. We examined the associative difference between latent profiles and independent biomarkers in hospitalisation for 12 diverse disease categories. Three unique biomarker signatures arose from the data. Gradients in risk were exposed in more than half of all cases, and risk for infection-related hospitalisation was identified by the *high-risk* latent profile but not the independent biomarkers. The latter finding highlighting that latent profiles offered marginally more evidence on risk for disease-related hospitalisation than individual biomarkers alone. Results point to the potential underestimation or inflation of risk by independent biomarkers, possibly contributing to evidential inconsistencies and overstated epidemiological claims,^15,16^ but given the overlap of 95%CIs, results are not conclusive. With the exception of IGF-1, there was general concordance between the profiles and independent biomarkers, with a striking lack of efficacy in determining hospitalisation for digestive, nervous, and skin disorders. Results are encouraging and support the idea that biological clusters have a common aetiology,^11^ though levels vary between clusters. This is important because observational studies tend to treat groups as homogenous when natural variation exists, leading to the proposal of ineffective or unnecessary interventions that pose harm to individuals inappropriately intervened on.

A key principle in biomedicine is the interplay between pro-inflammatory and anti-inflammatory signals, coordinated through complementarity of multisystem of pathways involving autocrine, paracrine, and endocrine signalling. ^1,2,7,17^ During inflammation, genes that produce small mediator molecules are activated, often involving proteins not typically present in healthy individuals. In contrast, anti-inflammatory cytokines help regulate this response by controlling acute-phase proteins (e.g., CRP, fibrinogen) and leukocyte activity, either dampening or halting the inflammatory cascade.^4^ Thus, the problem with inflammation is not in its modulation, but its failure to subside and preserve a physiological balance of expression across systems to guarantee health.^1,4,7,18^ However, understanding the molecular mechanisms driving this balance remains a challenge.^2^ Considering this physiological counterbalancing, the dysregulation of a single biomarker is typically not sufficient to radically alter the balance of all others to the point of disease.^1,18,19^ As was confirmed in part here, they are less reflective of this complex process, offering a weaker signal of what is occurring systemically. A dependency on single biomarkers may, thus, offer partial reason for evidential inconsistencies from the same biomarker,^20,21^ why cohort studies infer longitudinal associations not causally confirmed by genetic studies,^22^ why no biomarker universally indicates inflammation-induced disease manifestation,^23^ and why single biomarkers putatively relate to a range of disorders when orchestrated latent contributory effects are more likely.^24^ As biomarkers function within a broader extracellular network, inconsistent results across studies are expected.^4^ The immune and endocrine profiles exemplify the dynamism of synthesis more accurately, and offer an opportunity to leverage a multi-dimensional molecular structure to better understand disease risk. Fortunately, studies examining combinations of biomarkers are increasingly feasible given the burgeoning availability of resources with deep phenotyping (precise, comprehensive data on phenotypic variation including quantitative measures of biomarkers).^19^

Differences in the molecular structure and response timing of cytokines and hormones may explain their varying associations with hospitalisation outcomes observed in the present.^10,18^ These distinctions likely account for why acute-phase proteins (APPs) performed better than IGF-1 in this analysis.^18^ Despite its weaker individual signal, IGF-1 still offers valuable insight into disease risk when combined with other markers.^25^ Endocrine regulation of immune function occurs through autocrine and paracrine interactions among growth factors, cytokines, and immune cells, influencing the development and activity of progenitor and myeloid cells.^25^ Unlike more immediate inflammatory markers like APPs and leukocytes, IGF-1’s role is more indirect and stable over time, which may weaken its statistical signal over the 10-year follow-up period.

Congruent with the present study, Morrisette-Thomas and colleagues (2014)^26^ found that key axes of a novel biological structure played an important role in disease by submitting 19 inflammatory biomarkers to principal component analysis (PCA). However, authors detected two stable axes of variation that explained just 29% of the total variance among the biomarkers highlighting that the system dynamics driving most of the variance could not be characterised by PCA. Whereas LPA applied longitudinally offers a more parsimonious approach that is easier to interpret, overcomes statistical complications, and enhances clinical utility.^9^ Overall, associations identified between the profiles and independent biomarkers for nine of the 12 disease categories requiring hospitalisation offer support of previous reports.^17,23,24^ That said, results challenge the prevailing notion of multimorbidity through pathophysiologically-defined diseases, given the underlying inflammatory morphology of disease, with interrelated genomic and proteomic characteristics.^7^

The lack of association between immune and endocrine markers and hospitalisation for digestive, skin, and especially nervous system disorders was unexpected. Chronic low-grade and neuroinflammation have been linked to cognitive decline, memory impairment, and changes in brain structure, particularly in older adults.^7^ One possible reason for the null findings is that the biomarker panel in the present study was too crude to detect associations.^3^ Alternatively, the link between inflammation and nervous disorders may be weak, with disease development more strongly influenced by environmental factors or central nervous system immune cells not measured in this study. Reverse causation is also possible, where disease drives inflammation rather than the other way around, previous associations may have been confounded by factors creating spurious links.^27,28^

The strengths of this study are in our use of an extensive longitudinal resource, with objective measures linked to real-world evidence in the same individuals. Owing to ICD-10 codes, we allow for international comparisons in clinical assessment. Genetic predisposition is accounted for, which while normative in trials through random assignment, is underutilised in observational research. Our method solved statistical complications not previously feasible in this context.^9^

Notwithstanding, results should be interpreted in light of some limitations. Ideally, a more comprehensive panel of biomarkers that traverses multiple biological systems and includes proximal and distal inflammatory markers of disease, would have been submitted to LPA. Our restricted range limits the scope of our conclusions, but this work is in its infancy and further work is needed to establish best practices for biomarker selection to yield the most predictive profiles.^23^ Although our findings favour greater profile efficacy, the benefit of independent biomarkers cannot be excluded. Despite statistical standardisation, comparisons of absolute biomarker levels may be limited by different assays, population structures, laboratory methods and use of plasma versus serum. Hospitalisation likely captures only severe disorder requiring admission, excluding private care, and both contribute to an underestimated number of individuals at risk, since prodromal symptoms predate admission, typically identified in primary care. Some conditions are also more likely to be diagnosed than others.

## Conclusion

Inflammation plays a significant role in psychiatric and physical disorders, involving a complex intracellular and intercellular network of biomarkers that coordinate to influence disease. While isolated biomarker, siloed system approaches hold promise for diagnosis and prognosis, there remains challenge in translating research to practice. Here, we expand the understanding of risk for disease to the point of hospitalisation. Latent biomarker profiles, controlled for genetic variation, better capture the biologic complexity of this microenvironment. They index overall inflammatory load, assign specificity to population risk that facilitates treatment personalisation, and improve clinical decision-making for better health outcomes.

## Declarations

Funding. ELSA is funded by the National Institute on Aging (R01AG017644), and by UK Government Departments coordinated by the National Institute for Health and Care Research (NIHR). AS is the director of the study. OSH is supported by the ESRC and the Biotechnology and Biological Sciences Research Council (BBSRC), UCL Soc-B Doctoral Studentship (ES/P000347/1). Ethics Approval. The National Research Ethics Service (London Multicentre Research Ethics Committee [MREC/01/2/91] <u>nres.npsa.nhs.uk</u>) granted ethical approval for each of the ELSA waves. All participants provided informed consent, and research was performed in accordance with research and data protection guidelines. Data Availability. The data supporting the conclusions of this article are deposited in the UK Data Archive and are freely available through the UK Data Service (SN 8688 and 5050 <u>discover.ukdataservice.ac.uk</u>). Declaration of Interests. Authors declare that there are no conflicts of interest: no support from any organisation for the submitted work; no financial relationships with any organisations that might have an interest in the submitted work in the previous three years, no other relationships or activities that could appear to have influenced the submitted work. Contributorship. Study funding from AS and OSH. Conception by OSH, AS, and PF. Planning by OSH and AS. Statistical analysis plan by OSH, SS, and OA. Analyses and initial interpretation by OSH, with input from all authors. All authors had access to the data, take responsibility for its integrity, the accuracy of analysis, and its interpretation.

## Data Availability

The data supporting the conclusions of this article are deposited in the UK Data Archive and are freely available through the UK Data Service (SN 8688 and 5050 discover.ukdataservice.ac.uk).

https://discover.ukdataservice.ac.uk

