## Supplementary Figures for "A Time-to-Event Comparison of Immune and Endocrine Biomarkers and Latent Profiles in Hospitalisation: An Outcome-wide Approach"

***Figure S1.*** **Directed acyclic graph of *a priory* confounding**

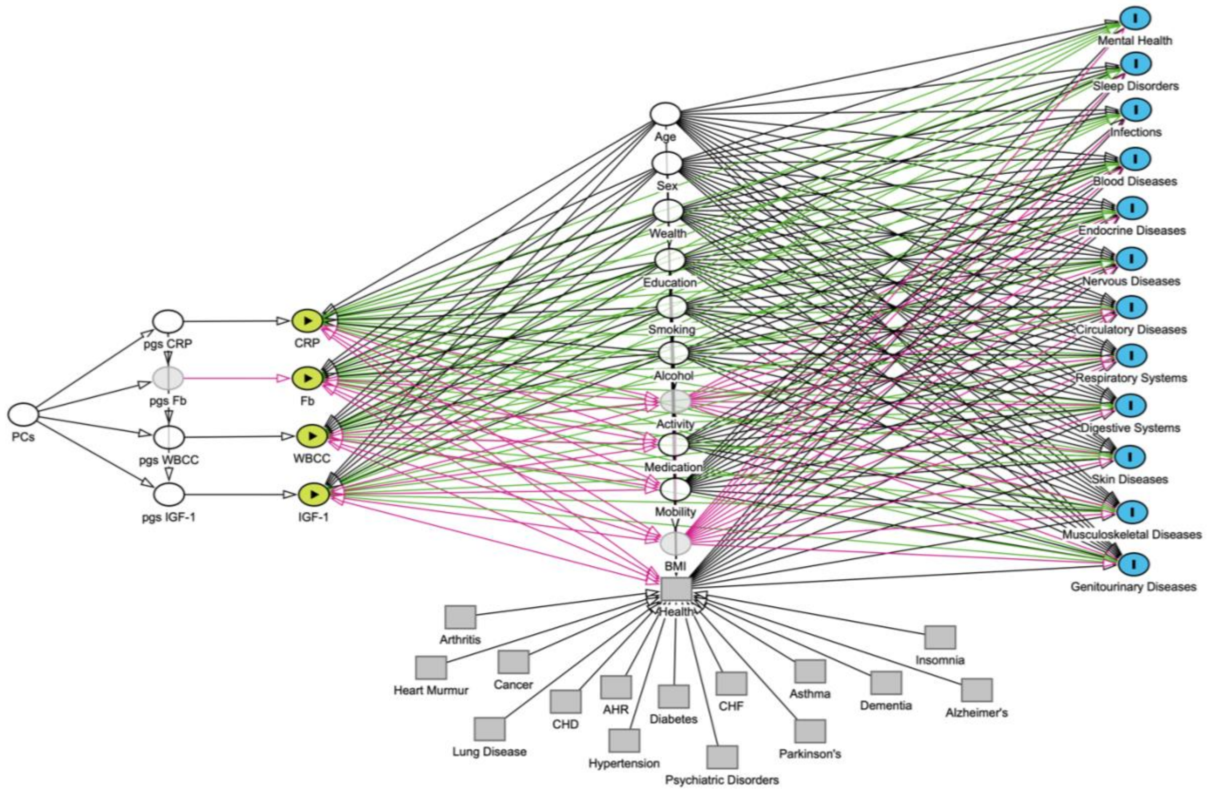


**KEY**

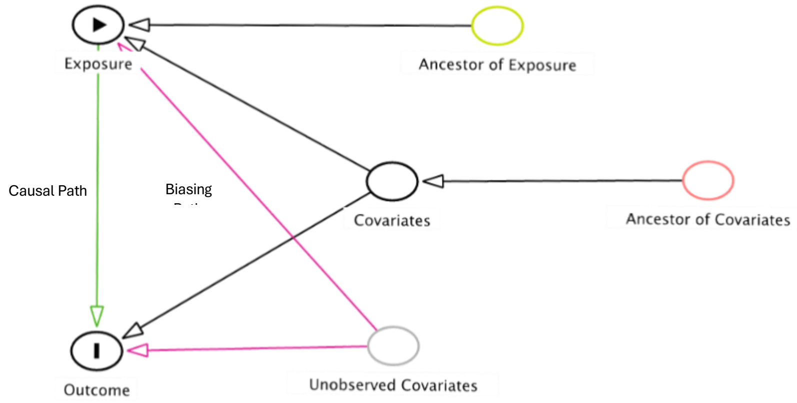

***Figure S2.*** **Akaike Information Criterion (AIC) and Bayesian Information Criterion (BIC) Values of Immune and Neuroendocrine Profiles to Assess Model Fit**

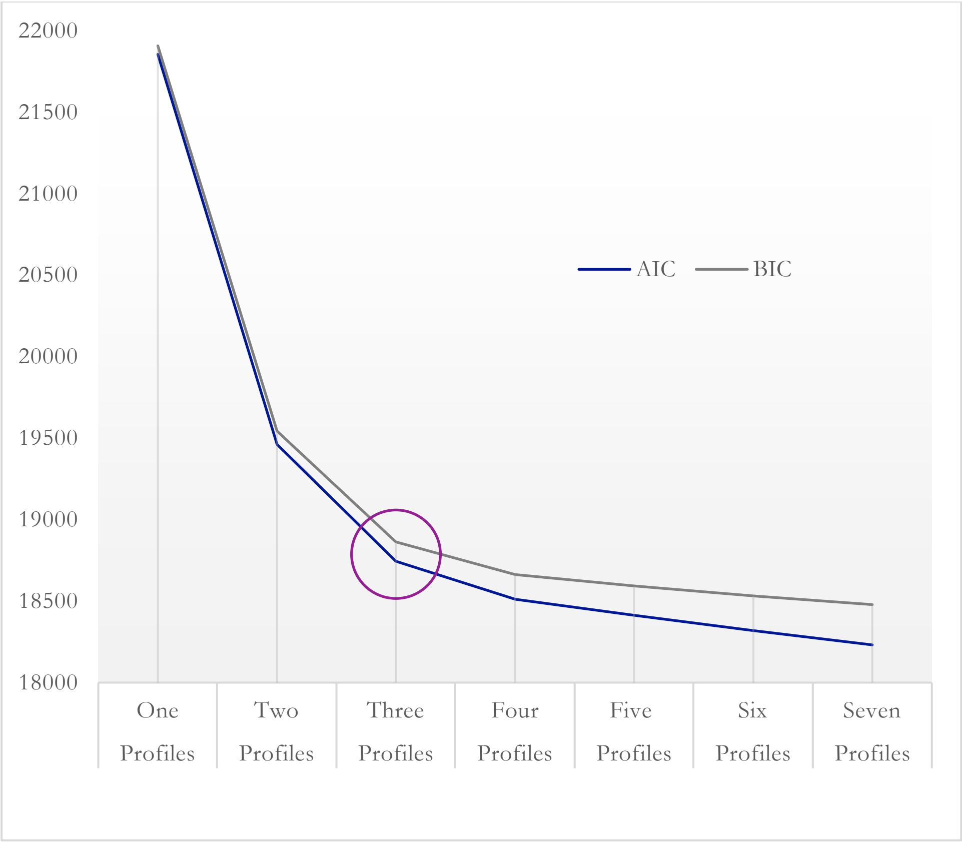

***Figure S3.*** **Entropy and Normalised Entropy Values of Immune and Neuroendocrine Biomarker Profiles to Assess Profile Quality**

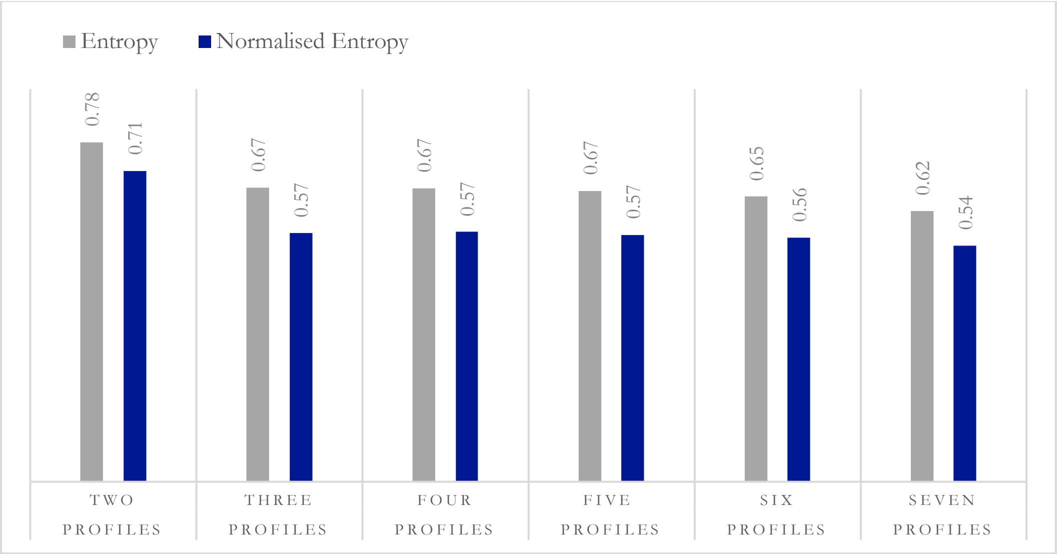

***Figure S4.*** **Mean Posterior Probabilities of Immune and Neuroendocrine Biomarker Profiles to Assess Membership Confidence (≥5%)**

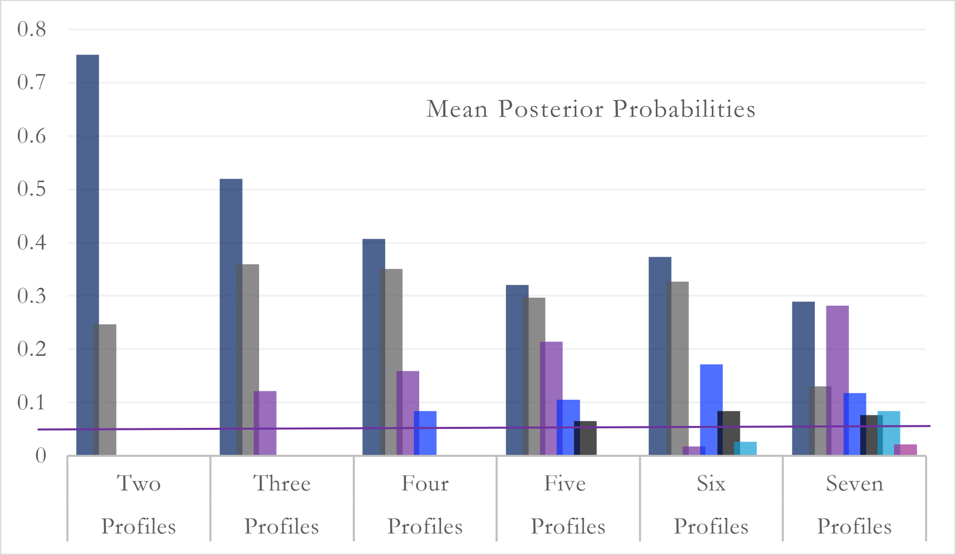

*Figure S5* [a-g]. **Predicted mean of immune and neuroendocrine biomarker levels for a one to seven profile solution (N = 4,940)**

a

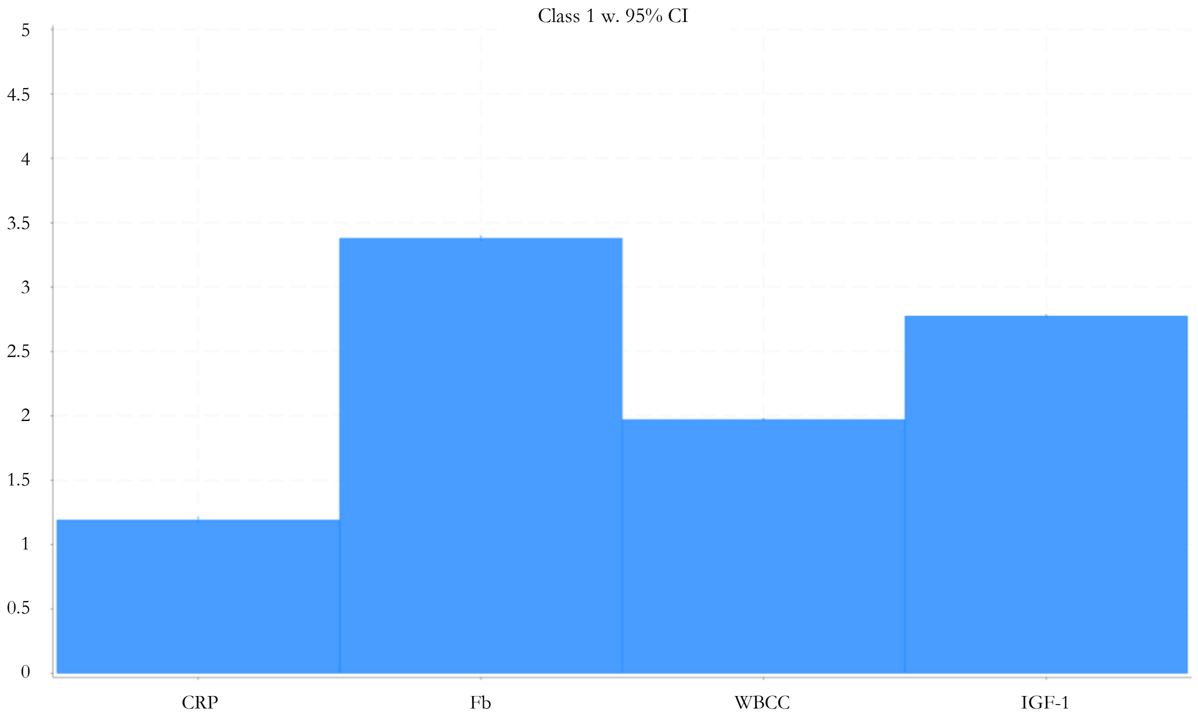


b

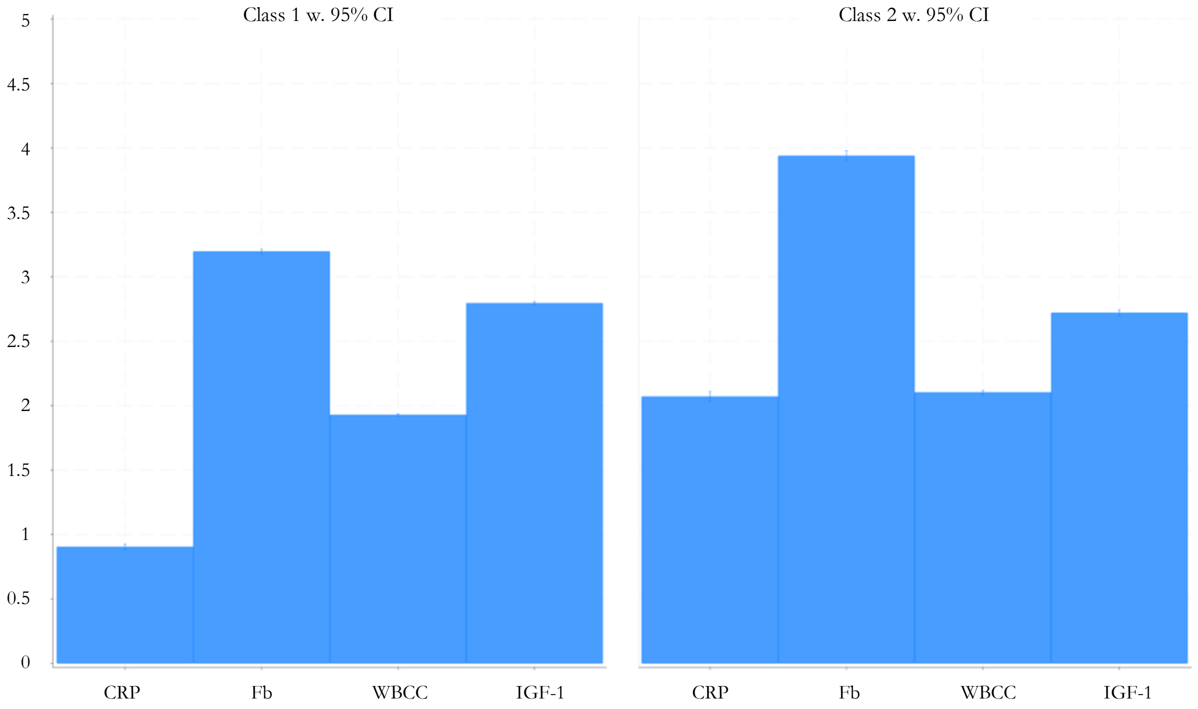

c

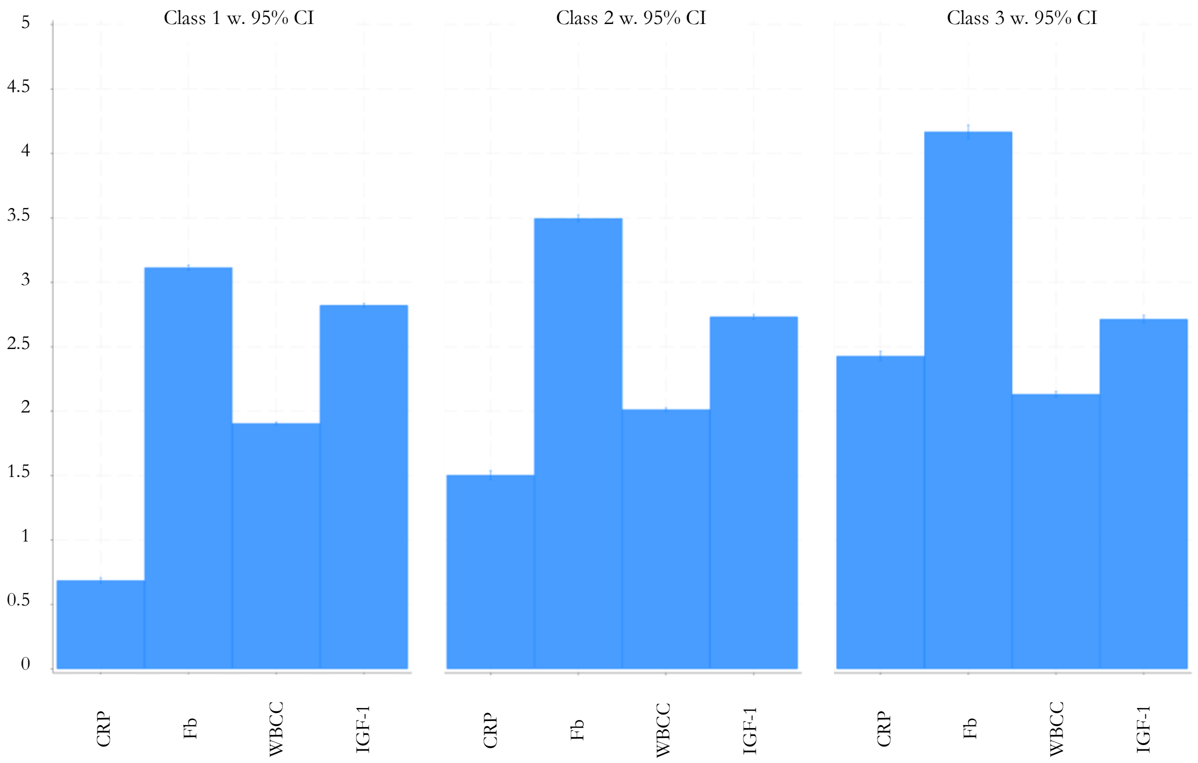
Class 1 w. 95% CI
Class 2 w. 95% CI
Class 3 w. 95% CI
5
4.5
4
3.5
3
2.5
2
1.5
1
0.5
0
CRP
Fb
WBCC
IGF-1

d

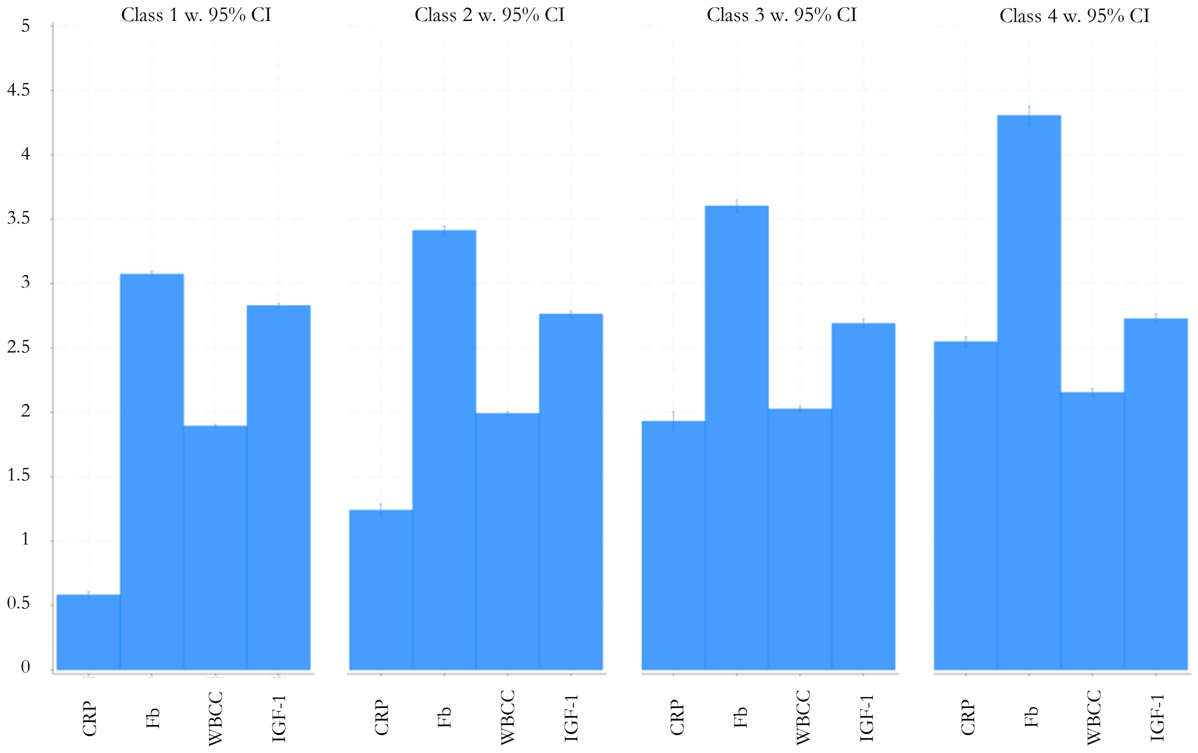
Class 1 w. 95% CI
Class 2 w. 95% CI
Class 3 w. 95% CI
Class 4 w. 95% CI
5
4.5
4
3.5
3
2.5
2
1.5
1
0.5
0
CRP
Fb
WBCC
IGF-1

e

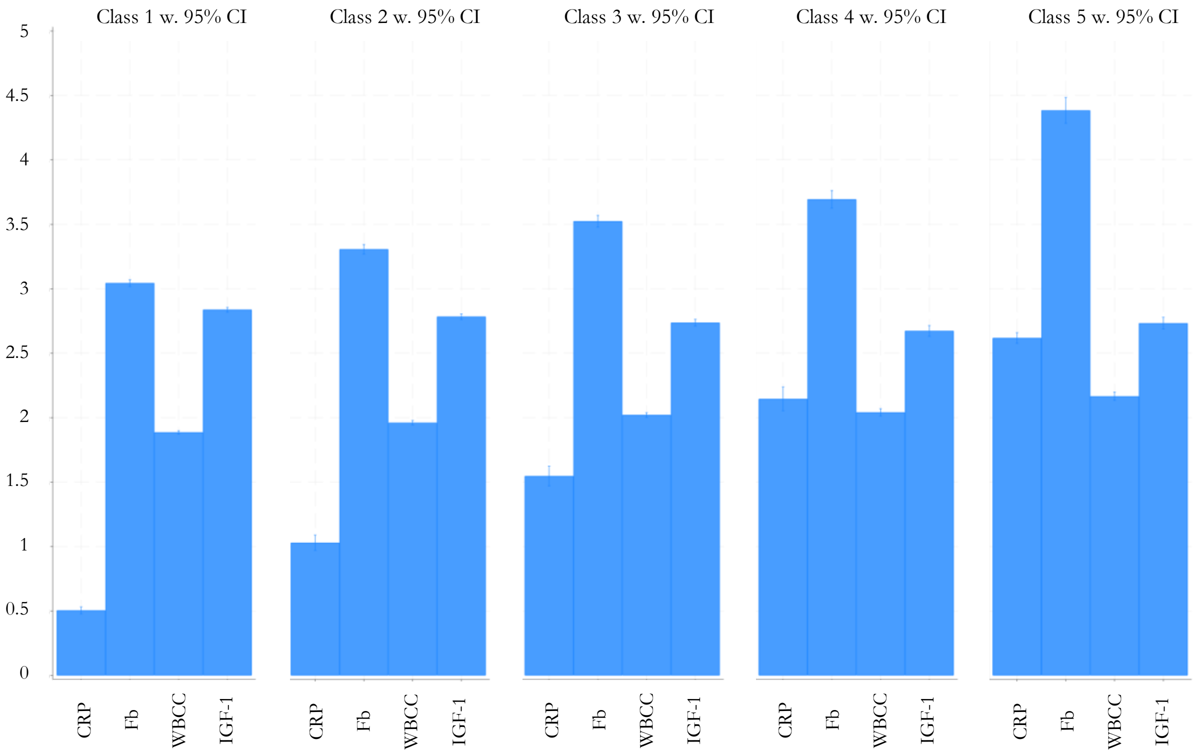

Class 1 w. 95% CI
Class 2 w. 95% CI
Class 3 w. 95% CI
Class 4 w. 95% CI
Class 5 w. 95% CI
0
0.5
1
1.5
2
2.5
3
3.5
4
4.5
5
CRP
Fb
WBCC
IGF-1


f

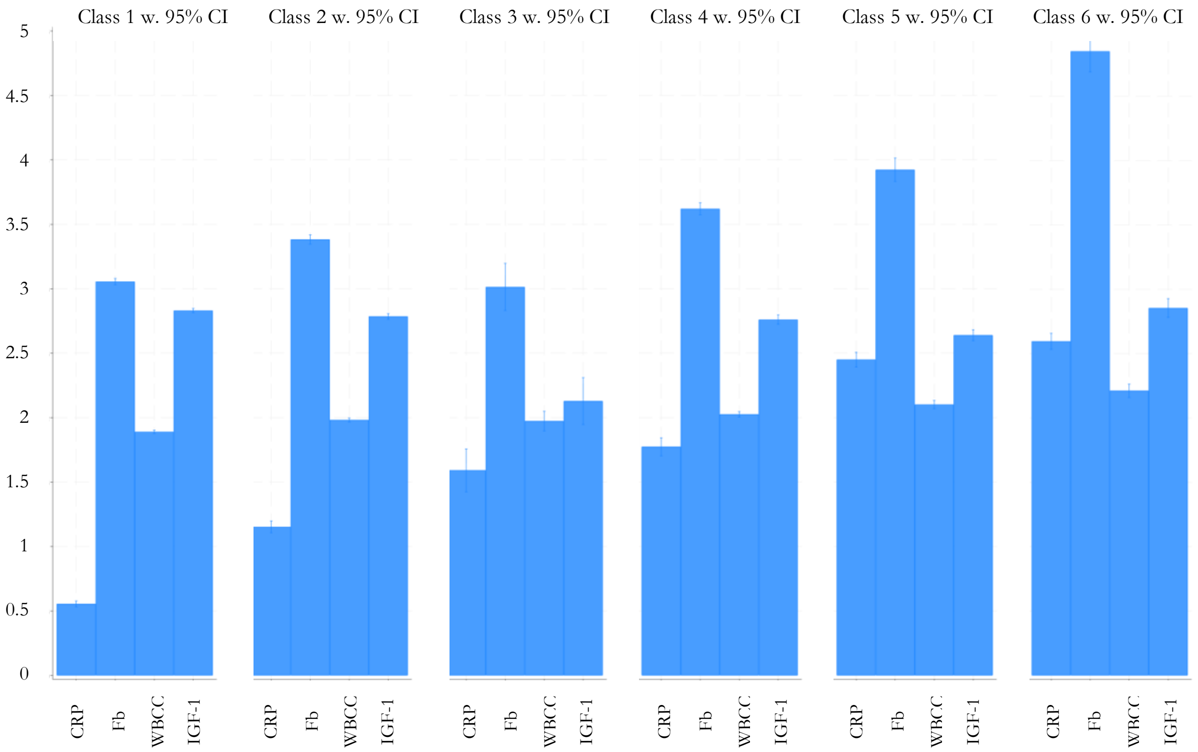

Class 1 w. 95% CI
Class 2 w. 95% CI
Class 3 w. 95% CI
Class 4 w. 95% CI
Class 5 w. 95% CI
Class 6 w. 95% CI
0
0.5
1
1.5
2
2.5
3
3.5
4
4.5
5
CRP
Fb
WBCC
IGF-1

g

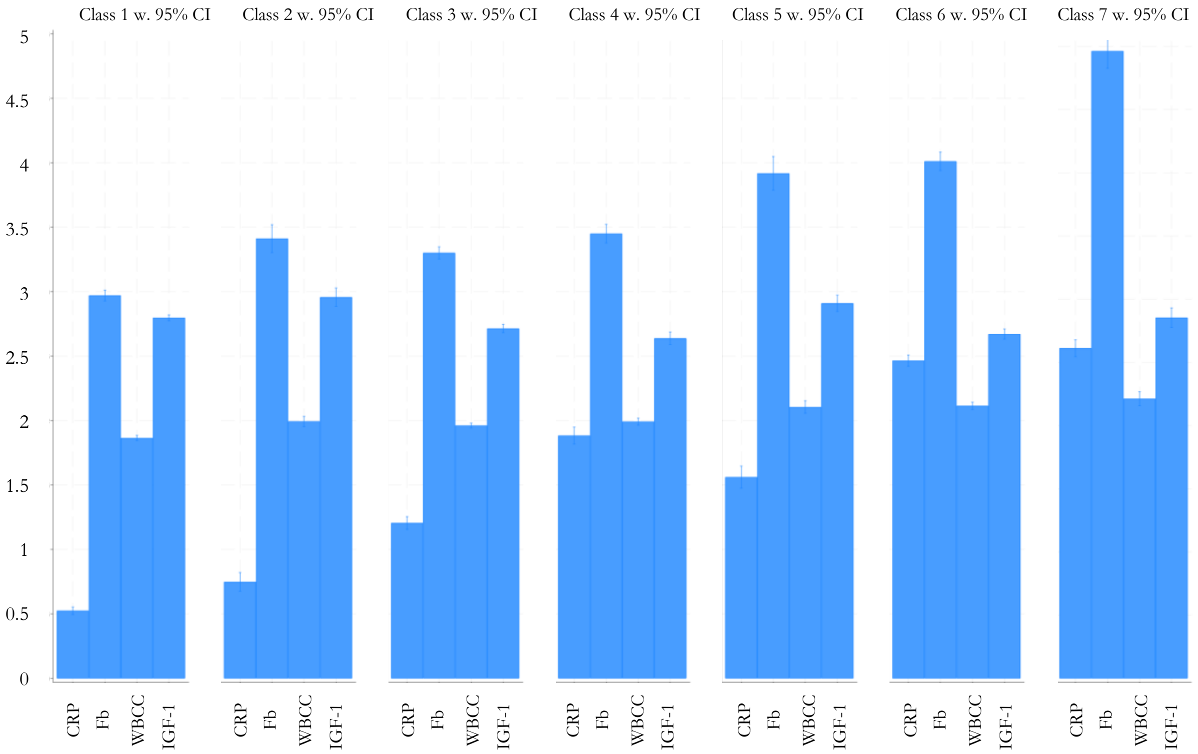

Class 1 w. 95% CI
Class 2 w. 95% CI
Class 3 w. 95% CI
Class 4 w. 95% CI
Class 5 w. 95% CI
Class 6 w. 95% CI
Class 7 w. 95% CI
0
0.5
1
1.5
2
2.5
3
3.5
4
4.5
5
CRP
Fb
WBCC
IGF-1

*Figure S6.* **The percentage of participants belonging to each immune and neuroendocrine biomarker profile with 95% confidence intervals for the wave 4 three-profile solution (N = 4,940)**

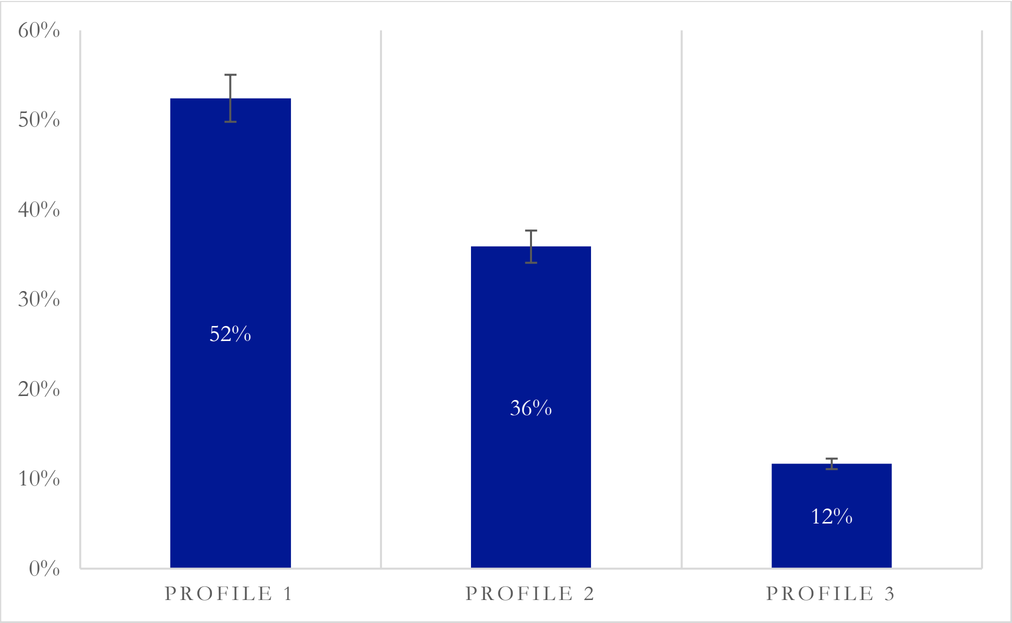


| Profile | N | % |
|---|---|---|
| **1** | 2,590 | 52.43 |
| **2** | 1,773 | 35.89 |
| **3** | 577 | 11.68 |

*Figure S7.* **Predicted Margins of Immune and Neuroendocrine Profiles**

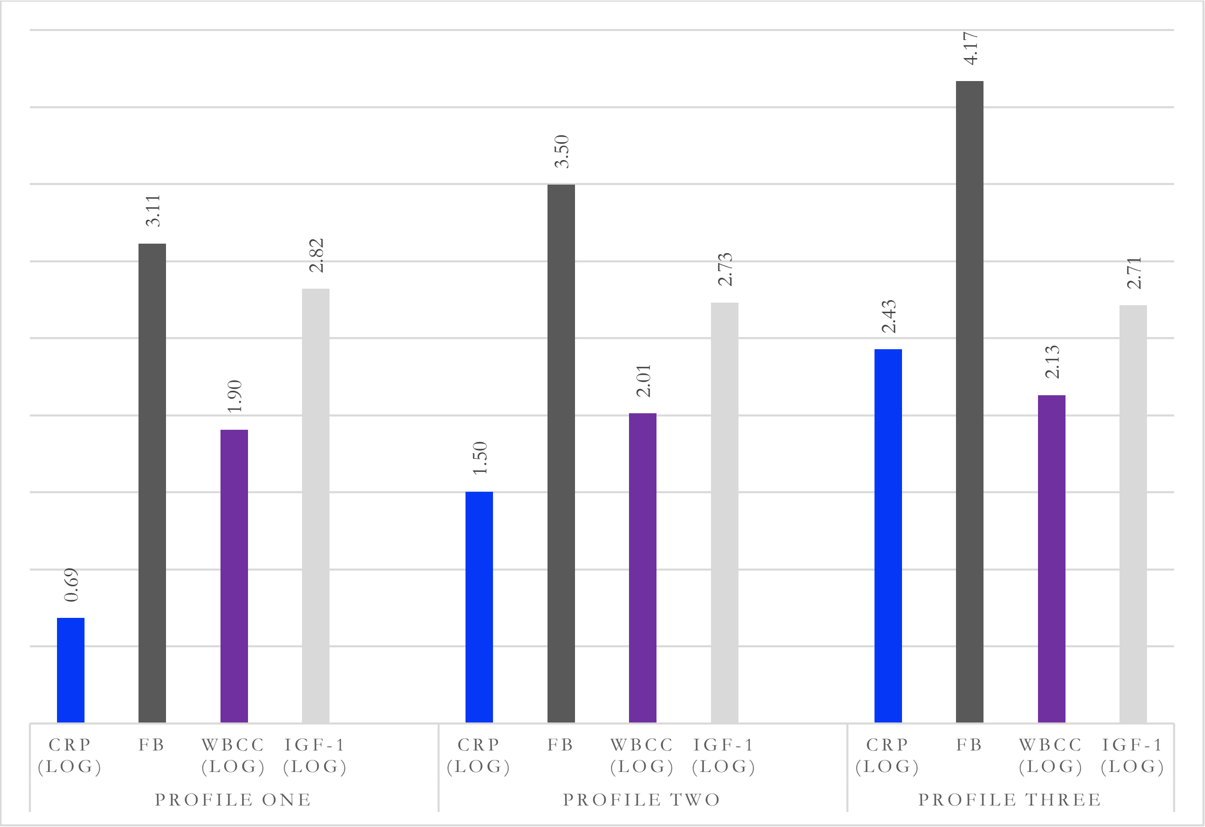
