## Supplementary Materials for "A Time-to-Event Comparison of Immune and Endocrine Biomarkers and Latent Profiles in Hospitalisation: An Outcome-wide Approach"

**Supplementary Materials 1 - Data**

Fully anonymised data were drawn from the English Longitudinal Study of Ageing (ELSA), an ongoing multidisciplinary, observational study, with a household response rate of 70% (Steptoe et al., 2013). Aligned with the National Census, ELSA is representative of the non-institutionalised general population aged ≥50 in England. Data collection is performed in participants' homes, via computer-assisted personal interviews (CAPI) and self-completed questionnaires biennially, then nurse visits quadrennially for biological samples. Not all participants had genetic data at wave 4. All participants provided written consent, and ethical approval was granted by the National Research Ethics Service (London Multicentre Research Ethics Committee). Individual-level administration data, linked to ELSA, were drawn from the Hospital Episode Statistics (HES) database (Boyd et al., 2017). The HES includes demographic, diagnostic, and procedural records for care delivered by all National Health Service (NHS) hospitals in England. Clinicians record all episodes of inpatient, outpatient, and accident and emergency (A&E) care, which amounts to ~14 million records annually. Collected data are submitted to NHS Digital on a monthly basis, then checked by the Secondary Uses Service (SUS). The World Health Organization (WHO) details all diagnoses through the International Statistical Classification of Diseases, Tenth Revision (ICD-10). Records with missing or invalid HES data on patient identifier, date of birth, sex, admission date, episode start date, episode end date, or discharge method were excluded.

**Supplementary Materials 2 - Biomarkers**

**C-reactive Protein.** High-sensitivity plasma CRP (mg/L) was assayed using the N Latex CRP mono Immunoassay on the Behring Nephelometer II analyser (Dade Behring, Milton Keynes, UK). Intra and inter-assay coefficients of variation were <2%. The lower detection limit of the assay was 0.2 mg/L. CRP values >20 mg/L were excluded from analyses ($n$=116), as these were taken to reflect acute inflammatory processes rather than chronic inflammation (Hamilton et al., 2021). CRP was treated as continuous, with higher values indicating greater levels of inflammation.

**Fibrinogen.** Plasma fibrinogen (g/L) was analysed using a modification of the Clauss thrombin clotting method on the Organon Teknika MDA 180 coagulation analyser (Organon Teknika, Durham, USA). Intra and inter-assay coefficients of variation were <7%. The lower detection limit of the assay was 0.5 g/L. Fibrinogen was treated as continuous, with higher values indicating greater levels of inflammation.

**Leukocytes (White Blood Cell Counts [WBCC]).** WBCC was analysed as continuous counts per 109/L; measured on a haematology-automated analyser (Abbott Diagnostics Cell-Dyn 4000 and Sysmex XE), with higher values indicating greater levels of inflammation.

**Insulin-like Growth Factor-1.** Serum IGF-1 (nmol/L) was measured using the DPC Immulite 2000 method, by an electrochemiluminescent immunoassay on IDS ISYS Analyser. Inter and intra-assay coefficients of variation were <14%. IGF-1 was treated as continuous, with lower values indicating greater neuroendocrine activity.

**Supplementary Materials 3 - Directed Acyclic Graph (DAG)**

The DAG serves as validation for the proper parameterisation of the models, to reduce overadjustment bias, and to ensure the adherence of assumptions, *inter alia* homoscedasticity and an absence of interactions (Nilsson et al., 2021; Van Zwieten et al., 2022). It is important to note that while the DAG identifies the presence of bias, it does not explicitly specify the type nor the magnitude of the bias, whether there are competing biases, or whether the observed bias is clinically meaningful (Lipsky & Greenland, 2022).

**Supplementary Materials 4 - Polygenic Scores (PGSs) Derivation**

The genome-wide genotyping, funded by the Economic and Social Research Council (ESRC), was performed at University College London (UCL) Genomics in 2013-2014 using the llumina HumanOmni2.5 BeadChips (HumanOmni2.5-4v1, HumanOmni2.5-8v1.3), which measures ~2.5 million markers that capture the genomic variation down to 2.5% minor allele frequency (MAF).

**Genetics Data Quality Control.** The methods employed for quality control of genomic data in the ELSA study are those outlined by the Health and Retirement Study (HRS; Sonnega et al., 2014) This was done to harmonise the research across the age-related longitudinal studies by adopting a consistent methodology. Single-nucleotide polymorphism (SNPs) were excluded if they were non-autosomal, MAF was <1%, if more than 2% of genotype data were missing and if the Hardy-Weinberg Equilibrium $p<10^{-4}$. Samples were removed based on call rate (<0.99), heterozygosity, relatedness and if the recorded sex phenotype was inconsistent with genetic sex. To identify ancestrally homogenous analytic samples the ELSA genomic samples use a combination of both self-reported ethnicity and analyses of genetic ancestry. To improve genome coverage, we imputed untyped quality-controlled genotypes to the Haplotype Reference Consortium (McCarthy et al., 2016) using the University of Michigan Imputation Server (Das et al., 2016). Post-imputation, we kept variants that were genotyped or imputed at INFO>0.80, in low linkage disequilibrium ($R^2<0.1$) and with Hardy-Weinberg Equilibrium $p$-value$>10^{-5}$. After the sample quality control 7179780 variants were retained for further analyses. To account for potentially biasing ancestry differences in genetic structures, a principal components (PCs) analysis was conducted, retaining the top 10 PCs (Price et al., 2006), which were subsequently used to adjust for possible population stratification in the association analyses (Price et al., 2006; Wang et al., 2009). Genetic ancestry was estimated via comparison of participants' genotypes to global reference populations using principal component analyses (PCA). Because PCA allows examining population structure in a cohort by determining the average genome-wide genetic similarities of individual samples, derived principal components (PCs) can be used to group individuals with shared genetic ancestry, to identify outliers, and as covariates, to reduce false positives due to population stratification. Although up to 98% of the ELSA participants self-described to be of European cultural background, PC highlighted the present of ancestral admixture in *n*=65 (0.9%) individuals (implying these individuals had ancestors from two or more populations. Even though this type of labelling of ancestral populations oversimplifies the complexity of human genetic variation, accounting for systematic differences in allele frequencies is necessary for genetic analyses. Therefore, these participants with ancestral admixture were removed from the analyses. The final sample includes all self-reported European participants that had PC loadings within ± (a standard deviation) from the mean for eigenvectors one. PCs were then re-calculated to further account for population stratification. Therefore, our analytic sample included the full ELSA sample that provided genetic samples and passed quality control. We further utilized the PCs for adjusting for possible population stratification in the association analyses.

**PGS**

All PGS followed the protocols in Ajnakina and Steptoe (2022), outlined here for completeness.

The PGS for C-reactive protein were contracted using the results from UK Biobank (UKB) genome-wide association studies (GWAS), based on 427,367 individuals (Said et al., 2022). The GWAS identified 49,164 genetic loci at a genome-wide significance of $p<5\times10^{-8}$. Linear Mixed Model (LMM) regression using BOLT-LMM version 2.343 was performed on CRP levels in UKB. This model accounts for cryptic relatedness within the sample. An additive genetic model was used for all 8.9 million measured and imputed genetic variants. The model was adjusted for age, sex, UKB array (UKB vs UK BiLEVE to account for the different genotyping chips) and 40 genetic principal components. Serum CRP levels (mg/l) was measured by immunoturbidimetry. CRP levels were transformed using natural log and the resulting range was from -2.53-4.38, excluding individuals with extreme values ±4 from the mean. 1.8% of the sample was removed because they had an autoimmune disorder and were on immunosuppressive drugs.

The PGS for WBCC in ELSA were calculated using summary statistics from GWAS meta-analyses that included data from the UKB and a largescale international collaborative effort, including data for 563,085 European ancestry participants. There were 27,090,932 genetic loci at a genome-wide significance of $p<5\times10^{-8}$, with 5,106 new genetic variants independently associated with 29 blood cell phenotypes covering a range of variation impacting hematopoiesis. The WBCC phenotype (109/L), an aggregate number of white blood cells per unit volume of blood, is one of several quantitative clinical laboratory measures that together reflect hematopoietic progenitor cell production, hemoglobin synthesis, maturation, release from the bone marrow, and clearance of mature or senescent blood cells from the circulation (Vuckovic et al., 2020). Raw phenotypes were regressed on age, $age^2$, sex, principal components, and cohort specific covariates. WBCC related traits were $log^{10}$ transformed before regression modeling. Residuals from the modeling were obtained and then inverse normalised. The cohort level association analyses were conducted using a linear mixed effects model to account for known or cryptic relatedness (e.g., BOLT-LMM, EPACTS https://github.com/statgen/EPACTS and rvtests with the additive genetic model). Linear mixed effects models have been shown to effectively account for both population structure and inter-individual relatedness within the UK Biobank cohort, along with having increased discovery power over simple linear regression with principal components.

The PGS for IGF-1 were calculated using summary statistics from GWAS that included 10,280 men and women in the analyses, comprising 1,712 participants in the Cardiovascular Health Study (CHS), 3,507 in the Framingham Heart Study (FHS), 1,607 participants in the Cooperative Research in the Region of Augsburg (KORA) study and 3,454 in the Study of Health in Pomerania (SHIP; Kaplan et al., 2011). Analyses of SNP associated with IGF-1 concentrations revealed that rs700752 was associated with IGF-I concentrations ($p=4.9\times10^{-9}$), but this was attenuated (meta-analysis $p=0.038$) after adjustment for IGFBP-3 concentrations. Three additional SNPs achieved $p<10^{-6}$ in relation to IGF-I concentrations: rs2153960 on chromosome 6q21, MAF=0.31, $p=5.1\times10^{-7}$; rs1245541 on chromosome 10q22.1, MAF=0.39, $p=5.0\times10^{-7}$; rs7780564 on chromosome 7p21.3, MAF=0.45, $p=3.9\times10^{-7}$.

The PGS for Insomnia were calculated using the GWAS results from the UK Biobank including ~73 million genetic variants in 152,249 individuals. The first ~50,000 samples were genotyped on the UK BiLEVE custom array, and the remaining ~100,000 samples were genotyped on the UK Biobank Axiom array. After standard quality control of the SNPs and samples, which was performed by UK Biobank, the data set comprised 641,018 autosomal SNPs in 113,006 samples of European ancestry for phasing and imputation. Imputation was performed with a reference panel that included the UK10K haplotype panel and the 1000 Genomes Project Phase 3 reference panel. Association tests were performed in SNPTEST using logistic regression with the covariates age, sex (for the full sample), genotyping array, the top five genetically determined PCs and additional PCs out of ten further ones that were associated with the phenotype (tested by logistic regression). The PGS contain 803,361 SNPs that overlapped between the ELSA genetic database and the GWAS meta-analysis; these SNPs were included in the PGS for Insomnia.

The PGS for Major Depressive Disorder (MDD) were created using results from a 2018 GWAS conducted by the MDD working group of the Psychiatric GWAS Consortium (PGC). At the time of preparing the PGS for MDD, the GWAS meta-analysis files are available on the PGC website: http://www.med.unc.edu/pgc/results-and-downloads. PGC conducted a genome-wide association meta-analysis based in 135,458 cases and 344,901 controls, which identified 44 independent and significant loci. MDD cases were required to have a DSM-IV lifetime MDD diagnosis that was collected by a clinician using structured interviews or clinician administered DSM-IV checklists. Most controls were randomly selected and screened for lifetime MDD. GWAS summary statistics contained 8,483,301 SNPs; of these, 1,197,733 SNPs overlapped with the ELSA genetic; these SNPs were included in the PGS for MDD.

The PGS for Anxiety Disorders (Anxiety) were calculated using the GWAS meta-analysis that combined results from nine studies participating in the Anxiety NeuroGenetics STudy (ANGST) Consortium for >18,000 unrelated individuals. Anxiety included generalised anxiety, panic disorder and phobias. The combined case-control meta-analysis included *n*=17,310 and the continuous factor score GWAS included *n*=18,186. All cohorts imputed SNPs to the 1000 Genomes Project references data and approximately 6.5 million SNPs were included in the combined meta-analysis. The regression analyses were adjusted for sex and age at interview, as they were significant predictors of the phenotypes. PCs were estimated for each sample and included on a sample-by-sample basis depending on their correlation with the phenotypes. The authors conducted two types of analyses in each sample based on complementary approaches to modelling the comorbidity and common genetic risk across anxiety disorders: (1) CC comparisons, in which cases were designated as having 'any anxiety disorder' versus supernormal controls, and (2) quantitative FS estimated for every subject in the sample using confirmatory factor analysis. From the ANGST meta-analysis, 1,137,311 SNPs overlapped with the ELSA genetic database and were included in the PGS for Anxiety (factor score) phenotype and 1,068,194 SNPs overlapped with the ELSA genetic database and were included in the PGS for Anxiety (case-control) phenotype.

The PGSs for Schizophrenia (2020) were created using results from a 2020 GWAS conducted by the Schizophrenia Working Group of the Psychiatric Genomics Consortium (PGC). The schizophrenia GWAS combined meta-analysis included 69,369 people with schizophrenia and 236,642 controls and identified 270 independent associations spanning 130 genes. The PGS contain 1,862,381 SNPs that overlapped between the ELSA genetic database and the GWAS meta-analysis; these SNPs were included in the PGS for Schizophrenia.

The PGS for Bipolar Disorder (BD) were created using results from a GWAS sample comprising of 32 cohorts from 14 countries in Europe, North America and Australia, totalling 20,352 cases and 31,358 controls of European descent. Cases were required to meet international consensus criteria (DSM-IV or ICD-10) for a lifetime diagnosis of BD established using structured diagnostic instruments from assessments by trained interviewers, clinician-administered checklists, or medical record review. In most cohorts, controls were screened for the absence of lifetime psychiatric disorders and randomly selected from the population. Variant dosages were imputed using the 1000 Genomes reference panel, retaining association results for 9,372,253 autosomal variants with imputation quality score INFO >0.3 and minor allele frequency (MAF) ≥1% in both cases and controls. Logistic regression of case status on imputed variant dosage was performed using genetic ancestry covariates. The resulting genomic inflation factor ($\lambda$GC) was 1.23, 1.01 when scaled to 1,000 cases and 1,000 controls ($\lambda$1000). The linkage disequilibrium (LD) score regression intercept was 1.021 (s.e.m.=0.010), and the attenuation ratio of 0.053 (s.e.m.=0.027) was nonsignificant, indicating that the observed genomic inflation is indicative of polygenicity rather than stratification or cryptic population structure. The LD score regression SNP heritability estimates for BD were 0.17-0.23 on the liability scale assuming population prevalence of 0.52%.

The PGS for Alzheimer's disease (AD; 2019) were created using results from a large GWAS of clinically diagnosed AD and AD-by-proxy (71,880 cases, 383,378 controls). Participants in this study were obtained from multiple sources, including raw data from case-control samples collected by PGC-ALZ and ADSP (publicly available through dbGaP), summary data from the case-control samples in the IGAP, and raw data from the population-based UKB sample which was used to create a weighted AD-byproxy phenotype. An additional independent case-control sample (deCODE) was used for replication. AD-by-proxy, based on parental diagnoses, showed strong genetic correlation with AD (*rg*=0.81). Cumulatively, the meta-analysis identified 29 risk loci, implicating 215 potential causative genes. Adjustment covariates within each contributing cohort included age, sex, and genetic PCs. The PGS for AD contain 1,712,973 SNPs that overlapped between the ELSA genetic database and the GWAS meta-analysis; these SNPs were included in the PGS for AD.

The PGSs for Migraine were calculated using data from a meta-analysis of 22 GWAS, including data for a total of 59,674 affected subjects and 316,078 controls collected from six tertiary headache clinics and 27 population-based cohorts throughout worldwide collaboration with the International Headache Genetics Consortium (IHGC). This combined data set contained more than 35,000 new migraine cases not included in previously published GWAS. These case samples came from both individuals diagnosed by a doctor and individuals with self-reported migraine as stated on questionnaires. The final combined sample consisted of 59,674 case samples and 316,078 controls in 22 non-overlapping case–control data sets. All subjects were of European ancestry (EUR). Missing genotypes were imputed into each sample using a common 1000 Genomes Project reference panel. Association analyses were carried out within each study using logistic regression on the imputed marker dosages, with adjustments made for sex and other covariates where necessary. The association results were combined in an inverse-variance weighted fixed-effects meta-analysis. Markers were filtered for imputation quality and other metrics, leaving 8,094,889 variants for consideration in our primary analysis. It is important to note that only the genetic markers that reached suggestive $P_{GWAS}<5x10^{-6}$ were made available for the public download. Therefore, PGSs for Migraine in ELSA were calculated based on 7,208 markers, of which 1,188 overlapped with the ELSA data; these SNPs were included in the PGS for Migraine.

The PGS for coronary artery disease (CAD) were created using results from a 2011 study conducted by the Coronary ARtery DIsease Genome wide Replication and Meta-analysis (CARDIoGRAM) consortium. The GWAS meta-analysis consisted of 14 studies with a total of 22,233 individuals with CAD (cases) and 64,762 without CAD (controls) of European descent imputed to the HapMap3 CEU panel. Replication was performed in a sample of 56,682 individuals (approximately half cases and half controls). This analysis identified 13 loci newly associated with CAD at $P_{GWAS}<5\times10^{-8}$, which had risk allele frequencies from 0.13-0.91 and were associated with a 6-17% increase in the risk of CAD per allele. The results of these analyses also confirmed the association of 10 of 12 previously reported CAD loci. Study-specific GWAS adjusted for age of onset (cases) or age of recruitment (controls), gender, and genetic PCs. The PGS contain 783,413 SNPs that overlapped between the ELSA genetic database and the GWAS meta-analysis; these SNPs were included in the PGS for CAD.

The PGSs for myocardial infarction (MI) were created using 2015 results from a subgroup analysis of coronary artery disease (CAD) conducted by the Coronary ARtery DIsease Genome wide Replication and Meta-analysis (CARDIoGRAM) consortium. The GWAS is a meta-analysis of 48 studies of mainly European, South Asian, and East Asian, descent imputed using the 1000 Genomes phase 1 v3 training set with 38 million variants. The study interrogated 9.4 million variants and involved 60,801 CAD cases and 123,504 controls. Case status was defined by an inclusive CAD diagnosis (e.g., myocardial infarction, acute coronary syndrome, chronic stable angina or coronary stenosis of >50%). 37 previous loci and 10 new loci achieved genome-wide significance in these analyses. MI subgroup analysis was performed in cases with a reported history of MI (~70% of the total number of cases). No additional loci reached genome-wide significance in the MI analysis. The European ancestry PGSs contain 1,299,282 SNPs that overlapped with the ELSA genetic data and the GWAS meta-analysis; these SNPs were included in the PGS for MI.

The PGSs for Type II Diabetes (T2D, 2018) were created using GWAS meta-analysis that combined the DIAGRAMv3 (stage 1) GWAS meta-analysis with a stage 2 meta-analysis comprising 22,669 cases and 58,190 controls genotyped with Metabochip, including 1,178 cases and 2,472 controls of Pakistani descent (Pakistan Risk Of Myocardial Infarction Study [PROMIS]). Combining stage 1 and stage 2 meta-analyses included 34,840 cases and 114,981 controls overwhelmingly of European descent leading to identification to eight new T2D susceptibility loci at genome-wide significance ($P<5\times10^{-8}$); these SNPs were included in the PGS for T2D.

The PGSs for Rheumatoid Arthritis (RA) were created using results from a 2014 GWAS that was performed in a total of >100,000 subjects of European and Asian ancestries (29,880 RA cases and 73,758 controls), by evaluating 10 million SNPs. From these analyses, 42 novel RA risk loci at a genome-wide level of significance were discovered, bringing the total to 101. After applying quality control criteria, whole-genome genotype imputation was performed using 1000 Genomes Project Phase I (α) European (*n*=381) and Asian (*n*=286) data as references. Associations of SNPs with RA were evaluated by logistic regression models assuming additive effects of the allele dosages including top 5/10 PCs as covariates (where available) using mach2dat v.1.0.16. To calculate the PGS for RA, the negative ORs value from the GWAS summary statistics (the OR <1), the OR measures were not converted to positive values and the reference allele were flipped to represent phenotype increasing PGS. A total of 8,747,962 SNPs were included in the meta-analysis summary statistics for RA. Of these, 1,100,616 SNPs overlapped with the ELSA genetic database; these SNPs were included in the PGS for RA.

The PGSs for chronic pain (Pain) were calculated using the summary statistics from a large-scale GWAS of Multisite Chronic Pain (MCP) in 387,649 UK Biobank participants. To define MCP phenotype, UK Biobank participants were asked via a touchscreen questionnaire about "*pain types experienced in the last month*", with possible answers: '*None of the above*'; '*Prefer not to answer*'; pain at seven different body sites (head, face, neck/shoulder, back, stomach/abdomen, hip, knee); or '*all over the body*'. Where patients reported recent pain at one or more body sites, or all over the body, they were additionally asked whether this pain had lasted for 3 months or longer. Those who chose '*all over the body*' could not also select from the seven individual body sites. MCP was defined as the sum of body sites at which chronic pain (at least 3 months duration) was recorded: 0-7 sites. Those who answered that they had chronic pain '*all over the body*' were excluded from the GWAS. A total of 1,351,316 SNPs overlapped with the ELSA genetic database with the GWAS summary statistics; these SNPs were included in the PGS for Pain.

### Supplementary Materials 5 - Multiple Imputation

Owing to a better powered sample and a greater possibility of bias from case-wise deletion (Sterne et al., 2009), analyses were conducted using imputed datasets. Imputation was conducted in R v.4.2.0: RStudio v.2022.02.2. Random Forests is non-parametric, it works with high-dimensional data, and it evaluates entropy and information gain, so it is robust to noisy data and multicollinearity. It has outperformed prominent imputation methods, such as multivariate imputation by chained equations (MICE) and *k*-nearest neighbours (KNN) in all metrics (Stekhoven & Bühlmann, 2012). In ELSA, socioeconomic and health-related variables are the main drivers of attrition (Steptoe et al., 2013) but these variables were included in the imputation models, so the assumption that missingness was at random (MAR) was likely to be met. Using ELSA data, results from complete case analyses have been consistent with analyses performed in imputed data (Hamilton & Steptoe, 2022; Hamilton et al., 2023). The imputation yielded minimal variable error (continuous: normalized root mean squared error=0.02%; categorical: proportion of falsely classified=0.07%).

**Supplementary Materials 6 - Latent Profile Analysis (LPA)**

Outcomes of immune and endocrine biomarkers were entered into the LPA, which included high-sensitivity plasma C-reactive protein (CRP; mg/L), plasma fibrinogen (Fb; g/L), leukocytes/white blood cell counts (WBCC; 109/L), and serum insulin-like growth factor-1 (IGF-1; mmol/L). A stepwise approach was taken to identify the optimal number of latent profiles; starting with a single-profile model, additional profiles were added to improve model fit. The number of latent profiles was determined on the basis of the Akaike information criterion (AIC; Akaike, 1998), Bayesian information criterion (BIC; Schwarz, 1978), and adjusted Bayesian information criterion (aBIC; Bozdogan, 1987), with the best model having the lowest AIC, BIC, and aBIC values. The information criterion and the likelihood ratio tests represent the goodness of fit for different models. The entropy statistic provides the quality of the classification model, and the average posterior probabilities for each latent profile indicates profile membership classification errors (Celeux & Soromenho, 1996). The closer to 1 these indicators were, the better the classification quality (Morin et al., 2016), although a common cut-off point for posterior probabilities is ≥0.70 (Nagin, 2009). An ≥0.80 entropy indicates clear profile separation (Kamata et al., 2018). Each profile must contain >5% of participants and the profiles must be of good theoretical interpretability (Herle et al., 2020). Each individual within the sample was then assigned to a cluster for which they had the largest posterior probability (i.e., the profile that they most likely belonged to).

**Supplementary Materials 7 - Association Analyses**

The Cox proportional hazards model is $\lambda(t|X) = \lambda 0(t)\exp(\beta_1 X_1 + \beta_2 X_2 + \ldots + \beta_p X_p)$. Where $\lambda(t|X)$ is the hazard function at time $t_t$ given the values of the covariates $X_X$; $\lambda 0(t)\lambda 0(t)$ is the baseline hazard function; and $\beta_p,\beta_p$ are the regression coefficients corresponding to the covariates $\beta_x,\beta_x$.

**Supplementary Materials 8 - Sensitivity Analyses**

All assumptions, including proportionate hazards, were met. For stepwise model adjustments, changes in HRs due to the inclusion of covariates were calculated using the equation $\exp(\beta_{j,\mathrm{new}} - \beta_j)$, which represents the multiplicative change in the HRs for a given covariate $X_j$ after adjusting for the effects of other covariates. Where $\beta_j$ is the original coefficient associated with the covariate $X_j$ in the model before the inclusion of additional covariates, and $\beta_{j,\mathrm{new}}$ is the new coefficient associated with the covariate $X_j$ after the inclusion of additional covariates, reflecting the adjusted effect of $X_j$ in the presence of other covariates.
