## Supplementary Tables for "A Time-to-Event Comparison of Immune and Endocrine Biomarkers and Latent Profiles in Hospitalisation: An Outcome-wide Approach"

*Table S1.* **Diagnostic chapters, codes, and descriptions for hospitalisation episodes**

| Chapter | Code | Description |
|---|---|---|
| **Chapter V** Psychiatric (mental and behavioural) disorders | F00-F09 | Organic, including symptomatic, mental disorders |
| | F10-F19 | Mental and behavioural disorders due to psychoactive substance use |
| | F20-F29 | Schizophrenia, schizotypal and delusional disorders |
| | F30-F39 | Mood [affective] disorders |
| | F40-F48 | Neurotic, stress-related and somatoform disorders |
| | F50, F52-F59 | Behavioural syndromes associated with physiological disturbances and physical factors |
| | F60-F69 | Disorders of adult personality and behaviour |
| | F70-F79 | Mental retardation |
| | F80-F89 | Disorders of psychological development |
| | F90-F98 | Behavioural and emotional disorders with onset usually occurring in childhood and adolescence |
| | F99-F99 | Unspecified mental disorder |
| **Chapter V (F51) and Chapter VI (G47)** Sleep disorders | F51 | Non-organic sleep disorders |
| | F51.0 | Nonorganic insomnia |
| | F51.1 | Nonorganic hypersomnia |
| | F51.2 | Nonorganic disorder of the sleep-wake schedule |
| | F51.3 | Sleepwalking [somnambulism] |
| | F51.4 | Sleep terrors [night terrors] |
| | F51.5 | Nightmares |
| | F51.8 | Other nonorganic sleep disorders |
| | F51.9 | Nonorganic sleep disorder, unspecified |
| | G47 | Sleep disorders |
| | G47.0 | Disorders of initiating and maintaining sleep [insomnias] |
| | G47.1 | Disorders of excessive somnolence [hypersomnias] |
| | G47.2 | Disorders of the sleep-wake schedule |
| | G47.3 | Sleep apnoea |
| | G47.4 | Narcolepsy and cataplexy |
| | G47.8 | Other sleep disorders |
| | G47.9 | Sleep disorder, unspecified |
| **Chapter I** Infectious and parasitic disorders | A00-A09 | Intestinal infectious diseases |
| | A15-A19 | Tuberculosis |
| | A20-A28 | Certain zoonotic bacterial diseases |
| | A30-A49 | Other bacterial diseases |
| | A50-A64 | Infections with a predominantly sexual mode of transmission |
| | A65-A69 | Other spirochaetal diseases |
| | A70-A74 | Other diseases caused by chlamydiae |
| | A75-A79 | Rickettsioses |
| | A80-A89 | Viral infections of the central nervous system |
| | A92-A99 | Arthropod-borne viral fevers and viral haemorrhagic fevers |
| | B00-B09 | Viral infections characterized by skin and mucous membrane lesions |
| | B15-B19 | Viral hepatitis |
| | B20-B24 | Human immunodeficiency virus [HIV] disease |
| | B25-B34 | Other viral diseases |
| | B35-B49 | Mycoses |
| | B50-B64 | Protozoal diseases |

| | | |
|---|---|---|
| | B65-B83 | Helminthiases |
| | B85-B89 | Pediculosis, acariasis and other infestations |
| | B90-B94 | Sequelae of infectious and parasitic diseases |
| | B95-B98 | Bacterial, viral and other infectious agents |
| | B99-B99 | Other infectious diseases |
| **Chapter III** Disorders of the blood and blood-forming organs and certain disorders involving the immune mechanism | D50-D53 | Nutritional anaemias |
| | D55-D59 | Haemolytic anaemias |
| | D60-D64 | Aplastic and other anaemias |
| | D65-D69 | Coagulation defects, purpura and other haemorrhagic conditions |
| | D70-D77 | Other diseases of blood and blood-forming organs |
| | D80-D89 | Certain disorders involving the immune mechanism |
| **Chapter IV** Endocrine, nutritional and metabolic disorders | E00-E07 | Disorders of thyroid gland |
| | E10-E14 | Diabetes mellitus |
| | E15-E16 | Other disorders of glucose regulation and pancreatic internal secretion |
| | E20-E35 | Disorders of other endocrine glands |
| | E40-E46 | Malnutrition |
| | E50-E64 | Other nutritional deficiencies |
| | E65-E68 | Obesity and other hyperalimentation |
| | E70-E90 | Metabolic disorders |
| **Chapter VI** Disorders of the nervous system | G00-G09 | Inflammatory diseases of the central nervous system |
| | G10-G14 | Systemic atrophies primarily affecting the central nervous system |
| | G20-G26 | Extrapyramidal and movement disorders |
| | G30-G32 | Other degenerative diseases of the nervous system |
| | G35-G37 | Demyelinating diseases of the central nervous system |
| | G40-G46 | Episodic and paroxysmal disorders |
| | G50-G59 | Nerve, nerve root and plexus disorders |
| | G60-G64 | Polyneuropathies and other disorders of the peripheral nervous system |
| | G70-G73 | Diseases of myoneural junction and muscle |
| | G80-G83 | Cerebral palsy and other paralytic syndromes |
| | G90-G99 | Other disorders of the nervous system |
| **Chapter IX** Disorders of the circulatory system | I00-I02 | Acute rheumatic fever |
| | I05-I09 | Chronic rheumatic heart diseases |
| | I10-I15 | Hypertensive diseases |
| | I20-I25 | Ischaemic heart diseases |
| | I26-I28 | Pulmonary heart disease and diseases of pulmonary circulation |
| | I30-I52 | Other forms of heart disease |
| | I60-I69 | Cerebrovascular diseases |
| | I70-I79 | Diseases of arteries, arterioles and capillaries |
| | I80-I89 | Diseases of veins, lymphatic vessels and lymph nodes, not elsewhere classified |
| | I95-I99 | Other and unspecified disorders of the circulatory system |
| **Chapter X** Disorders of the respiratory system | J00-J06 | Acute upper respiratory infections |
| | J09-J18 | Influenza and pneumonia |
| | J20-J22 | Other acute lower respiratory infections |
| | J30-J39 | Other diseases of upper respiratory tract |
| | J40-J47 | Chronic lower respiratory diseases |
| | J60-J70 | Lung diseases due to external agents |
| | J80-J84 | Other respiratory diseases principally affecting the interstitium |

| | | |
|---|---|---|
| | J85-J86 | Suppurative and necrotic conditions of lower respiratory tract |
| | J90-J94 | Other diseases of pleura |
| | J95-J99 | Other diseases of the respiratory system |
| **Chapter XI** Disorders of the digestive system | K00-K14 | Diseases of oral cavity, salivary glands and jaws |
| | K20-K31 | Diseases of oesophagus, stomach and duodenum |
| | K35-K38 | Diseases of appendix |
| | K40-K46 | Hernia |
| | K50-K52 | Noninfective enteritis and colitis |
| | K55-K64 | Other diseases of intestines |
| | K65-K67 | Diseases of peritoneum |
| | K70-K77 | Diseases of liver |
| | K80-K87 | Disorders of gallbladder, biliary tract and pancreas |
| | K90-K93 | Other diseases of the digestive system |
| **Chapter XII** Disorders of the skin and subcutaneous tissue | L00-L08 | Infections of the skin and subcutaneous tissue |
| | L10-L14 | Bullous disorders |
| | L20-L30 | Dermatitis and eczema |
| | L40-L45 | Papulosquamous disorders |
| | L50-L54 | Urticaria and erythema |
| | L55-L59 | Radiation-related disorders of the skin and subcutaneous tissue |
| | L60-L75 | Disorders of skin appendages |
| | L80-L99 | Other disorders of the skin and subcutaneous tissue |
| **Chapter XIII** Disorders of the musculoskeletal system and connective tissue | M00-M03 | Infectious arthropathies |
| | M05-M14 | Inflammatory polyarthropathies |
| | M15-M19 | Arthrosis |
| | M20-M25 | Other joint disorders |
| | M30-M36 | Systemic connective tissue disorders |
| | M40-M43 | Deforming dorsopathies |
| | M45-M49 | Spondylopathies |
| | M50-M54 | Other dorsopathies |
| | M60-M63 | Disorders of muscles |
| | M65-M68 | Disorders of synovium and tendon |
| | M70-M79 | Other soft tissue disorders |
| | M80-M85 | Disorders of bone density and structure |
| | M86-M90 | Other osteopathies |
| | M91-M94 | Chondropathies |
| | M95-M99 | Other disorders of the musculoskeletal system and connective tissue |
| **Chapter XIV** Disorders of the genitourinary system | N00-N08 | Glomerular diseases |
| | N10-N16 | Renal tubulo-interstitial diseases |
| | N17-N19 | Renal failure |
| | N20-N23 | Urolithiasis |
| | N25-N29 | Other disorders of kidney and ureter |
| | N30-N39 | Other diseases of urinary system |
| | N40-N51 | Diseases of male genital organs |
| | N60-N64 | Disorders of breast |
| | N70-N77 | Inflammatory diseases of female pelvic organs |
| | N80-N98 | Noninflammatory disorders of female genital tract |
| | N99-N99 | Other disorders of the genitourinary system |

*Table S2.* **Comparison of sample characteristics**

| **Variables** | | | **CHAPTERS** | | | | | | | | | | | |
|---|---|---|---|---|---|---|---|---|---|---|---|---|---|---|
| | | **ELSA** | **V** | **V (F51) & VI (G47)** | **III** | **IX** | **XI** | **IV** | **XIV** | **I** | **XIII** | **VI** | **X** | **XII** |
| | | ***n*** **(%) \| M ±** | ***n*** **(%) \| M ±** | ***n*** **(%) \| M ±** | ***n*** **(%) \| M ±** | ***n*** **(%) \| M ±** | ***n*** **(%) \| M ±** | ***n*** **(%) \| M ±** | ***n*** **(%) \| M ±** | ***n*** **(%) \| M ±** | ***n*** **(%) \| M ±** | ***n*** **(%) \| M ±** | ***n*** **(%) \| M ±** | ***n*** **(%) \| M ±** |
| | | 4,940 | 4,181 | 4,276 | 4,139 | 3,163 | 3,188 | 3,749 | 3,524 | 4,118 | 3,441 | 4,057 | 3,846 | 4,045 |
| Age | | 66.3 ±9.4 | 66.7 ±9.3 | 66.7 ±9.3 | 66.5 ±9.3 | 65.3 ±8.7 | 65.8 ±9.2 | 66.2 ±9.2 | 66.5 ±9.3 | 66.5 ±9.3 | 66.1 ±9.2 | 66.5 ±9.3 | 66.4 ±9.2 | 66.7 ±9.3 |
| Age (Binary) | < Md | 2,436 (49.31) | 2,007 (48.00) | 2047 (47.87) | 2,008 (48.51) | 1,713 (54.16) | 1,667 (52.29) | 1,884 (50.25) | 1,721 (48.84) | 1,990 (48.32) | 1,724 (50.10) | 1,957 (48.24) | 1,884 (48.99) | 1,949 (48.18) |
| | ≥ Md | 2,504 (50.69) | 2,174 (52.00) | 2229 (52.13) | 2,131 (51.49) | 1,450 (45.84) | 1,521 (47.71) | 1,865 (49.75) | 1,803 (51.16) | 2,128 (51.68) | 1,717 (49.90) | 2,100 (51.76) | 1,962 (51.01) | 2,096 (51.82) |
| Sex | Male | 2,237 (45.28) | 1,904 (45.54) | 1,949 (45.58) | 1,897 (45.83) | 1,404 (44.39) | 1,428 (44.79) | 1,720 (45.88) | 1,694 (48.07) | 1,889 (45.87) | 1,595 (46.35) | 1,864 (45.95) | 1,749 (45.48) | 1,857 (45.91) |
| | Female | 2,703 (54.72) | 2,277 (54.46) | 2,327 (54.42) | 2,242 (54.17) | 1,759 (55.61) | 1,760 (55.21) | 2,029 (54.12) | 1,830 (51.93) | 2,229 (54.13) | 1,846 (53.65) | 2,193 (54.05) | 2,097 (54.52) | 2,188 (54.09) |
| Education | Higher | 1,589 (32.17) | 1,370 (32.77) | 1,388 (32.46) | 1,364 (32.95) | 1,099 (34.75) | 1,084 (34.00) | 1,265 (33.74) | 1,181 (33.51) | 1,357 (32.95) | 1,157 (33.62) | 1,325 (32.66) | 1,294 (33.65) | 1,314 (32.48) |
| | Primary/ Secondary/ Tertiary | 1,543 (31.23) | 1,303 (31.16) | 1,335 (31.22) | 1,286 (31.07) | 1,013 (32.03) | 1,019 (31.96) | 1,190 (31.74) | 1,108 (31.44) | 1,285 (31.20) | 1,095 (31.82) | 1,270 (31.30) | 1,196 (31.10) | 1,273 (31.47) |
| | Alternative/ None | 1,808 (36.60) | 1,508 (36.07) | 1,553 (36.32) | 1,489 (35.97) | 1,051 (33.23) | 1,019 (31.96) | 1,294 (34.52) | 1,235 (35.05) | 1,476 (35.84) | 1,189 (34.55) | 1,462 (36.04) | 1,356 (35.26) | 1,458 (36.04) |
| Wealth | Lowest | 1,573 (31.84) | 1,388 (33.20) | 1,448 (33.86) | 1,383 (33.41) | 976 (30.86) | 1,010 (31.68) | 1,201 (32.04) | 1,165 (33.06) | 1,373 (33.34) | 1,082 (31.44) | 1,347 (33.20) | 1,237 (32.16) | 1,357 (33.55) |
| | Middle | 2,014 (40.77) | 1,753 (41.93) | 1,786 (41.77) | 1,732 (41.85) | 1,343 (42.46) | 1,339 (42.00) | 1,583 (42.22) | 1,470 (41.71) | 1,719 (41.74) | 1,458 (42.37) | 1,709 (42.12) | 1,634 (42.49) | 1,692 (41.83) |
| | Highest | 1,353 (27.39) | 1,040 (24.87) | 1,042 (24.37) | 1,024 (24.74) | 844 (26.68) | 839 (26.32) | 965 (25.74) | 889 (25.23) | 1,026 (24.92) | 901 (26.18) | 1,001 (24.67) | 975 (25.35) | 996 (24.62) |
| Smoking Status | Never/ Ex-Smokers | 4,312 (87.29) | 3,673 (87.85) | 3,736 (87.37) | 3,614 (87.32) | 2,744 (86.75) | 2,768 (86.83) | 3,278 (87.44) | 3,082 (87.46) | 3,596 (87.32) | 3,005 (87.33) | 3,545 (87.38) | 3,383 (87.96) | 3,545 (87.64) |
| | Current Smoker | 628 (12.71) | 508 (12.15) | 540 (12.63) | 525 (12.68) | 419 (13.25) | 420 (13.17) | 471 (12.56) | 442 (12.54) | 522 (12.68) | 436 (12.67) | 512 (12.62) | 463 (12.04) | 500 (12.36) |
| Alcohol Consumption | <3 days a week | 3,175 (64.27) | 2,680 (64.10) | 2,748 (64.27) | 2,642 (63.83) | 1,977 (62.50) | 2,013 (63.14) | 2,364 (63.06) | 2,219 (62.97) | 2,633 (63.94) | 2,161 (62.80) | 2,582 (63.64) | 2,446 (63.60) | 2,589 (64.00) |
| | ≥3 days a week | 1,765 (35.73) | 1,501 (35.90) | 1,528 (35.73) | 1,497 (36.17) | 1,186 (37.50) | 1,175 (36.86) | 1,385 (36.94) | 1,305 (37.03) | 1,485 (36.06) | 1,280 (37.20) | 1,475 (36.36) | 1,400 (36.40) | 1,456 (36.00) |
| Mobility | Mobile | 2,678 (54.21) | 2,291 (54.80) | 2,363 (55.26) | 2,252 (54.41) | 1,534 (48.50) | 1,665 (52.23 | 1,952 (52.07) | 1,894 (53.75) | 2,240 (54.40) | 1,708 (49.64) | 2,182 (53.78) | 2,029 (52.76) | 2,202 (54.44) |
| | Not Mobile | 2,262 (45.79) | 1,890 (45.20) | 1,913 (44.74) | 1,887 (45.59) | 1,629 (51.50) | 1,523 (47.77) | 1,797 (47.93) | 1,630 (46.25) | 1,878 (45.60) | 1,733 (50.36) | 1,875 (46.22) | 1,817 (47.24) | 1,843 (45.56) |
| Medication | Not Medicated | 4,887 (98.93) | 4,139 (99.00) | 4,230 (98.92) | 4,098 (99.01) | 3,142 (99.34) | 3,163 (99.22) | 3,720 (99.23) | 3,487 (98.95) | 4,074 (98.93) | 3,415 (99.24) | 4,020 (99.09) | 3,814 (99.17) | 4,003 (98.96) |
| | Medicated | 53 (1.07) | 42 ( 1.00) | 46 ( 1.08) | 41 (0.99) | 21 ( 0.66) | 25 (0.78) | 29 (0.77) | 37 (1.05) | 44 (1.07) | 26 (0.76) | 37 (0.91) | 32 (0.83) | 42 (1.04) |
| Physical Activity | Sedentary | 1,340 (27.13) | 1,120 (26.79) | 1,169 (27.34) | 1,109 (26.79) | 737 (23.30) | 785 (24.62) | 941 (25.10) | 922 (26.16) | 1,094 (26.57) | 837 (24.32) | 1,079 (26.60) | 970 (25.22) | 1,079 (26.67) |
| | Active | 3,600 (72.87) | 3,061 (73.21) | 3,107 (72.66) | 3,030 (73.21) | 2,426 (76.70) | 2,403 (75.38) | 2,808 (74.90) | 2,602 (73.84) | 3,024 (73.43) | 2,604 (75.68) | 2,978 (73.40) | 2,876 (74.78) | 2,966 (73.33) |
| BMI | <25/ Normal | 1,312 (26.56) | 1,085 (25.95) | 1,110 (25.96) | 1,070 (25.85) | 894 (28.26) | 879 (27.57) | 1,013 (27.02) | 933 (26.48) | 1,080 (26.23) | 950 (27.61) | 1,076 (26.52) | 1,009 (26.24) | 1,057 (26.13) |
| | 25-30, Overweight: Pre-obese | 2,213 (44.80) | 1,877 (44.89) | 1,919 (44.88) | 1,866 (45.08) | 1,432 (45.27) | 1,393 (43.70) | 1,692 (45.13) | 1,581 (44.86) | 1,844 (44.78) | 1,552 (45.10) | 1,813 (44.69) | 1,742 (45.29) | 1,822 (45.04) |
| | ≥30, Obese | 1,415 (28.64) | 1,219 (29.16) | 1,247 (29.16) | 1,203 (29.06) | 837 (26.46) | 916 (28.73) | 1,044 (27.85) | 1,010 (28.66) | 1,194 (28.99) | 939 (27.29) | 1,168 (28.79) | 1,095 (28.47) | 1,166 (28.83) |
| Health | No health condition | 3,319 (67.19) | 2,754 (65.87) | 2,809 (65.69) | 2,744 (66.30) | 2,245 (70.98) | 2,216 (69.51) | 2,567 (68.47) | 2,369 (67.22) | 2,732 (66.34) | 2,421 (70.36) | 2,705 (66.67) | 2,629 (68.36) | 2,685 (66.38) |
| | At least one health condition | 1,621 (32.81) | 1,427 (34.13) | 1,467 (34.31) | 1,395 (33.70) | 918 (29.02) | 972 (30.49) | 1,182 (31.53) | 1,155 (32.78) | 1,386 (33.66) | 1,020 (29.64) | 1,352 (33.33) | 1,217 (31.64) | 1,360 (33.62) |
| CRP* (mg/L) | | 1.2 ±0.7 | 1.2 ±0.7 | 1.2 ±0.7 | 1.2 ±0.7 | 1.2 ±0.7 | 1.2 ±0.7 | 1.2 ±0.7 | 1.2 ±0.7 | 1.2 ±0.7 | 1.2 ±0.7 | 1.2 ±0.7 | 1.2 ±0.7 | 1.2 ±0.7 |
| Fb (g/L) | | 3.4 ±0.6 | 3.4 ±0.6 | 3.4 ±0.6 | 3.4 ±0.6 | 3.4 ±0.5 | 3.4 ±0.6 | 3.4 ±0.6 | 3.4 ±0.6 | 3.4 ±0.6 | 3.4 ±0.6 | 3.4 ±0.6 | 3.4 ±0.5 | 3.4 ±0.6 |
| WBCC* (nmol/L) | | 2.0 ±0.3 | 2.0 ±0.3 | 2.0 ±0.3 | 2.0 ±0.3 | 2.0 ±0.3 | 2.0 ±0.3 | 2.0 ±0.3 | 2.0 ±0.3 | 2.0 ±0.3 | 2.0 ±0.3 | 2.0 ±0.3 | 2.0 ±0.3 | 2.0 ±0.3 |
| IGF-1* (nmol/L) | | 2.8 ±0.3 | 2.8 ±0.3 | 2.8 ±0.3 | 2.8 ±0.3 | 2.8 ±0.3 | 2.8 ±0.3 | 2.8 ±0.3 | 2.8 ±0.3 | 2.8 ±0.3 | 2.8 ±0.3 | 2.8 ±0.3 | 2.8 ±0.3 | 2.8 ±0.3 |
| I-N Profiles | *Low-risk* | 2,590 (52.43) | 2,199 (52.60) | 2,234 (52.25) | 2,176 (52.57) | 1,739 (54.98) | 1,728 (54.20) | 1,994 (53.19) | 1,897 (53.83) | 2,176 (52.84) | 1,887 (54.84) | 2,128 (52.45) | 2,075 (53.95) | 2,145 (53.03) |
| | *Moderate-risk* | 1,773 (35.89) | 1,493 (35.71) | 1,535 (35.90) | 1,481 (35.78) | 1,091 (34.49) | 1,113 (34.91) | 1,330 (35.48) | 1,234 (35.02) | 1,467 (35.62) | 1,189 (34.55) | 1,453 (35.81) | 1,356 (35.26) | 1,441 (35.62) |
| | *High-risk* | 577 (11.68) | 489 (11.70) | 507 (11.86) | 482 (11.65) | 333 (10.53) | 347 (10.88) | 425 (11.34) | 393 (11.15) | 475 (11.53) | 365 (10.61) | 476 (11.73) | 415 (10.79) | 459 (11.35) |

Notes: ELSA = English Longitudinal Study of Ageing; Chapter V = Mental and behavioural disorders; Chapter V (F51) and Chapter VI (G47) = Sleep disorders; Chapter III = Diseases of the blood and blood-forming organs and certain disorders involving the immune mechanism; Chapter IX = Diseases of the circulatory system; Chapter XI = Diseases of the digestive system; Chapter IV = Endocrine, nutritional, and metabolic diseases; Chapter XIV = Diseases of the genitourinary system; Chapter I = Infectious and parasitic diseases; Chapter XIII = Diseases of the musculoskeletal system and connective tissue; Chapter VI = Diseases of the nervous system; Chapter X = Diseases of the respiratory system; Chapter XII = Diseases of the skin and subcutaneous tissue; *n* = observations; M = Mean; Md = Median; % = percentage frequencies; ± = standard deviations; < = less than; ≥ = greater than or equal to; BMI = Body Mass Index; PGS = Polygenic Score; CRP = C-reactive protein; Fb = Fibrinogen; WBCC = White Blood Cell Counts; IGF-1 = Insulin-growth factor-1; * Log-transformed variable; I-N = Immune and Neuroendocrine.

*Table S3.* **Sample size and size difference, along with the number of failures and time to risk.**

| Disorder Chapter | *n* | Difference | Failures | Time at Risk |
|---|---|---|---|---|
| **ELSA Sample** | **4,940** | - | - | - |
| **Chapter V (F51) and Chapter VI (G47)** Sleep disorders | 4,276 | 664 | 49 | 36,019.76 |
| **Chapter V** Psychiatric (mental and behavioural) disorders | 4,181 | 759 | 740 | 33,316.10 |
| **Chapter III** Diseases of the blood and blood-forming organs and certain disorders involving the immune mechanism | 4,139 | 801 | 449 | 33,870.95 |
| **Chapter I** Infectious and parasitic diseases | 4,118 | 822 | 553 | 33,474.10 |
| **Chapter VI** Diseases of the nervous system | 4,057 | 883 | 539 | 32,794.69 |
| **Chapter XII** Diseases of the skin and subcutaneous tissue | 4,045 | 895 | 472 | 32,794.12 |
| **Chapter X** Diseases of the respiratory system | 3,846 | 1,094 | 896 | 30,151.82 |
| **Chapter IV** Endocrine, nutritional, and metabolic diseases | 3,749 | 1,191 | 1,174 | 27,624.52 |
| **Chapter XIV** Diseases of the genitourinary system | 3,524 | 1,416 | 906 | 26,907.90 |
| **Chapter XIII** Diseases of the musculoskeletal system and connective tissue | 3,441 | 1,499 | 1,080 | 25,155.85 |
| **Chapter XI** Diseases of the digestive system | 3,188 | 1,752 | 1,155 | 22,686.37 |
| **Chapter IX** Diseases of the circulatory system | 3,163 | 1,777 | 1,406 | 21,543.72 |

***Table S4.*** **Correlations between biomarkers and polygenic scores**

| | **CRP** | **Fb** | **WBCC** | **IGF-1** | **PGS CRP** | **PGS WBCC** | **PGS IGF-1** | **PGS Anxiety** | **PGS Depression** | **PGS Schizophrenia** | **PGS Insomnia** | **PGS Pain** |
|---|---|---|---|---|---|---|---|---|---|---|---|---|
| **CRP** | 1 | | | | | | | | | | | |
| **Fb** | **0.567*** | 1 | | | | | | | | | | |
| | *<0.001* | | | | | | | | | | | |
| **WBCC** | **0.301*** | **0.245*** | 1 | | | | | | | | | |
| | *<0.001* | *<0.001* | | | | | | | | | | |
| **IGF-1** | **-0.164*** | **0.051*** | 0.003 | 1 | | | | | | | | |
| | *<0.001* | *0.001* | *0.831* | | | | | | | | | |
| **PGS CRP** | **0.043*** | -0.005 | 0.006 | **-0.035*** | 1 | | | | | | | |
| | *0.005* | *0.734* | *0.674* | *0.024* | | | | | | | | |
| **PGS WBCC** | 0.027 | -0.009 | **0.085*** | 0.024 | 0.001 | 1 | | | | | | |
| | *0.078* | *0.556* | *<0.001* | *0.111* | *0.972* | | | | | | | |
| **PGS IGF-1** | 0.018 | **0.034*** | 0.004 | 0.023 | -0.019 | 0.018 | 1 | | | | | |
| | *0.242* | *0.026* | *0.803* | *0.126* | *0.207* | *0.234* | | | | | | |
| **PGS Anxiety** | 0.004 | -0.002 | 0.027 | 0.002 | -0.011 | **0.578*** | 0.014 | 1 | | | | |
| | *0.786* | *0.883* | *0.073* | *0.915* | *0.476* | *<0.001* | *0.349* | | | | | |
| **PGS Depression** | 0.029 | 0.008 | **0.047*** | -0.003 | -0.021 | **0.273*** | 0.008 | **0.213*** | 1 | | | |
| | *0.061* | *0.584* | *0.002* | *0.861* | *0.164* | *<0.001* | *0.623* | *<0.001* | | | | |
| **PGS Schizophrenia** | 0.025 | 0.006 | **0.040*** | 0.022 | -0.023 | **0.789*** | 0.008 | **0.557*** | **0.275*** | 1 | | |
| | *0.101* | *0.679* | *0.009* | *0.151* | *0.126* | *<0.001* | *0.619* | *<0.001* | *<0.001* | | | |
| **PGS Insomnia** | -0.006 | 0.019 | 0.002 | 0.013 | 0.024 | **-0.032*** | -0.029 | **-0.103*** | **-0.037*** | **-0.124*** | 1 | |
| | *0.698* | *0.224* | *0.906* | *0.386* | *0.124* | *0.036* | *0.058* | *<0.001* | *0.015* | *<0.001* | | |
| **PGS Pain** | **0.075*** | **0.059*** | **0.054*** | -0.004 | -0.020 | **0.215*** | **-0.031*** | **0.167*** | **0.267*** | **0.214*** | **0.043*** | 1 |
| | *<0.001* | *<0.001* | *<0.001* | *0.777* | *0.202* | *<0.001* | *0.040* | *<0.001* | *<0.001* | *<0.001* | *0.005* | |

**Notes:** PGS = Polygenic Score; CRP = C-reactive protein; Fb = Fibrinogen; WBCC = White Blood Cell Counts; IGF-1 = Insulin-growth factor-1.

***Table S5.*** **Latent Profile Analysis**

| | **One Profile** | **Two Profiles** | **Three Profiles** | **Four Profiles** | **Five Profiles** | **Six Profiles** | **Seven Profiles** |
|---|---|---|---|---|---|---|---|
| AIC | 21856.03 | 19459.96 | 18745.36 | 18512.47 | 18412.80 | 18318.50 | 18231.59 |
| AIC Difference (N) | - | 2396.07 | 714.60 | 232.89 | 99.67 | 94.30 | 86.91 |
| AIC Difference (%) | - | 12.31 | 3.81 | 1.26 | 0.54 | 0.51 | 0.48 |
| BIC | 21908.07 | 19544.53 | 18862.45 | 18662.08 | 18594.94 | 18533.17 | 18478.78 |
| BIC Difference (N) | - | 2363.54 | 682.08 | 200.37 | 67.14 | 61.77 | 54.39 |
| BIC Difference (%) | - | 12.09 | 3.62 | 1.07 | 0.36 | 0.33 | 0.29 |
| aBIC | 21882.65 | 19503.22 | 18805.26 | 18589.00 | 18505.97 | 18428.31 | 18358.03 |
| aBIC Difference (N) | - | 2379.43 | 697.97 | 216.26 | 83.03 | 77.66 | 70.28 |
| aBIC Difference (%) | - | 12.20 | 3.71 | 1.16 | 0.45 | 0.42 | 0.38 |
| Entropy | - | 0.78 | 0.67 | 0.67 | 0.67 | 0.65 | 0.62 |
| Normalised Entropy | - | 0.71 | 0.57 | 0.57 | 0.57 | 0.56 | 0.54 |
| M Posterior Probabilities (SE) | - | 0.753 (.010) | 0.519 (.012) | 0.407 (.014) | 0.320 (.016) | 0.373 (.014) | 0.289 (.025) |
| | | 0.247 (.010) | 0.359 (.011) | 0.350 (.011) | 0.297 (.013) | 0.327 (.012) | 0.130 (.031) |
| | | | 0.122 (.007) | 0.159 (.011) | 0.214 (.013) | 0.017 (.006) | 0.282 (.019) |
| | | | | 0.084 (.007) | 0.105 (.009) | 0.172 (.011) | 0.118 (.013) |
| | | | | | 0.065 (.007) | 0.084 (.007) | 0.076 (.017) |
| | | | | | | 0.026 (.005) | 0.084 (.006) |
| | | | | | | | 0.021 (.004) |
| N classes >5% | Yes | Yes | Yes | Yes | Yes | No | No |

***Table S6.*** **Latent Profile Predicted Means and Percentage Point Change**

| | | Profile 1 | Profile 2 | Profile 3 |
|---|---|---|---|---|
| Predicted Means | CRP* | 0.69 | 1.5 | 2.43 |
| | **% Change** | - | **54%** | **38%** |
| | Fb | 3.11 | 3.5 | 4.17 |
| | **% Change** | - | **11%** | **16%** |
| | WBCC* | 1.9 | 2.01 | 2.13 |
| | **% Change** | - | **5%** | **6%** |
| | IGF-1* | 2.82 | 2.73 | 2.71 |
| | **% Change** | - | **-3%** | **-1%** |

**Notes:** % = percentage frequencies; CRP = C-reactive protein; Fb = Fibrinogen; WBCC = White Blood Cell Counts; IGF-1 = Insulin-growth factor-1; * Log-transformed variable.

*Table S7.* **Fully adjusted longitudinal associations between immune and neuroendocrine profiles and all-cause hospitalisation, with *n* distribution.**

| Adjustments | Immune-Neuroendocrine Profiles | | | | |
|---|---|---|---|---|---|
| | HR | SE | 95% CI | | *p* |
| ***Psychiatric Disorders*** **(*n*=4,181)** | | | | | |
| Moderate Level - Model 3 [a] | 1.12 | 0.09 | 0.95 | 1.32 | 0.182 |
| High Level - Model 3 [a] | 1.30 | 0.15 | 1.05 | 1.62 | 0.018 |
| ***Sleep Disorders*** **(*n*=4,276)** | | | | | |
| Moderate Level - Model 3 [a] | 2.29 | 0.77 | 1.19 | 4.41 | 0.013 |
| High Level - Model 3 [a] | 3.54 | 1.49 | 1.56 | 8.08 | 0.003 |
| ***Blood Disorders*** **(*n*=4,139)** | | | | | |
| Moderate Level - Model 3 [a] | 1.10 | 0.12 | 0.89 | 1.36 | 0.388 |
| High Level - Model 3 [a] | 1.68 | 0.23 | 1.28 | 2.19 | <0.001 |
| ***Circulatory Disorders*** **(*n*=3,163)** | | | | | |
| Moderate Level - Model 3 [a] | 1.26 | 0.08 | 1.12 | 1.42 | <0.001 |
| High Level - Model 3 [a] | 1.37 | 0.12 | 1.16 | 1.63 | <0.001 |
| ***Digestive Disorders*** **(*n*=3,188)** | | | | | |
| Moderate Level - Model 3 [a] | 1.04 | 0.07 | 0.92 | 1.18 | 0.545 |
| High Level - Model 3 [a] | 0.93 | 0.09 | 0.76 | 1.13 | 0.460 |
| ***Endocrine Disorders*** **(*n*=3,749)** | | | | | |
| Moderate Level - Model 3 [a] | 1.26 | 0.08 | 1.11 | 1.43 | <0.001 |
| High Level - Model 3 [a] | 1.38 | 0.13 | 1.16 | 1.66 | <0.001 |
| ***Genitourinary Disorders*** **(*n*=3,524)** | | | | | |
| Moderate Level - Model 3 [a] | 1.18 | 0.09 | 1.02 | 1.36 | 0.027 |
| High Level - Model 3 [a] | 1.35 | 0.14 | 1.10 | 1.65 | 0.004 |
| ***Infectious Disorders*** **(*n*=4,118)** | | | | | |
| Moderate Level - Model 3 [a] | 1.02 | 0.10 | 0.85 | 1.23 | 0.810 |
| High Level - Model 3 [a] | 1.38 | 0.18 | 1.08 | 1.78 | 0.011 |
| ***Musculoskeletal Disorders*** **(*n*=3,441)** | | | | | |
| Moderate Level - Model 3 [a] | 1.13 | 0.08 | 0.99 | 1.30 | 0.063 |
| High Level - Model 3 [a] | 1.28 | 0.13 | 1.06 | 1.55 | 0.012 |
| ***Nervous Disorders*** **(*n*=4,057)** | | | | | |
| Moderate Level - Model 3 [a] | 1.12 | 0.11 | 0.93 | 1.35 | 0.220 |
| High Level - Model 3 [a] | 0.97 | 0.14 | 0.73 | 1.28 | 0.808 |
| ***Respiratory Disorders*** **(*n*=3,846)** | | | | | |
| Moderate Level - Model 3 [a] | 1.26 | 0.10 | 1.09 | 1.47 | 0.002 |
| High Level - Model 3 [a] | 1.99 | 0.19 | 1.64 | 2.40 | <0.001 |
| ***Skin Disorders*** **(*n*=4,045)** | | | | | |
| Moderate Level - Model 3 [a] | 1.00 | 0.10 | 0.82 | 1.23 | 0.968 |
| High Level - Model 3 [a] | 1.19 | 0.17 | 0.90 | 1.58 | 0.231 |

**Notes:** The *low-risk* group is the reference; HR = Hazard ratio; SE = standard errors; CI = confidence interval; *p* = significance value.
a All variables: Baseline immune and neuroendocrine profiles; age; sex; 10 PCs; CRP PGS; WBCC PGS; IGF-1 PGS; PGS Anxiety; PGS Depression; PGS Schizophrenia; PGS Insomnia; PGS; Pain; education; wealth; smoking status; alcohol consumption; mobility

*Table S8.* **False discovery rate (FDR) adjusted longitudinal associations of immune-neuroendocrine profiles and individual biomarkers with all-cause hospitalisation (*n*=4,181)**

| Disorder | Risk Level | Exposure | Alpha | | FDR | | Change |
|---|---|---|---|---|---|---|---|
| | | | p value | Significant | q value | Significant | |
| Psychiatric | Moderate | Profiles | 0.182 | FALSE | 0.273 | FALSE | FALSE |
| Psychiatric | High | Profiles | 0.018 | TRUE | 0.033 | TRUE | FALSE |
| Sleep | Moderate | Profiles | 0.013 | TRUE | 0.026 | TRUE | FALSE |
| Sleep | High | Profiles | 0.003 | TRUE | 0.009 | TRUE | FALSE |
| Blood | Moderate | Profiles | 0.388 | FALSE | 0.490 | FALSE | FALSE |
| Blood | High | Profiles | <0.001 | TRUE | <0.001 | TRUE | FALSE |
| Circulatory | Moderate | Profiles | <0.001 | TRUE | <0.001 | TRUE | FALSE |
| Circulatory | High | Profiles | <0.001 | TRUE | <0.001 | TRUE | FALSE |
| Digestive | Moderate | Profiles | 0.545 | FALSE | 0.623 | FALSE | FALSE |
| Digestive | High | Profiles | 0.460 | FALSE | 0.552 | FALSE | FALSE |
| Endocrine | Moderate | Profiles | <0.001 | TRUE | <0.001 | TRUE | FALSE |
| Endocrine | High | Profiles | <0.001 | TRUE | <0.001 | TRUE | FALSE |
| Genitourinary | Moderate | Profiles | 0.027 | TRUE | 0.046 | TRUE | FALSE |
| Genitourinary | High | Profiles | 0.004 | TRUE | 0.011 | TRUE | FALSE |
| Infectious | Moderate | Profiles | 0.810 | FALSE | 0.845 | FALSE | FALSE |
| Infectious | High | Profiles | 0.011 | TRUE | 0.026 | TRUE | FALSE |
| Musculoskeletal | Moderate | Profiles | 0.063 | FALSE | 0.101 | FALSE | FALSE |
| Musculoskeletal | High | Profiles | 0.012 | TRUE | 0.026 | TRUE | FALSE |
| Nervous | Moderate | Profiles | 0.220 | FALSE | 0.308 | FALSE | FALSE |
| Nervous | High | Profiles | 0.808 | FALSE | 0.845 | FALSE | FALSE |
| Respiratory | Moderate | Profiles | 0.002 | TRUE | 0.007 | TRUE | FALSE |
| Respiratory | High | Profiles | <0.001 | TRUE | <0.001 | TRUE | FALSE |
| Skin | Moderate | Profiles | 0.968 | FALSE | 0.968 | FALSE | FALSE |
| Skin | High | Profiles | 0.231 | FALSE | 0.308 | FALSE | FALSE |
| Blood | n/a | CRP | 0.001 | TRUE | 0.004 | TRUE | FALSE |
| Blood | n/a | Fb | 0.007 | TRUE | 0.021 | TRUE | FALSE |
| Blood | n/a | IGF1 | 0.349 | FALSE | 0.487 | FALSE | FALSE |
| Blood | n/a | WBCC | 0.052 | FALSE | 0.113 | FALSE | FALSE |
| Circulatory | n/a | CRP | 5e-04 | TRUE | 0.003 | TRUE | FALSE |
| Circulatory | n/a | Fb | <0.001 | TRUE | <0.001 | TRUE | FALSE |
| Circulatory | n/a | IGF1 | 0.273 | FALSE | 0.400 | FALSE | FALSE |
| Circulatory | n/a | WBCC | 5e-04 | TRUE | 0.003 | TRUE | FALSE |
| Digestive | n/a | CRP | 0.620 | FALSE | 0.775 | FALSE | FALSE |
| Digestive | n/a | Fb | 0.580 | FALSE | 0.773 | FALSE | FALSE |
| Digestive | n/a | IGF1 | 0.962 | FALSE | 0.962 | FALSE | FALSE |
| Digestive | n/a | WBCC | 0.962 | FALSE | 0.962 | FALSE | FALSE |
| Endocrine | n/a | CRP | 5e-04 | TRUE | 0.003 | TRUE | FALSE |

| | | | | | | | |
|---|---|---|---|---|---|---|---|
| Endocrine | n/a | Fb | <0.001 | TRUE | <0.001 | TRUE | FALSE |
| Endocrine | n/a | IGF1 | 0.106 | FALSE | 0.197 | FALSE | FALSE |
| Endocrine | n/a | WBCC | 0.001 | TRUE | 0.004 | TRUE | FALSE |
| Genitourinary | n/a | CRP | 0.002 | TRUE | 0.007 | TRUE | FALSE |
| Genitourinary | n/a | Fb | 0.009 | TRUE | 0.024 | TRUE | FALSE |
| Genitourinary | n/a | IGF1 | 0.732 | FALSE | 0.817 | FALSE | FALSE |
| Genitourinary | n/a | WBCC | 0.003 | TRUE | 0.010 | TRUE | FALSE |
| Infectious | n/a | CRP | 0.076 | FALSE | 0.159 | FALSE | FALSE |
| Infectious | n/a | Fb | 0.111 | FALSE | 0.197 | FALSE | FALSE |
| Infectious | n/a | IGF1 | 0.761 | FALSE | 0.830 | FALSE | FALSE |
| Infectious | n/a | WBCC | 0.093 | FALSE | 0.186 | FALSE | FALSE |
| Musculoskeletal | n/a | CRP | 5e-04 | TRUE | 0.003 | TRUE | FALSE |
| Musculoskeletal | n/a | Fb | 0.273 | FALSE | 0.400 | FALSE | FALSE |
| Musculoskeletal | n/a | IGF1 | 0.617 | FALSE | 0.775 | FALSE | FALSE |
| Musculoskeletal | n/a | WBCC | 0.197 | FALSE | 0.326 | FALSE | FALSE |
| Nervous | n/a | CRP | 0.630 | FALSE | 0.775 | FALSE | FALSE |
| Nervous | n/a | Fb | 0.866 | FALSE | 0.921 | FALSE | FALSE |
| Nervous | n/a | IGF1 | 0.178 | FALSE | 0.305 | FALSE | FALSE |
| Nervous | n/a | WBCC | 0.703 | FALSE | 0.813 | FALSE | FALSE |
| Psychiatric | n/a | CRP | 0.018 | TRUE | 0.041 | TRUE | FALSE |
| Psychiatric | n/a | Fb | 0.109 | FALSE | 0.197 | FALSE | FALSE |
| Psychiatric | n/a | IGF1 | 0.231 | FALSE | 0.370 | FALSE | FALSE |
| Psychiatric | n/a | WBCC | 0.008 | TRUE | 0.023 | TRUE | FALSE |
| Respiratory | n/a | CRP | 5e-04 | TRUE | 0.003 | TRUE | FALSE |
| Respiratory | n/a | Fb | <0.001 | TRUE | <0.001 | TRUE | FALSE |
| Respiratory | n/a | IGF1 | 0.002 | TRUE | 0.007 | TRUE | FALSE |
| Respiratory | n/a | WBCC | 5e-04 | TRUE | 0.003 | TRUE | FALSE |
| Skin | n/a | CRP | 0.355 | FALSE | 0.487 | FALSE | FALSE |
| Skin | n/a | Fb | 0.883 | FALSE | 0.921 | FALSE | FALSE |
| Skin | n/a | IGF1 | 0.711 | FALSE | 0.813 | FALSE | FALSE |
| Skin | n/a | WBCC | 0.275 | FALSE | 0.400 | FALSE | FALSE |
| Sleep | n/a | CRP | 0.001 | TRUE | 0.004 | TRUE | FALSE |
| Sleep | n/a | Fb | 0.013 | TRUE | 0.031 | TRUE | FALSE |
| Sleep | n/a | IGF1 | 0.700 | FALSE | 0.813 | FALSE | FALSE |
| Sleep | n/a | WBCC | 0.013 | TRUE | 0.031 | TRUE | FALSE |

***Table S9a.*** **Longitudinal associations between immune and neuroendocrine profiles and hospitalisation for mental and behavioural disorders (*n*=4,181)**

| Adjustments | Immune and Neuroendocrine Profiles | | | | |
|---|---|---|---|---|---|
| | HR | SE | 95% CI | | *p* |
| ***Moderate-risk*** **Profile \| Mental and Behavioural Disorders** | | | | | |
| Model 1: *Unadjusted* | 1.52 | 0.12 | 1.30 | 1.78 | <0.001 |
| Model 2: *Model 1 + demographics & genetics* [a] | 1.33 | 0.11 | 1.13 | 1.55 | 0.001 |
| Model 3: *Model 2 + Fully Adjusted* [b] | 1.12 | 0.09 | 0.95 | 1.32 | 0.182 |
| Model 2a: *Demographics* | 1.37 | 0.11 | 1.17 | 1.60 | <0.001 |
| Model 2b: *Genetics* | 1.33 | 0.11 | 1.13 | 1.55 | 0.001 |
| Model 3a: Model 3 + *Medication* | 1.12 | 0.09 | 0.95 | 1.31 | 0.187 |
| Model 3b: Model 3 + *Physical Activity* | 1.10 | 0.09 | 0.93 | 1.29 | 0.269 |
| Model 3c: Model 3 + *Body Mass Index* | 1.09 | 0.09 | 0.92 | 1.28 | 0.339 |
| Model 3d: Model 3 + *Fully Adjusted* [c] | 1.07 | 0.09 | 0.90 | 1.26 | 0.458 |
| Model 4a: Model 2 + *Health* | 1.30 | 0.11 | 1.11 | 1.53 | 0.001 |
| Model 4b: Model 3 + *Health* | 1.11 | 0.09 | 0.94 | 1.30 | 0.223 |
| Model 4c: Model 3 + *Fully Adjusted* [d] | 1.06 | 0.09 | 0.89 | 1.25 | 0.514 |
| ***High-risk*** **Profile \| Mental and Behavioural Disorders** | | | | | |
| Model 1: *Unadjusted* | 1.95 | 0.21 | 1.57 | 2.41 | <0.001 |
| Model 2: *Model 1 + demographics & genetics* [a] | 1.71 | 0.19 | 1.38 | 2.12 | <0.001 |
| Model 3: *Model 2 + Fully Adjusted* [b] | 1.30 | 0.15 | 1.05 | 1.62 | 0.018 |
| Model 2a: *Demographics* | 1.75 | 0.19 | 1.41 | 2.17 | <0.001 |
| Model 2b: *Genetics* | 1.71 | 0.19 | 1.38 | 2.12 | <0.001 |
| Model 3a: Model 3 + *Medication* | 1.31 | 0.15 | 1.05 | 1.63 | 0.016 |
| Model 3b: Model 3 + *Physical Activity* | 1.24 | 0.14 | 1.00 | 1.55 | 0.052 |
| Model 3c: Model 3 + *Body Mass Index* | 1.26 | 0.14 | 1.00 | 1.57 | 0.046 |
| Model 3d: Model 3 + *Fully Adjusted* [c] | 1.21 | 0.14 | 0.96 | 1.51 | 0.101 |
| Model 4a: Model 2 + *Health* | 1.69 | 0.19 | 1.36 | 2.09 | <0.001 |
| Model 4b: Model 3 + *Health* | 1.28 | 0.14 | 1.03 | 1.60 | 0.026 |
| Model 4c: Model 3 + *Fully Adjusted* [d] | 1.19 | 0.14 | 0.95 | 1.50 | 0.125 |

**Notes:** The *low-risk* group is the reference; HR = hazard ratio; SE = standard error; CI = confidence interval; *p* = significance value.

a *Demographic and genetic variables*: age; sex; 10 principal components (PCs); C-reactive Protein (CRP) polygenic score (PGS); White Blood Cell Counts (WBCC) PGS; Insulin Growth Factor-1 (IGF-1) PGS; Anxiety PGS; Depression PGS; Schizophrenia PGS; Insomnia PGS; Pain PGS.

b All variables: age; sex; 10 PCs; CRP PGS; WBCC PGS; IGF-1 PGS; PGS; Anxiety PGS; Depression PGS; Schizophrenia PGS; Insomnia PGS; Pain PGS; education; wealth; smoking status; alcohol consumption; mobility.

c Additional variables: medication, physical activity; BMI.

d Additional variables: health (i.e., chronic lung disease; coronary heart disease; abnormal heart rhythm; heart murmur; congestive heart failure; angina; hypertension; diabetes; cancer; Parkinson's; Alzheimer's; dementia; asthma; arthritis; osteoporosis; psychiatric disorder).

***Table S9b.*** **Longitudinal associations between C-reactive protein and hospitalisation for mental and behavioural disorders (*n*=4,181)**

| Adjustments | CRP | | | | |
|---|---|---|---|---|---|
| | HR | SE | 95% CI | | *p* |
| **Mental and Behavioural Disorders** | | | | | |
| Model 1: *Unadjusted* | 1.40 | 0.07 | 1.27 | 1.56 | <0.001 |
| Model 2: *Model 1 + demographics & genetics* [a] | 1.30 | 0.07 | 1.17 | 1.448 | <0.001 |
| Model 3: *Model 2 + Fully Adjusted* [b] | 1.14 | 0.06 | 1.02 | 1.27 | 0.018 |
| Model 2a: *Demographics* | 1.30 | 0.07 | 1.17 | 1.45 | <0.001 |
| Model 2b: *Genetics* | 1.39 | 0.07 | 1.25 | 1.54 | <0.001 |
| Model 3a: Model 3 + *Medication* | 1.14 | 0.06 | 1.02 | 1.27 | 0.017 |
| Model 3b: Model 3 + *Physical Activity* | 1.11 | 0.06 | 1.00 | 1.24 | 0.053 |
| Model 3c: Model 3 + *Body Mass Index* | 1.12 | 0.07 | 1.00 | 1.25 | 0.059 |
| Model 3d: Model 3 + *Fully Adjusted* [c] | 1.09 | 0.06 | 0.97 | 1.22 | 0.132 |
| Model 4a: Model 2 + *Health* | 1.29 | 0.07 | 1.16 | 1.430 | <0.001 |
| Model 4b: Model 3 + *Health* | 1.13 | 0.06 | 1.01 | 1.259 | 0.029 |
| Model 4c: Model 3 + *Fully Adjusted* [d] | 1.08 | 0.06 | 0.97 | 1.213 | 0.175 |

**Notes:** The *low-risk* group is the reference; HR = hazard ratio; SE = standard error; CI = confidence interval; *p* = significance value.

a *Demographic and genetic variables*: age; sex; 10 principal components (PCs); C-reactive Protein (CRP) polygenic score (PGS); White Blood Cell Counts (WBCC) PGS; Insulin Growth Factor-1 (IGF-1) PGS; Anxiety PGS; Depression PGS; Schizophrenia PGS; Insomnia PGS; Pain PGS.

b All variables: age; sex; 10 PCs; CRP PGS; WBCC PGS; IGF-1 PGS; PGS; Anxiety PGS; Depression PGS; Schizophrenia PGS; Insomnia PGS; Pain PGS; education; wealth; smoking status; alcohol consumption; mobility.

c Additional variables: medication, physical activity; BMI.

d Additional variables: health (i.e., chronic lung disease; coronary heart disease; abnormal heart rhythm; heart murmur; congestive heart failure; angina; hypertension; diabetes; cancer; Parkinson's; Alzheimer's; dementia; asthma; arthritis; osteoporosis; psychiatric disorder).

***Table S9c.*** **Longitudinal associations between fibrinogen and hospitalisation for mental and behavioural disorders (*n*=4,181)**

| **Adjustments** | **Fb** | | | | |
|---|---|---|---|---|---|
| | **HR** | **SE** | **95% CI** | | ***p*** |
| **Mental and Behavioural Disorders** | | | | | |
| Model 1: *Unadjusted* | 1.53 | 0.10 | 1.35 | 1.73 | <0.001 |
| Model 2: *Model 1 + demographics & genetics* [a] | 1.37 | 0.09 | 1.21 | 1.56 | <0.001 |
| Model 3: *Model 2 + Fully Adjusted* [b] | 1.11 | 0.08 | 0.98 | 1.27 | 0.109 |
| Model 2a: *Demographics* | 1.40 | 0.09 | 1.23 | 1.59 | <0.001 |
| Model 2b: *Genetics* | 1.49 | 0.09 | 1.32 | 1.69 | <0.001 |
| Model 3a: Model 3 + *Medication* | 1.11 | 0.08 | 0.98 | 1.27 | 0.109 |
| Model 3b: Model 3 + *Physical Activity* | 1.09 | 0.07 | 0.96 | 1.25 | 0.187 |
| Model 3c: Model 3 + *Body Mass Index* | 1.10 | 0.08 | 0.96 | 1.25 | 0.177 |
| Model 3d: Model 3 + *Fully Adjusted* [c] | 1.08 | 0.07 | 0.94 | 1.23 | 0.275 |
| Model 4a: Model 2 + *Health* | 1.37 | 0.09 | 1.20 | 1.56 | <0.001 |
| Model 4b: Model 3 + *Health* | 1.12 | 0.08 | 0.98 | 1.27 | 0.106 |
| Model 4c: Model 3 + *Fully Adjusted* [d] | 1.08 | 0.07 | 0.94 | 1.23 | 0.262 |

**Notes:** The *low-risk* group is the reference; HR = hazard ratio; SE = standard error; CI = confidence interval; *p* = significance value.

a *Demographic and genetic variables*: age; sex; 10 principal components (PCs); C-reactive Protein (CRP) polygenic score (PGS); White Blood Cell Counts (WBCC) PGS; Insulin Growth Factor-1 (IGF-1) PGS; Anxiety PGS; Depression PGS; Schizophrenia PGS; Insomnia PGS; Pain PGS.

b All variables: age; sex; 10 PCs; CRP PGS; WBCC PGS; IGF-1 PGS; PGS; Anxiety PGS; Depression PGS; Schizophrenia PGS; Insomnia PGS; Pain PGS; education; wealth; smoking status; alcohol consumption; mobility.

c Additional variables: medication, physical activity; BMI.

d Additional variables: health (i.e., chronic lung disease; coronary heart disease; abnormal heart rhythm; heart murmur; congestive heart failure; angina; hypertension; diabetes; cancer; Parkinson's; Alzheimer's; dementia; asthma; arthritis; osteoporosis; psychiatric disorder).

***Table S9d.*** **Longitudinal associations between white blood cell counts and hospitalisation for mental and behavioural disorders (*n*=4,181)**

| Adjustments | WBCC | | | | |
|---|---|---|---|---|---|
| | HR | SE | 95% CI | | *p* |
| **Mental and Behavioural Disorders** | | | | | |
| Model 1: *Unadjusted* | 3.69 | 0.55 | 2.76 | 4.94 | <0.001 |
| Model 2: *Model 1 + demographics & genetics* [a] | 3.66 | 0.56 | 2.72 | 4.92 | <0.001 |
| Model 3: *Model 2 + Fully Adjusted* [b] | 1.53 | 0.25 | 1.12 | 2.09 | 0.008 |
| Model 2a: *Demographics* | 3.45 | 0.52 | 2.57 | 4.63 | <0.001 |
| Model 2b: *Genetics* | 3.82 | 0.57 | 2.85 | 5.12 | <0.001 |
| Model 3a: Model 3 + *Medication* | 1.54 | 0.25 | 1.12 | 2.10 | 0.007 |
| Model 3b: Model 3 + *Physical Activity* | 1.48 | 0.24 | 1.08 | 2.03 | 0.014 |
| Model 3c: Model 3 + *Body Mass Index* | 1.48 | 0.24 | 1.08 | 2.04 | 0.015 |
| Model 3d: Model 3 + *Fully Adjusted* [c] | 1.45 | 0.23 | 1.06 | 1.99 | 0.022 |
| Model 4a: Model 2 + *Health* | 3.57 | 0.54 | 2.65 | 4.80 | <0.001 |
| Model 4b: Model 3 + *Health* | 1.51 | 0.24 | 1.10 | 2.07 | 0.010 |
| Model 4c: Model 3 + *Fully Adjusted* [d] | 1.43 | 0.23 | 1.05 | 1.97 | 0.025 |

**Notes:** The *low-risk* group is the reference; HR = hazard ratio; SE = standard error; CI = confidence interval; *p* = significance value.

a *Demographic and genetic variables*: age; sex; 10 principal components (PCs); C-reactive Protein (CRP) polygenic score (PGS); White Blood Cell Counts (WBCC) PGS; Insulin Growth Factor-1 (IGF-1) PGS; Anxiety PGS; Depression PGS; Schizophrenia PGS; Insomnia PGS; Pain PGS.

b All variables: age; sex; 10 PCs; CRP PGS; WBCC PGS; IGF-1 PGS; PGS; Anxiety PGS; Depression PGS; Schizophrenia PGS; Insomnia PGS; Pain PGS; education; wealth; smoking status; alcohol consumption; mobility.

c Additional variables: medication, physical activity; BMI.

d Additional variables: health (i.e., chronic lung disease; coronary heart disease; abnormal heart rhythm; heart murmur; congestive heart failure; angina; hypertension; diabetes; cancer; Parkinson's; Alzheimer's; dementia; asthma; arthritis; osteoporosis; psychiatric disorder).

***Table S9e.*** **Longitudinal associations between insulin growth factor-1 and hospitalisation for mental and behavioural disorders (*n*=4,181)**

| Adjustments | IGF-1 | | | | |
| --- | --- | --- | --- | --- | --- |
| | HR | SE | 95% CI | | *p* |
| **Mental and Behavioural Disorders** | | | | | |
| Model 1: *Unadjusted* | 0.65 | 0.07 | 0.52 | 0.81 | <0.001 |
| Model 2: *Model 1 + demographics & genetics* [a] | 0.89 | 0.10 | 0.71 | 1.11 | 0.293 |
| Model 3: *Model 2 + Fully Adjusted* [b] | 0.88 | 0.10 | 0.70 | 1.09 | 0.231 |
| Model 2a: *Demographics* | 0.88 | 0.10 | 0.70 | 1.10 | 0.246 |
| Model 2b: *Genetics* | 0.66 | 0.07 | 0.53 | 0.82 | <0.001 |
| Model 3a: Model 3 + *Medication* | 0.87 | 0.10 | 0.70 | 1.09 | 0.220 |
| Model 3b: Model 3 + *Physical Activity* | 0.89 | 0.10 | 0.71 | 1.10 | 0.278 |
| Model 3c: Model 3 + *Body Mass Index* | 0.88 | 0.10 | 0.70 | 1.09 | 0.235 |
| Model 3d: Model 3 + *Fully Adjusted* [c] | 0.89 | 0.10 | 0.71 | 1.10 | 0.276 |
| Model 4a: Model 2 + *Health* | 0.89 | 0.10 | 0.71 | 1.11 | 0.305 |
| Model 4b: Model 3 + *Health* | 0.87 | 0.10 | 0.70 | 1.09 | 0.226 |
| Model 4c: Model 3 + *Fully Adjusted* [d] | 0.89 | 0.10 | 0.71 | 1.10 | 0.270 |

**Notes:** The *low-risk* group is the reference; HR = hazard ratio; SE = standard error; CI = confidence interval; *p* = significance value.

a *Demographic and genetic variables*: age; sex; 10 principal components (PCs); C-reactive Protein (CRP) polygenic score (PGS); White Blood Cell Counts (WBCC) PGS; Insulin Growth Factor-1 (IGF-1) PGS; Anxiety PGS; Depression PGS; Schizophrenia PGS; Insomnia PGS; Pain PGS.

b All variables: age; sex; 10 PCs; CRP PGS; WBCC PGS; IGF-1 PGS; PGS; Anxiety PGS; Depression PGS; Schizophrenia PGS; Insomnia PGS; Pain PGS; education; wealth; smoking status; alcohol consumption; mobility.

c Additional variables: medication, physical activity; BMI.

d Additional variables: health (i.e., chronic lung disease; coronary heart disease; abnormal heart rhythm; heart murmur; congestive heart failure; angina; hypertension; diabetes; cancer; Parkinson's; Alzheimer's; dementia; asthma; arthritis; osteoporosis; psychiatric disorder).

*Table S9f.* **Longitudinal associations between immune and neuroendocrine profiles and hospitalisation for sleep disorders (*n*=4,181)**

| **Adjustments** | **Immune and Neuroendocrine Profiles** | | | | |
|---|---|---|---|---|---|
| | **HR** | **SE** | **95% CI** | | ***p*** |
| ***Moderate-risk* Profile \| Sleep Disorders** | | | | | |
| Model 1: *Unadjusted* | 2.18 | 0.71 | 1.15 | 4.12 | 0.017 |
| Model 2: *Model 1 + demographics & genetics* [a] | 2.33 | 0.77 | 1.22 | 4.46 | 0.010 |
| Model 3: *Model 2 + Fully Adjusted* [b] | 2.29 | 0.77 | 1.19 | 4.41 | 0.013 |
| Model 2a: *Demographics* | 2.47 | 0.81 | 1.30 | 4.69 | 0.006 |
| Model 2b: *Genetics* | 2.08 | 0.68 | 1.09 | 3.94 | 0.025 |
| Model 3a: Model 3 + *Medication* | 2.29 | 0.77 | 1.19 | 4.41 | 0.013 |
| Model 3b: Model 3 + *Physical Activity* | 2.13 | 0.71 | 1.10 | 4.11 | 0.025 |
| Model 3c: Model 3 + *Body Mass Index* | 1.91 | 0.66 | 0.98 | 3.75 | 0.058 |
| Model 3d: Model 3 + *Fully Adjusted* [c] | 1.78 | 0.62 | 0.91 | 3.50 | 0.093 |
| Model 4a: Model 2 + *Health* | 2.33 | 0.77 | 1.22 | 4.46 | 0.010 |
| Model 4b: Model 3 + *Health* | 2.28 | 0.76 | 1.18 | 4.40 | 0.014 |
| Model 4c: Model 3 + *Fully Adjusted* [d] | 1.78 | 0.61 | 0.91 | 3.50 | 0.093 |
| ***High-risk* Profile \| Sleep Disorders** | | | | | |
| Model 1: *Unadjusted* | 3.10 | 1.25 | 1.41 | 6.84 | 0.005 |
| Model 2: *Model 1 + demographics & genetics* [a] | 3.51 | 1.45 | 1.57 | 7.87 | 0.002 |
| Model 3: *Model 2 + Fully Adjusted* [b] | 3.54 | 1.49 | 1.56 | 8.08 | 0.003 |
| Model 2a: *Demographics* | 3.65 | 1.49 | 1.65 | 8.11 | 0.001 |
| Model 2b: *Genetics* | 3.02 | 1.23 | 1.36 | 6.70 | 0.007 |
| Model 3a: Model 3 + *Medication* | 3.54 | 1.49 | 1.56 | 8.08 | 0.003 |
| Model 3b: Model 3 + *Physical Activity* | 3.06 | 1.30 | 1.33 | 7.04 | 0.009 |
| Model 3c: Model 3 + *Body Mass Index* | 2.82 | 1.22 | 1.21 | 6.59 | 0.016 |
| Model 3d: Model 3 + *Fully Adjusted* [c] | 2.44 | 1.07 | 1.04 | 5.75 | 0.041 |
| Model 4a: Model 2 + *Health* | 3.51 | 1.45 | 1.57 | 7.87 | 0.00 |
| Model 4b: Model 3 + *Health* | 3.53 | 1.49 | 1.55 | 8.06 | 0.00 |
| Model 4c: Model 3 + *Fully Adjusted* [d] | 2.44 | 1.07 | 1.04 | 5.75 | 0.04 |

**Notes:** The *low-risk* group is the reference; HR = hazard ratio; SE = standard error; CI = confidence interval; *p* = significance value.

a *Demographic and genetic variables*: age; sex; 10 principal components (PCs); C-reactive Protein (CRP) polygenic score (PGS); White Blood Cell Counts (WBCC) PGS; Insulin Growth Factor-1 (IGF-1) PGS; Anxiety PGS; Depression PGS; Schizophrenia PGS; Insomnia PGS; Pain PGS.

b All variables: age; sex; 10 PCs; CRP PGS; WBCC PGS; IGF-1 PGS; PGS; Anxiety PGS; Depression PGS; Schizophrenia PGS; Insomnia PGS; Pain PGS; education; wealth; smoking status; alcohol consumption; mobility.

c Additional variables: medication, physical activity; BMI.

d Additional variables: health (i.e., chronic lung disease; coronary heart disease; abnormal heart rhythm; heart murmur; congestive heart failure; angina; hypertension; diabetes; cancer; Parkinson's; Alzheimer's; dementia; asthma; arthritis; osteoporosis; psychiatric disorder).

***Table S9g.*** **Longitudinal associations between C-reactive protein and hospitalisation for sleep disorders (*n*=4,276)**

| **Adjustments** | **CRP** | | | | |
|---|---|---|---|---|---|
| | **HR** | **SE** | **95% CI** | | ***p*** |
| **Sleep Disorders** | | | | | |
| Model 1: *Unadjusted* | 1.92 | 0.37 | 1.31 | 2.81 | 0.001 |
| Model 2: *Model 1 + demographics & genetics* [a] | 2.06 | 0.42 | 1.39 | 3.06 | <0.001 |
| Model 3: *Model 2 + Fully Adjusted* [b] | 2.05 | 0.42 | 1.37 | 3.07 | 0.001 |
| Model 2a: *Demographics* | 2.11 | 0.42 | 1.43 | 3.11 | <0.001 |
| Model 2b: *Genetics* | 1.88 | 0.38 | 1.27 | 2.78 | 0.002 |
| Model 3a: Model 3 + *Medication* | 2.05 | 0.42 | 1.37 | 3.07 | 0.001 |
| Model 3b: Model 3 + *Physical Activity* | 1.90 | 0.40 | 1.26 | 2.86 | 0.002 |
| Model 3c: Model 3 + *Body Mass Index* | 1.81 | 0.39 | 1.18 | 2.77 | 0.006 |
| Model 3d: Model 3 + *Fully Adjusted* [c] | 1.68 | 0.37 | 1.09 | 2.58 | 0.019 |
| Model 4a: Model 2 + *Health* | 2.06 | 0.42 | 1.39 | 3.06 | <0.001 |
| Model 4b: Model 3 + *Health* | 2.04 | 0.42 | 1.36 | 3.07 | 0.001 |
| Model 4c: Model 3 + *Fully Adjusted* [d] | 1.68 | 0.37 | 1.09 | 2.58 | 0.019 |

**Notes:** The *low-risk* group is the reference; HR = hazard ratio; SE = standard error; CI = confidence interval; *p* = significance value.

a *Demographic and genetic variables*: age; sex; 10 principal components (PCs); C-reactive Protein (CRP) polygenic score (PGS); White Blood Cell Counts (WBCC) PGS; Insulin Growth Factor-1 (IGF-1) PGS; Anxiety PGS; Depression PGS; Schizophrenia PGS; Insomnia PGS; Pain PGS.

b All variables: age; sex; 10 PCs; CRP PGS; WBCC PGS; IGF-1 PGS; PGS; Anxiety PGS; Depression PGS; Schizophrenia PGS; Insomnia PGS; Pain PGS; education; wealth; smoking status; alcohol consumption; mobility.

c Additional variables: medication, physical activity; BMI.

d Additional variables: health (i.e., chronic lung disease; coronary heart disease; abnormal heart rhythm; heart murmur; congestive heart failure; angina; hypertension; diabetes; cancer; Parkinson's; Alzheimer's; dementia; asthma; arthritis; osteoporosis; psychiatric disorder).

***Table S9h.*** **Longitudinal associations between fibrinogen and hospitalisation for sleep disorders (*n*=4,276)**

| **Adjustments** | **Fb** | | | | |
|---|---|---|---|---|---|
| | **HR** | **SE** | **95% CI** | | ***p*** |
| **Sleep Disorders** | | | | | |
| Model 1: *Unadjusted* | 1.64 | 0.39 | 1.03 | 2.62 | 0.037 |
| Model 2: *Model 1 + demographics & genetics* [a] | 1.80 | 0.41 | 1.15 | 2.82 | 0.010 |
| Model 3: *Model 2 + Fully Adjusted* [b] | 1.80 | 0.43 | 1.13 | 2.86 | 0.013 |
| Model 2a: *Demographics* | 1.81 | 0.42 | 1.16 | 2.84 | 0.009 |
| Model 2b: *Genetics* | 1.60 | 0.38 | 1.00 | 2.55 | 0.048 |
| Model 3a: Model 3 + *Medication* | 1.80 | 0.43 | 1.13 | 2.86 | 0.014 |
| Model 3b: Model 3 + *Physical Activity* | 1.68 | 0.41 | 1.04 | 2.70 | 0.032 |
| Model 3c: Model 3 + *Body Mass Index* | 1.67 | 0.41 | 1.02 | 2.71 | 0.040 |
| Model 3d: Model 3 + *Fully Adjusted* [c] | 1.58 | 0.40 | 0.96 | 2.58 | 0.070 |
| Model 4a: Model 2 + *Health* | 1.77 | 0.41 | 1.13 | 2.78 | 0.013 |
| Model 4b: Model 3 + *Health* | 1.79 | 0.43 | 1.12 | 2.85 | 0.014 |
| Model 4c: Model 3 + *Fully Adjusted* [d] | 1.57 | 0.40 | 0.96 | 2.58 | 0.071 |

**Notes:** The *low-risk* group is the reference; HR = hazard ratio; SE = standard error; CI = confidence interval; *p* = significance value.

a *Demographic and genetic variables*: age; sex; 10 principal components (PCs); C-reactive Protein (CRP) polygenic score (PGS); White Blood Cell Counts (WBCC) PGS; Insulin Growth Factor-1 (IGF-1) PGS; Anxiety PGS; Depression PGS; Schizophrenia PGS; Insomnia PGS; Pain PGS.

b All variables: age; sex; 10 PCs; CRP PGS; WBCC PGS; IGF-1 PGS; PGS; Anxiety PGS; Depression PGS; Schizophrenia PGS; Insomnia PGS; Pain PGS; education; wealth; smoking status; alcohol consumption; mobility.

c Additional variables: medication, physical activity; BMI.

d Additional variables: health (i.e., chronic lung disease; coronary heart disease; abnormal heart rhythm; heart murmur; congestive heart failure; angina; hypertension; diabetes; cancer; Parkinson's; Alzheimer's; dementia; asthma; arthritis; osteoporosis; psychiatric disorder).

***Table S9i.*** **Longitudinal associations between white blood cell counts and hospitalisation for sleep disorders (*n*=4,276)**

| **Adjustments** | **WBCC** | | | | |
|---|---|---|---|---|---|
| | **HR** | **SE** | **95% CI** | | ***p*** |
| **Sleep Disorders** | | | | | |
| Model 1: *Unadjusted* | 3.77 | 2.12 | 1.25 | 11.36 | 0.018 |
| Model 2: *Model 1 + demographics & genetics* [a] | 1.80 | 0.41 | 1.15 | 2.82 | 0.010 |
| Model 3: *Model 2 + Fully Adjusted* [b] | 1.80 | 0.43 | 1.13 | 2.86 | 0.013 |
| Model 2a: *Demographics* | 1.81 | 0.42 | 1.16 | 2.84 | 0.009 |
| Model 2b: *Genetics* | 1.60 | 0.38 | 1.00 | 2.55 | 0.048 |
| Model 3a: Model 3 + *Medication* | 1.80 | 0.43 | 1.13 | 2.86 | 0.014 |
| Model 3b: Model 3 + *Physical Activity* | 1.68 | 0.41 | 1.04 | 2.70 | 0.032 |
| Model 3c: Model 3 + *Body Mass Index* | 1.67 | 0.41 | 1.02 | 2.71 | 0.040 |
| Model 3d: Model 3 + *Fully Adjusted* [c] | 1.58 | 0.40 | 0.96 | 2.58 | 0.070 |
| Model 4a: Model 2 + *Health* | 1.77 | 0.41 | 1.13 | 2.78 | 0.013 |
| Model 4b: Model 3 + *Health* | 1.79 | 0.43 | 1.12 | 2.85 | 0.014 |
| Model 4c: Model 3 + *Fully Adjusted* [d] | 1.57 | 0.40 | 0.96 | 2.58 | 0.071 |

**Notes:** The *low-risk* group is the reference; HR = hazard ratio; SE = standard error; CI = confidence interval; *p* = significance value.

a *Demographic and genetic variables*: age; sex; 10 principal components (PCs); C-reactive Protein (CRP) polygenic score (PGS); White Blood Cell Counts (WBCC) PGS; Insulin Growth Factor-1 (IGF-1) PGS; Anxiety PGS; Depression PGS; Schizophrenia PGS; Insomnia PGS; Pain PGS.

b All variables: age; sex; 10 PCs; CRP PGS; WBCC PGS; IGF-1 PGS; PGS; Anxiety PGS; Depression PGS; Schizophrenia PGS; Insomnia PGS; Pain PGS; education; wealth; smoking status; alcohol consumption; mobility.

c Additional variables: medication, physical activity; BMI.

d Additional variables: health (i.e., chronic lung disease; coronary heart disease; abnormal heart rhythm; heart murmur; congestive heart failure; angina; hypertension; diabetes; cancer; Parkinson's; Alzheimer's; dementia; asthma; arthritis; osteoporosis; psychiatric disorder).

***Table S9j.*** **Longitudinal associations between insulin growth factor-1 and hospitalisation for sleep disorders (*n*=4,276)**

| **Adjustments** | **IGF-1** | | | | |
|---|---|---|---|---|---|
| | **HR** | **SE** | **95% CI** | | ***p*** |
| **Sleep Disorders** | | | | | |
| Model 1: *Unadjusted* | 1.09 | 0.48 | 0.46 | 2.57 | 0.854 |
| Model 2: *Model 1 + demographics & genetics* [a] | 0.79 | 0.36 | 0.33 | 1.91 | 0.606 |
| Model 3: *Model 2 + Fully Adjusted* [b] | 0.84 | 0.38 | 0.35 | 2.02 | 0.700 |
| Model 2a: *Demographics* | 0.75 | 0.34 | 0.31 | 1.80 | 0.513 |
| Model 2b: *Genetics* | 1.13 | 0.50 | 0.48 | 2.67 | 0.786 |
| Model 3a: Model 3 + *Medication* | 0.84 | 0.38 | 0.35 | 2.02 | 0.700 |
| Model 3b: Model 3 + *Physical Activity* | 0.87 | 0.38 | 0.37 | 2.07 | 0.751 |
| Model 3c: Model 3 + *Body Mass Index* | 0.88 | 0.39 | 0.37 | 2.08 | 0.761 |
| Model 3d: Model 3 + *Fully Adjusted* [c] | 0.92 | 0.40 | 0.39 | 2.16 | 0.845 |
| Model 4a: Model 2 + *Health* | 0.81 | 0.36 | 0.34 | 1.95 | 0.640 |
| Model 4b: Model 3 + *Health* | 0.85 | 0.38 | 0.35 | 2.03 | 0.712 |
| Model 4c: Model 3 + *Fully Adjusted* [d] | 0.92 | 0.40 | 0.39 | 2.17 | 0.848 |

**Notes:** The *low-risk* group is the reference; HR = hazard ratio; SE = standard error; CI = confidence interval; *p* = significance value.

a *Demographic and genetic variables*: age; sex; 10 principal components (PCs); C-reactive Protein (CRP) polygenic score (PGS); White Blood Cell Counts (WBCC) PGS; Insulin Growth Factor-1 (IGF-1) PGS; Anxiety PGS; Depression PGS; Schizophrenia PGS; Insomnia PGS; Pain PGS.

b All variables: age; sex; 10 PCs; CRP PGS; WBCC PGS; IGF-1 PGS; PGS; Anxiety PGS; Depression PGS; Schizophrenia PGS; Insomnia PGS; Pain PGS; education; wealth; smoking status; alcohol consumption; mobility.

c Additional variables: medication, physical activity; BMI.

d Additional variables: health (i.e., chronic lung disease; coronary heart disease; abnormal heart rhythm; heart murmur; congestive heart failure; angina; hypertension; diabetes; cancer; Parkinson's; Alzheimer's; dementia; asthma; arthritis; osteoporosis; psychiatric disorder).

***Table S9k.*** **Longitudinal associations between immune and neuroendocrine profiles and hospitalisation for diseases of the blood and blood-forming organs and certain disorders involving the immune mechanism (*n*=4,139)**

| Adjustments | Immune and Neuroendocrine Profiles | | | | |
|---|---|---|---|---|---|
| | **HR** | **SE** | **95% CI** | | ***p*** |
| ***Moderate-risk*** **Profile \| Blood Disorders** | | | | | |
| Model 1: *Unadjusted* | 1.38 | 0.15 | 1.13 | 1.70 | 0.002 |
| Model 2: *Model 1 + demographics & genetics* [a] | 1.19 | 0.13 | 0.97 | 1.47 | 0.099 |
| Model 3: *Model 2 + Fully Adjusted* [b] | 1.10 | 0.12 | 0.89 | 1.36 | 0.388 |
| Model 2a: *Demographics* | 1.21 | 0.13 | 0.99 | 1.49 | 0.070 |
| Model 2b: *Genetics* | 1.37 | 0.14 | 1.12 | 1.69 | 0.003 |
| Model 3a: Model 3 + *Medication* | 1.10 | 0.12 | 0.89 | 1.35 | 0.398 |
| Model 3b: Model 3 + *Physical Activity* | 1.07 | 0.12 | 0.87 | 1.32 | 0.532 |
| Model 3c: Model 3 + *Body Mass Index* | 1.09 | 0.12 | 0.87 | 1.35 | 0.458 |
| Model 3d: Model 3 + *Fully Adjusted* [c] | 1.06 | 0.12 | 0.86 | 1.32 | 0.585 |
| Model 4a: Model 2 + *Health* | 1.19 | 0.13 | 0.97 | 1.47 | 0.099 |
| Model 4b: Model 3 + *Health* | 1.09 | 0.12 | 0.88 | 1.34 | 0.432 |
| Model 4c: Model 3 + *Fully Adjusted* [d] | 1.06 | 0.12 | 0.85 | 1.31 | 0.623 |
| ***High-risk*** **Profile \| Blood Disorders** | | | | | |
| Model 1: *Unadjusted* | 2.21 | 0.29 | 1.71 | 2.85 | <0.001 |
| Model 2: *Model 1 + demographics & genetics* [a] | 1.92 | 0.26 | 1.48 | 2.49 | <0.001 |
| Model 3: *Model 2 + Fully Adjusted* [b] | 1.68 | 0.23 | 1.28 | 2.19 | <0.001 |
| Model 2a: *Demographics* | 1.96 | 0.26 | 1.51 | 2.54 | <0.001 |
| Model 2b: *Genetics* | 2.16 | 0.29 | 1.67 | 2.80 | <0.001 |
| Model 3a: Model 3 + *Medication* | 1.68 | 0.23 | 1.29 | 2.19 | <0.001 |
| Model 3b: Model 3 + *Physical Activity* | 1.59 | 0.22 | 1.21 | 2.07 | 0.001 |
| Model 3c: Model 3 + *Body Mass Index* | 1.65 | 0.23 | 1.26 | 2.17 | <0.001 |
| Model 3d: Model 3 + *Fully Adjusted* [c] | 1.58 | 0.22 | 1.20 | 2.08 | 0.001 |
| Model 4a: Model 2 + *Health* | 1.92 | 0.26 | 1.48 | 2.49 | <0.001 |
| Model 4b: Model 3 + *Health* | 1.67 | 0.23 | 1.28 | 2.18 | <0.001 |
| Model 4c: Model 3 + *Fully Adjusted* [d] | 1.58 | 0.22 | 1.20 | 2.08 | 0.001 |

**Notes:** The *low-risk* group is the reference; HR = hazard ratio; SE = standard error; CI = confidence interval; *p* = significance value.

a *Demographic and genetic variables*: age; sex; 10 principal components (PCs); C-reactive Protein (CRP) polygenic score (PGS); White Blood Cell Counts (WBCC) PGS; Insulin Growth Factor-1 (IGF-1) PGS; Anxiety PGS; Depression PGS; Schizophrenia PGS; Insomnia PGS; Pain PGS.

b All variables: age; sex; 10 PCs; CRP PGS; WBCC PGS; IGF-1 PGS; PGS; Anxiety PGS; Depression PGS; Schizophrenia PGS; Insomnia PGS; Pain PGS; education; wealth; smoking status; alcohol consumption; mobility.

c Additional variables: medication, physical activity; BMI.

d Additional variables: health (i.e., chronic lung disease; coronary heart disease; abnormal heart rhythm; heart murmur; congestive heart failure; angina; hypertension; diabetes; cancer; Parkinson's; Alzheimer's; dementia; asthma; arthritis; osteoporosis; psychiatric disorder).

***Table S91.*** **Longitudinal associations between C-reactive protein and hospitalisation for diseases of the blood and blood-forming organs and certain disorders involving the immune mechanism (*n*=4,139)**

| Adjustments | CRP | | | | |
|---|---|---|---|---|---|
| | HR | SE | 95% CI | | *p* |
| **Blood Disorders** | | | | | |
| Model 1: *Unadjusted* | 1.49 | 0.10 | 1.31 | 1.69 | <0.001 |
| Model 2: *Model 1 + demographics & genetics* [a] | 1.37 | 0.09 | 1.19 | 1.56 | <0.001 |
| Model 3: *Model 2 + Fully Adjusted* [b] | 1.27 | 0.09 | 1.11 | 1.46 | 0.001 |
| Model 2a: *Demographics* | 1.39 | 0.10 | 1.21 | 1.58 | <0.001 |
| Model 2b: *Genetics* | 1.47 | 0.10 | 1.29 | 1.67 | <0.001 |
| Model 3a: Model 3 + *Medication* | 1.28 | 0.09 | 1.11 | 1.46 | 0.001 |
| Model 3b: Model 3 + *Physical Activity* | 1.24 | 0.09 | 1.08 | 1.42 | 0.003 |
| Model 3c: Model 3 + *Body Mass Index* | 1.27 | 0.09 | 1.10 | 1.47 | 0.001 |
| Model 3d: Model 3 + *Fully Adjusted* [c] | 1.24 | 0.09 | 1.07 | 1.43 | 0.004 |
| Model 4a: Model 2 + *Health* | 1.37 | 0.09 | 1.19 | 1.56 | <0.001 |
| Model 4b: Model 3 + *Health* | 1.27 | 0.09 | 1.11 | 1.46 | 0.001 |
| Model 4c: Model 3 + *Fully Adjusted* [d] | 1.24 | 0.09 | 1.07 | 1.43 | 0.004 |

**Notes:** The *low-risk* group is the reference; HR = hazard ratio; SE = standard error; CI = confidence interval; *p* = significance value.

a *Demographic and genetic variables*: age; sex; 10 principal components (PCs); C-reactive Protein (CRP) polygenic score (PGS); White Blood Cell Counts (WBCC) PGS; Insulin Growth Factor-1 (IGF-1) PGS; Anxiety PGS; Depression PGS; Schizophrenia PGS; Insomnia PGS; Pain PGS.

b All variables: age; sex; 10 PCs; CRP PGS; WBCC PGS; IGF-1 PGS; PGS; Anxiety PGS; Depression PGS; Schizophrenia PGS; Insomnia PGS; Pain PGS; education; wealth; smoking status; alcohol consumption; mobility.

c Additional variables: medication, physical activity; BMI.

d Additional variables: health (i.e., chronic lung disease; coronary heart disease; abnormal heart rhythm; heart murmur; congestive heart failure; angina; hypertension; diabetes; cancer; Parkinson's; Alzheimer's; dementia; asthma; arthritis; osteoporosis; psychiatric disorder).

**_Table S9m._** **Longitudinal associations between fibrinogen and hospitalisation for diseases of the blood and blood-forming organs and certain disorders involving the immune mechanism (*n*=4,139)**

| **Adjustments** | **Fb** | | | | |
|---|---|---|---|---|---|
| | **HR** | **SE** | **95% CI** | | ***p*** |
| **Blood Disorders** | | | | | |
| Model 1: *Unadjusted* | 1.49 | 0.12 | 1.28 | 1.75 | <0.001 |
| Model 2: *Model 1 + demographics & genetics* [a] | 1.38 | 0.12 | 1.17 | 1.63 | <0.001 |
| Model 3: *Model 2 + Fully Adjusted* [b] | 1.26 | 0.11 | 1.07 | 1.49 | 0.007 |
| Model 2a: *Demographics* | 1.38 | 0.12 | 1.18 | 1.63 | <0.001 |
| Model 2b: *Genetics* | 1.48 | 0.12 | 1.26 | 1.73 | <0.001 |
| Model 3a: Model 3 + *Medication* | 1.26 | 0.11 | 1.07 | 1.49 | 0.007 |
| Model 3b: Model 3 + *Physical Activity* | 1.24 | 0.11 | 1.05 | 1.46 | 0.013 |
| Model 3c: Model 3 + *Body Mass Index* | 1.25 | 0.11 | 1.06 | 1.48 | 0.010 |
| Model 3d: Model 3 + *Fully Adjusted* [c] | 1.23 | 0.11 | 1.04 | 1.46 | 0.016 |
| Model 4a: Model 2 + *Health* | 1.37 | 0.12 | 1.17 | 1.62 | <0.001 |
| Model 4b: Model 3 + *Health* | 1.26 | 0.11 | 1.07 | 1.49 | 0.007 |
| Model 4c: Model 3 + *Fully Adjusted* [d] | 1.23 | 0.11 | 1.04 | 1.46 | 0.015 |

**Notes:** The *low-risk* group is the reference; HR = hazard ratio; SE = standard error; CI = confidence interval; *p* = significance value.

a *Demographic and genetic variables*: age; sex; 10 principal components (PCs); C-reactive Protein (CRP) polygenic score (PGS); White Blood Cell Counts (WBCC) PGS; Insulin Growth Factor-1 (IGF-1) PGS; Anxiety PGS; Depression PGS; Schizophrenia PGS; Insomnia PGS; Pain PGS.

b All variables: age; sex; 10 PCs; CRP PGS; WBCC PGS; IGF-1 PGS; PGS; Anxiety PGS; Depression PGS; Schizophrenia PGS; Insomnia PGS; Pain PGS; education; wealth; smoking status; alcohol consumption; mobility.

c Additional variables: medication, physical activity; BMI.

d Additional variables: health (i.e., chronic lung disease; coronary heart disease; abnormal heart rhythm; heart murmur; congestive heart failure; angina; hypertension; diabetes; cancer; Parkinson's; Alzheimer's; dementia; asthma; arthritis; osteoporosis; psychiatric disorder).

***Table S9n.*** **Longitudinal associations between white blood cell counts and hospitalisation for diseases of the blood and blood-forming organs and certain disorders involving the immune mechanism (*n*=4,139)**

| **Adjustments** | **WBCC** | | | | |
|---|---|---|---|---|---|
| | **HR** | **SE** | **95% CI** | | ***p*** |
| **Blood Disorders** | | | | | |
| Model 1: *Unadjusted* | 2.04 | 0.39 | 1.40 | 2.97 | <0.001 |
| Model 2: *Model 1 + demographics & genetics* [a] | 1.87 | 0.37 | 1.27 | 2.77 | 0.002 |
| Model 3: *Model 2 + Fully Adjusted* [b] | 1.50 | 0.31 | 1.00 | 2.25 | 0.052 |
| Model 2a: *Demographics* | 1.77 | 0.35 | 1.21 | 2.61 | 0.004 |
| Model 2b: *Genetics* | 2.11 | 0.41 | 1.44 | 3.07 | <0.001 |
| Model 3a: Model 3 + *Medication* | 1.50 | 0.31 | 1.00 | 2.25 | 0.051 |
| Model 3b: Model 3 + *Physical Activity* | 1.45 | 0.30 | 0.97 | 2.18 | 0.074 |
| Model 3c: Model 3 + *Body Mass Index* | 1.47 | 0.31 | 0.97 | 2.21 | 0.069 |
| Model 3d: Model 3 + *Fully Adjusted* [c] | 1.43 | 0.30 | 0.95 | 2.15 | 0.088 |
| Model 4a: Model 2 + *Health* | 1.82 | 0.36 | 1.23 | 2.68 | 0.003 |
| Model 4b: Model 3 + *Health* | 1.48 | 0.31 | 0.98 | 2.22 | 0.060 |
| Model 4c: Model 3 + *Fully Adjusted* [d] | 1.42 | 0.30 | 0.94 | 2.13 | 0.097 |

**Notes:** The *low-risk* group is the reference; HR = hazard ratio; SE = standard error; CI = confidence interval; *p* = significance value.

a *Demographic and genetic variables*: age; sex; 10 principal components (PCs); C-reactive Protein (CRP) polygenic score (PGS); White Blood Cell Counts (WBCC) PGS; Insulin Growth Factor-1 (IGF-1) PGS; Anxiety PGS; Depression PGS; Schizophrenia PGS; Insomnia PGS; Pain PGS.

b All variables: age; sex; 10 PCs; CRP PGS; WBCC PGS; IGF-1 PGS; PGS; Anxiety PGS; Depression PGS; Schizophrenia PGS; Insomnia PGS; Pain PGS; education; wealth; smoking status; alcohol consumption; mobility.

c Additional variables: medication, physical activity; BMI.

d Additional variables: health (i.e., chronic lung disease; coronary heart disease; abnormal heart rhythm; heart murmur; congestive heart failure; angina; hypertension; diabetes; cancer; Parkinson's; Alzheimer's; dementia; asthma; arthritis; osteoporosis; psychiatric disorder).

***Table S9o.*** **Longitudinal associations between insulin growth factor-1 and hospitalisation for diseases of the blood and blood-forming organs and certain disorders involving the immune mechanism (*n*=4,139)**

| **Adjustments** | **IGF-1** | | | | |
|---|---|---|---|---|---|
| | **HR** | **SE** | **95% CI** | | ***p*** |
| **Blood Disorders** | | | | | |
| Model 1: *Unadjusted* | 0.61 | 0.09 | 0.46 | 0.81 | 0.001 |
| Model 2: *Model 1 + demographics & genetics* [a] | 0.86 | 0.13 | 0.64 | 1.15 | 0.304 |
| Model 3: *Model 2 + Fully Adjusted* [b] | 0.87 | 0.13 | 0.66 | 1.16 | 0.349 |
| Model 2a: *Demographics* | 0.85 | 0.12 | 0.64 | 1.13 | 0.255 |
| Model 2b: *Genetics* | 0.62 | 0.09 | 0.47 | 0.82 | 0.001 |
| Model 3a: Model 3 + *Medication* | 0.87 | 0.13 | 0.66 | 1.16 | 0.342 |
| Model 3b: Model 3 + *Physical Activity* | 0.89 | 0.13 | 0.67 | 1.17 | 0.392 |
| Model 3c: Model 3 + *Body Mass Index* | 0.88 | 0.13 | 0.66 | 1.16 | 0.360 |
| Model 3d: Model 3 + *Fully Adjusted* [c] | 0.89 | 0.13 | 0.67 | 1.17 | 0.394 |
| Model 4a: Model 2 + *Health* | 0.86 | 0.13 | 0.65 | 1.15 | 0.309 |
| Model 4b: Model 3 + *Health* | 0.87 | 0.13 | 0.66 | 1.16 | 0.350 |
| Model 4c: Model 3 + *Fully Adjusted* [d] | 0.89 | 0.13 | 0.67 | 1.17 | 0.396 |

**Notes:** The *low-risk* group is the reference; HR = hazard ratio; SE = standard error; CI = confidence interval; *p* = significance value.

a *Demographic and genetic variables*: age; sex; 10 principal components (PCs); C-reactive Protein (CRP) polygenic score (PGS); White Blood Cell Counts (WBCC) PGS; Insulin Growth Factor-1 (IGF-1) PGS; Anxiety PGS; Depression PGS; Schizophrenia PGS; Insomnia PGS; Pain PGS.

b All variables: age; sex; 10 PCs; CRP PGS; WBCC PGS; IGF-1 PGS; PGS; Anxiety PGS; Depression PGS; Schizophrenia PGS; Insomnia PGS; Pain PGS; education; wealth; smoking status; alcohol consumption; mobility.

c Additional variables: medication, physical activity; BMI.

d Additional variables: health (i.e., chronic lung disease; coronary heart disease; abnormal heart rhythm; heart murmur; congestive heart failure; angina; hypertension; diabetes; cancer; Parkinson's; Alzheimer's; dementia; asthma; arthritis; osteoporosis; psychiatric disorder).

***Table S9p.*** **Longitudinal associations between immune and neuroendocrine profiles and hospitalisation for diseases of the circulatory system (*n*=3,163)**

| Adjustments | Immune and Neuroendocrine Profiles | | | | |
|---|---|---|---|---|---|
| | HR | SE | 95% CI | | *p* |
| ***Moderate-risk* Profile \| Circulatory Disorders** | | | | | |
| Model 1: *Unadjusted* | 1.49 | 0.09 | 1.33 | 1.67 | <0.001 |
| Model 2: *Model 1 + demographics & genetics* [a] | 1.34 | 0.08 | 1.19 | 1.50 | <0.001 |
| Model 3: *Model 2 + Fully Adjusted* [b] | 1.26 | 0.08 | 1.12 | 1.42 | <0.001 |
| Model 2a: *Demographics* | 1.35 | 0.08 | 1.20 | 1.51 | <0.001 |
| Model 2b: *Genetics* | 1.48 | 0.09 | 1.32 | 1.66 | <0.001 |
| Model 3a: Model 3 + *Medication* | 1.27 | 0.08 | 1.13 | 1.42 | <0.001 |
| Model 3b: Model 3 + *Physical Activity* | 1.25 | 0.07 | 1.11 | 1.40 | <0.001 |
| Model 3c: Model 3 + *Body Mass Index* | 1.19 | 0.07 | 1.05 | 1.34 | 0.005 |
| Model 3d: Model 3 + *Fully Adjusted* [c] | 1.18 | 0.07 | 1.05 | 1.34 | 0.006 |
| Model 4a: Model 2 + *Health* | 1.49 | 0.09 | 1.33 | 1.67 | <0.001 |
| Model 4b: Model 3 + *Health* | 1.25 | 0.07 | 1.12 | 1.41 | <0.001 |
| Model 4c: Model 3 + *Fully Adjusted* [d] | 1.18 | 0.07 | 1.04 | 1.33 | 0.009 |
| ***High-risk* Profile \| Circulatory Disorders** | | | | | |
| Model 1: *Unadjusted* | 1.72 | 0.14 | 1.46 | 2.03 | <0.001 |
| Model 2: *Model 1 + demographics & genetics* [a] | 1.51 | 0.13 | 1.27 | 1.78 | <0.001 |
| Model 3: *Model 2 + Fully Adjusted* [b] | 1.37 | 0.12 | 1.16 | 1.63 | <0.001 |
| Model 2a: *Demographics* | 1.53 | 0.13 | 1.30 | 1.81 | <0.001 |
| Model 2b: *Genetics* | 1.69 | 0.14 | 1.43 | 1.99 | <0.001 |
| Model 3a: Model 3 + *Medication* | 1.37 | 0.12 | 1.15 | 1.62 | <0.001 |
| Model 3b: Model 3 + *Physical Activity* | 1.35 | 0.12 | 1.14 | 1.60 | 0.001 |
| Model 3c: Model 3 + *Body Mass Index* | 1.28 | 0.11 | 1.07 | 1.52 | 0.006 |
| Model 3d: Model 3 + *Fully Adjusted* [c] | 1.25 | 0.11 | 1.05 | 1.49 | 0.011 |
| Model 4a: Model 2 + *Health* | 1.72 | 0.14 | 1.46 | 2.03 | <0.001 |
| Model 4b: Model 3 + *Health* | 1.36 | 0.12 | 1.15 | 1.61 | <0.001 |
| Model 4c: Model 3 + *Fully Adjusted* [d] | 1.25 | 0.11 | 1.05 | 1.48 | 0.014 |

**Notes:** The *low-risk* group is the reference; HR = hazard ratio; SE = standard error; CI = confidence interval; *p* = significance value.

a *Demographic and genetic variables*: age; sex; 10 principal components (PCs); C-reactive Protein (CRP) polygenic score (PGS); White Blood Cell Counts (WBCC) PGS; Insulin Growth Factor-1 (IGF-1) PGS; Anxiety PGS; Depression PGS; Schizophrenia PGS; Insomnia PGS; Pain PGS.

b All variables: age; sex; 10 PCs; CRP PGS; WBCC PGS; IGF-1 PGS; PGS; Anxiety PGS; Depression PGS; Schizophrenia PGS; Insomnia PGS; Pain PGS; education; wealth; smoking status; alcohol consumption; mobility.

c Additional variables: medication, physical activity; BMI.

d Additional variables: health (i.e., chronic lung disease; coronary heart disease; abnormal heart rhythm; heart murmur; congestive heart failure; angina; hypertension; diabetes; cancer; Parkinson's; Alzheimer's; dementia; asthma; arthritis; osteoporosis; psychiatric disorder).

***Table S9q.*** **Longitudinal associations between C-reactive protein and hospitalisation for diseases of the circulatory system (*n*=3,163)**

| Adjustments | CRP | | | | |
|---|---|---|---|---|---|
| | HR | SE | 95% CI | | *p* |
| **Circulatory Disorders** | | | | | |
| Model 1: *Unadjusted* | 1.41 | 0.05 | 1.31 | 1.52 | <0.001 |
| Model 2: *Model 1 + demographics & genetics* [a] | 1.31 | 0.05 | 1.21 | 1.42 | <0.001 |
| Model 3: *Model 2 + Fully Adjusted* [b] | 1.25 | 0.05 | 1.15 | 1.36 | <0.001 |
| Model 2a: *Demographics* | 1.32 | 0.05 | 1.22 | 1.43 | <0.001 |
| Model 2b: *Genetics* | 1.41 | 0.05 | 1.30 | 1.52 | <0.001 |
| Model 3a: Model 3 + *Medication* | 1.25 | 0.05 | 1.15 | 1.36 | <0.001 |
| Model 3b: Model 3 + *Physical Activity* | 1.24 | 0.05 | 1.14 | 1.34 | <0.001 |
| Model 3c: Model 3 + *Body Mass Index* | 1.20 | 0.05 | 1.10 | 1.30 | <0.001 |
| Model 3d: Model 3 + *Fully Adjusted* [c] | 1.19 | 0.05 | 1.09 | 1.29 | <0.001 |
| Model 4a: Model 2 + *Health* | 1.31 | 0.05 | 1.21 | 1.42 | <0.001 |
| Model 4b: Model 3 + *Health* | 1.24 | 0.05 | 1.15 | 1.35 | <0.001 |
| Model 4c: Model 3 + *Fully Adjusted* [d] | 1.18 | 0.05 | 1.08 | 1.28 | <0.001 |

**Notes:** The *low-risk* group is the reference; HR = hazard ratio; SE = standard error; CI = confidence interval; *p* = significance value.

a *Demographic and genetic variables*: age; sex; 10 principal components (PCs); C-reactive Protein (CRP) polygenic score (PGS); White Blood Cell Counts (WBCC) PGS; Insulin Growth Factor-1 (IGF-1) PGS; Anxiety PGS; Depression PGS; Schizophrenia PGS; Insomnia PGS; Pain PGS.

b All variables: age; sex; 10 PCs; CRP PGS; WBCC PGS; IGF-1 PGS; PGS; Anxiety PGS; Depression PGS; Schizophrenia PGS; Insomnia PGS; Pain PGS; education; wealth; smoking status; alcohol consumption; mobility.

c Additional variables: medication, physical activity; BMI.

d Additional variables: health (i.e., chronic lung disease; coronary heart disease; abnormal heart rhythm; heart murmur; congestive heart failure; angina; hypertension; diabetes; cancer; Parkinson's; Alzheimer's; dementia; asthma; arthritis; osteoporosis; psychiatric disorder).

***Table S9r.*** **Longitudinal associations between fibrinogen and hospitalisation for diseases of the circulatory system (*n*=3,163)**

| Adjustments | Fb | | | | |
|---|---|---|---|---|---|
| | **HR** | **SE** | **95% CI** | | ***p*** |
| **Circulatory Disorders** | | | | | |
| Model 1: *Unadjusted* | 1.42 | 0.07 | 1.29 | 1.57 | <0.001 |
| Model 2: *Model 1 + demographics & genetics* [a] | 1.29 | 0.07 | 1.17 | 1.43 | <0.001 |
| Model 3: *Model 2 + Fully Adjusted* [b] | 1.21 | 0.06 | 1.10 | 1.34 | <0.001 |
| Model 2a: *Demographics* | 1.31 | 0.07 | 1.18 | 1.44 | <0.001 |
| Model 2b: *Genetics* | 1.40 | 0.07 | 1.28 | 1.55 | <0.001 |
| Model 3a: Model 3 + *Medication* | 1.21 | 0.06 | 1.10 | 1.34 | <0.001 |
| Model 3b: Model 3 + *Physical Activity* | 1.20 | 0.06 | 1.08 | 1.33 | <0.001 |
| Model 3c: Model 3 + *Body Mass Index* | 1.17 | 0.06 | 1.05 | 1.29 | 0.003 |
| Model 3d: Model 3 + *Fully Adjusted* [c] | 1.16 | 0.06 | 1.05 | 1.29 | 0.004 |
| Model 4a: Model 2 + *Health* | 1.29 | 0.07 | 1.17 | 1.42 | <0.001 |
| Model 4b: Model 3 + *Health* | 1.21 | 0.06 | 1.10 | 1.34 | <0.001 |
| Model 4c: Model 3 + *Fully Adjusted* [d] | 1.16 | 0.06 | 1.05 | 1.29 | 0.004 |

**Notes:** The *low-risk* group is the reference; HR = hazard ratio; SE = standard error; CI = confidence interval; *p* = significance value.

a *Demographic and genetic variables*: age; sex; 10 principal components (PCs); C-reactive Protein (CRP) polygenic score (PGS); White Blood Cell Counts (WBCC) PGS; Insulin Growth Factor-1 (IGF-1) PGS; Anxiety PGS; Depression PGS; Schizophrenia PGS; Insomnia PGS; Pain PGS.

b All variables: age; sex; 10 PCs; CRP PGS; WBCC PGS; IGF-1 PGS; PGS; Anxiety PGS; Depression PGS; Schizophrenia PGS; Insomnia PGS; Pain PGS; education; wealth; smoking status; alcohol consumption; mobility.

c Additional variables: medication, physical activity; BMI.

d Additional variables: health (i.e., chronic lung disease; coronary heart disease; abnormal heart rhythm; heart murmur; congestive heart failure; angina; hypertension; diabetes; cancer; Parkinson's; Alzheimer's; dementia; asthma; arthritis; osteoporosis; psychiatric disorder).

***Table S9s.*** **Longitudinal associations between white blood cell counts and hospitalisation for diseases of the circulatory system (*n*=3,163)**

| **Adjustments** | **WBCC** | | | | |
|---|---|---|---|---|---|
| | **HR** | **SE** | **95% CI** | | ***p*** |
| **Circulatory Disorders** | | | | | |
| Model 1: *Unadjusted* | 2.06 | 0.22 | 1.67 | 2.54 | <0.001 |
| Model 2: *Model 1 + demographics & genetics* [a] | 1.95 | 0.22 | 1.56 | 2.43 | <0.001 |
| Model 3: *Model 2 + Fully Adjusted* [b] | 1.73 | 0.20 | 1.38 | 2.18 | <0.001 |
| Model 2a: *Demographics* | 1.94 | 0.21 | 1.56 | 2.41 | <0.001 |
| Model 2b: *Genetics* | 2.02 | 0.22 | 1.64 | 2.50 | <0.001 |
| Model 3a: Model 3 + *Medication* | 1.74 | 0.21 | 1.39 | 2.20 | <0.001 |
| Model 3b: Model 3 + *Physical Activity* | 1.69 | 0.20 | 1.34 | 2.13 | <0.001 |
| Model 3c: Model 3 + *Body Mass Index* | 1.61 | 0.19 | 1.28 | 2.03 | <0.001 |
| Model 3d: Model 3 + *Fully Adjusted* [c] | 1.59 | 0.19 | 1.26 | 2.00 | <0.001 |
| Model 4a: Model 2 + *Health* | 1.88 | 0.21 | 1.51 | 2.35 | <0.001 |
| Model 4b: Model 3 + *Health* | 1.70 | 0.20 | 1.35 | 2.14 | <0.001 |
| Model 4c: Model 3 + *Fully Adjusted* [d] | 1.56 | 0.19 | 1.24 | 1.98 | <0.001 |

**Notes:** The *low-risk* group is the reference; HR = hazard ratio; SE = standard error; CI = confidence interval; *p* = significance value.

a *Demographic and genetic variables*: age; sex; 10 principal components (PCs); C-reactive Protein (CRP) polygenic score (PGS); White Blood Cell Counts (WBCC) PGS; Insulin Growth Factor-1 (IGF-1) PGS; Anxiety PGS; Depression PGS; Schizophrenia PGS; Insomnia PGS; Pain PGS.

b All variables: age; sex; 10 PCs; CRP PGS; WBCC PGS; IGF-1 PGS; PGS; Anxiety PGS; Depression PGS; Schizophrenia PGS; Insomnia PGS; Pain PGS; education; wealth; smoking status; alcohol consumption; mobility.

c Additional variables: medication, physical activity; BMI.

d Additional variables: health (i.e., chronic lung disease; coronary heart disease; abnormal heart rhythm; heart murmur; congestive heart failure; angina; hypertension; diabetes; cancer; Parkinson's; Alzheimer's; dementia; asthma; arthritis; osteoporosis; psychiatric disorder).

*Table S9t.* **Longitudinal associations between insulin growth factor-1 and hospitalisation for diseases of the circulatory system (*n*=3,163)**

| Adjustments | IGF-1 | | | | |
|---|---|---|---|---|---|
| | HR | SE | 95% CI | | *p* |
| **Circulatory Disorders** | | | | | |
| Model 1: *Unadjusted* | 0.64 | 0.05 | 0.55 | 0.76 | <0.001 |
| Model 2: *Model 1 + demographics & genetics* [a] | 0.89 | 0.08 | 0.76 | 1.06 | 0.190 |
| Model 3: *Model 2 + Fully Adjusted* [b] | 0.91 | 0.08 | 0.77 | 1.08 | 0.273 |
| Model 2a: *Demographics* | 0.91 | 0.08 | 0.77 | 1.08 | 0.272 |
| Model 2b: *Genetics* | 0.64 | 0.05 | 0.55 | 0.75 | <0.001 |
| Model 3a: Model 3 + *Medication* | 0.92 | 0.08 | 0.78 | 1.08 | 0.298 |
| Model 3b: Model 3 + *Physical Activity* | 0.92 | 0.08 | 0.78 | 1.09 | 0.335 |
| Model 3c: Model 3 + *Body Mass Index* | 0.92 | 0.08 | 0.78 | 1.08 | 0.314 |
| Model 3d: Model 3 + *Fully Adjusted* [c] | 0.94 | 0.08 | 0.80 | 1.10 | 0.426 |
| Model 4a: Model 2 + *Health* | 0.90 | 0.08 | 0.76 | 1.06 | 0.221 |
| Model 4b: Model 3 + *Health* | 0.92 | 0.08 | 0.78 | 1.08 | 0.301 |
| Model 4c: Model 3 + *Fully Adjusted* [d] | 0.94 | 0.08 | 0.80 | 1.11 | 0.453 |

**Notes:** The *low-risk* group is the reference; HR = hazard ratio; SE = standard error; CI = confidence interval; *p* = significance value.

a *Demographic and genetic variables*: age; sex; 10 principal components (PCs); C-reactive Protein (CRP) polygenic score (PGS); White Blood Cell Counts (WBCC) PGS; Insulin Growth Factor-1 (IGF-1) PGS; Anxiety PGS; Depression PGS; Schizophrenia PGS; Insomnia PGS; Pain PGS.

b All variables: age; sex; 10 PCs; CRP PGS; WBCC PGS; IGF-1 PGS; PGS; Anxiety PGS; Depression PGS; Schizophrenia PGS; Insomnia PGS; Pain PGS; education; wealth; smoking status; alcohol consumption; mobility.

c Additional variables: medication, physical activity; BMI.

d Additional variables: health (i.e., chronic lung disease; coronary heart disease; abnormal heart rhythm; heart murmur; congestive heart failure; angina; hypertension; diabetes; cancer; Parkinson's; Alzheimer's; dementia; asthma; arthritis; osteoporosis; psychiatric disorder).

***Table S9u.*** **Longitudinal associations between immune and neuroendocrine profiles and hospitalisation for diseases of the digestive system (*n*=3,188)**

| **Adjustments** | **Immune and Neuroendocrine Profiles** | | | | |
|---|---|---|---|---|---|
| | **HR** | **SE** | **95% CI** | | ***p*** |
| ***Moderate-risk* Profile \| Digestive Disorders** | | | | | |
| Model 1: *Unadjusted* | 1.20 | 0.08 | 1.06 | 1.36 | 0.004 |
| Model 2: *Model 1 + demographics & genetics* [a] | 1.09 | 0.07 | 0.96 | 1.24 | 0.177 |
| Model 3: *Model 2 + Fully Adjusted* [b] | 1.04 | 0.07 | 0.92 | 1.18 | 0.545 |
| Model 2a: *Demographics* | 1.11 | 0.07 | 0.98 | 1.26 | 0.098 |
| Model 2b: *Genetics* | 1.18 | 0.08 | 1.04 | 1.34 | 0.009 |
| Model 3a: Model 3 + *Medication* | 1.04 | 0.07 | 0.91 | 1.18 | 0.574 |
| Model 3b: Model 3 + *Physical Activity* | 1.03 | 0.07 | 0.91 | 1.17 | 0.632 |
| Model 3c: Model 3 + *Body Mass Index* | 1.03 | 0.07 | 0.90 | 1.18 | 0.655 |
| Model 3d: Model 3 + *Fully Adjusted* [c] | 1.02 | 0.07 | 0.90 | 1.17 | 0.750 |
| Model 4a: Model 2 + *Health* | 1.09 | 0.07 | 0.96 | 1.24 | 0.177 |
| Model 4b: Model 3 + *Health* | 1.03 | 0.07 | 0.91 | 1.17 | 0.620 |
| Model 4c: Model 3 + *Fully Adjusted* [d] | 1.02 | 0.07 | 0.89 | 1.16 | 0.810 |
| ***High-risk* Profile \| Digestive Disorders** | | | | | |
| Model 1: *Unadjusted* | 1.12 | 0.11 | 0.93 | 1.36 | 0.236 |
| Model 2: *Model 1 + demographics & genetics* [a] | 1.00 | 0.10 | 0.82 | 1.22 | 0.998 |
| Model 3: *Model 2 + Fully Adjusted* [b] | 0.93 | 0.09 | 0.76 | 1.13 | 0.460 |
| Model 2a: *Demographics* | 1.03 | 0.10 | 0.85 | 1.26 | 0.733 |
| Model 2b: *Genetics* | 1.09 | 0.11 | 0.89 | 1.32 | 0.409 |
| Model 3a: Model 3 + *Medication* | 0.93 | 0.10 | 0.76 | 1.14 | 0.472 |
| Model 3b: Model 3 + *Physical Activity* | 0.91 | 0.09 | 0.75 | 1.12 | 0.376 |
| Model 3c: Model 3 + *Body Mass Index* | 0.92 | 0.10 | 0.75 | 1.13 | 0.407 |
| Model 3d: Model 3 + *Fully Adjusted* [c] | 0.91 | 0.10 | 0.74 | 1.11 | 0.354 |
| Model 4a: Model 2 + *Health* | 1.00 | 0.10 | 0.82 | 1.22 | 0.998 |
| Model 4b: Model 3 + *Health* | 0.92 | 0.09 | 0.75 | 1.12 | 0.404 |
| Model 4c: Model 3 + *Fully Adjusted* [d] | 0.90 | 0.09 | 0.74 | 1.11 | 0.322 |

**Notes:** The *low-risk* group is the reference; HR = hazard ratio; SE = standard error; CI = confidence interval; *p* = significance value.

a *Demographic and genetic variables*: age; sex; 10 principal components (PCs); C-reactive Protein (CRP) polygenic score (PGS); White Blood Cell Counts (WBCC) PGS; Insulin Growth Factor-1 (IGF-1) PGS; Anxiety PGS; Depression PGS; Schizophrenia PGS; Insomnia PGS; Pain PGS.

b All variables: age; sex; 10 PCs; CRP PGS; WBCC PGS; IGF-1 PGS; PGS; Anxiety PGS; Depression PGS; Schizophrenia PGS; Insomnia PGS; Pain PGS; education; wealth; smoking status; alcohol consumption; mobility.

c Additional variables: medication, physical activity; BMI.

d Additional variables: health (i.e., chronic lung disease; coronary heart disease; abnormal heart rhythm; heart murmur; congestive heart failure; angina; hypertension; diabetes; cancer; Parkinson's; Alzheimer's; dementia; asthma; arthritis; osteoporosis; psychiatric disorder).

***Table S9v.*** **Longitudinal associations between C-reactive protein and hospitalisation for diseases of the digestive system (*n*=3,188)**

| **Adjustments** | **CRP** | | | | |
|---|---|---|---|---|---|
| | **HR** | **SE** | **95% CI** | | ***p*** |
| **Digestive Disorders** | | | | | |
| Model 1: *Unadjusted* | 1.14 | 0.05 | 1.05 | 1.24 | 0.002 |
| Model 2: *Model 1 + demographics & genetics* [a] | 1.06 | 0.05 | 0.97 | 1.16 | 0.185 |
| Model 3: *Model 2 + Fully Adjusted* [b] | 1.02 | 0.05 | 0.94 | 1.12 | 0.620 |
| Model 2a: *Demographics* | 1.08 | 0.05 | 0.99 | 1.17 | 0.096 |
| Model 2b: *Genetics* | 1.13 | 0.05 | 1.04 | 1.23 | 0.006 |
| Model 3a: Model 3 + *Medication* | 1.02 | 0.05 | 0.94 | 1.12 | 0.623 |
| Model 3b: Model 3 + *Physical Activity* | 1.02 | 0.05 | 0.93 | 1.11 | 0.750 |
| Model 3c: Model 3 + *Body Mass Index* | 1.02 | 0.05 | 0.93 | 1.12 | 0.725 |
| Model 3d: Model 3 + *Fully Adjusted* [c] | 1.01 | 0.05 | 0.92 | 1.11 | 0.827 |
| Model 4a: Model 2 + *Health* | 1.06 | 0.05 | 0.97 | 1.16 | 0.185 |
| Model 4b: Model 3 + *Health* | 1.02 | 0.05 | 0.93 | 1.11 | 0.747 |
| Model 4c: Model 3 + *Fully Adjusted* [d] | 1.00 | 0.05 | 0.91 | 1.10 | 0.933 |

**Notes:** The *low-risk* group is the reference; HR = hazard ratio; SE = standard error; CI = confidence interval; *p* = significance value.

a *Demographic and genetic variables*: age; sex; 10 principal components (PCs); C-reactive Protein (CRP) polygenic score (PGS); White Blood Cell Counts (WBCC) PGS; Insulin Growth Factor-1 (IGF-1) PGS; Anxiety PGS; Depression PGS; Schizophrenia PGS; Insomnia PGS; Pain PGS.

b All variables: age; sex; 10 PCs; CRP PGS; WBCC PGS; IGF-1 PGS; PGS; Anxiety PGS; Depression PGS; Schizophrenia PGS; Insomnia PGS; Pain PGS; education; wealth; smoking status; alcohol consumption; mobility.

c Additional variables: medication, physical activity; BMI.

d Additional variables: health (i.e., chronic lung disease; coronary heart disease; abnormal heart rhythm; heart murmur; congestive heart failure; angina; hypertension; diabetes; cancer; Parkinson's; Alzheimer's; dementia; asthma; arthritis; osteoporosis; psychiatric disorder).

***Table S9w.*** **Longitudinal associations between fibrinogen and hospitalisation for diseases of the digestive system (*n*=3,188)**

| **Adjustments** | **Fb** | | | | |
|---|---|---|---|---|---|
| | **HR** | **SE** | **95% CI** | | ***p*** |
| **Digestive Disorders** | | | | | |
| Model 1: *Unadjusted* | 1.11 | 0.06 | 1.00 | 1.23 | 0.045 |
| Model 2: *Model 1 + demographics & genetics* [a] | 1.03 | 0.06 | 0.92 | 1.14 | 0.609 |
| Model 3: *Model 2 + Fully Adjusted* [b] | 0.97 | 0.05 | 0.87 | 1.08 | 0.580 |
| Model 2a: *Demographics* | 1.05 | 0.06 | 0.94 | 1.16 | 0.415 |
| Model 2b: *Genetics* | 1.09 | 0.06 | 0.98 | 1.21 | 0.106 |
| Model 3a: Model 3 + *Medication* | 0.97 | 0.05 | 0.87 | 1.08 | 0.568 |
| Model 3b: Model 3 + *Physical Activity* | 0.97 | 0.05 | 0.87 | 1.08 | 0.527 |
| Model 3c: Model 3 + *Body Mass Index* | 0.96 | 0.06 | 0.86 | 1.08 | 0.515 |
| Model 3d: Model 3 + *Fully Adjusted* [c] | 0.96 | 0.06 | 0.86 | 1.07 | 0.471 |
| Model 4a: Model 2 + *Health* | 1.02 | 0.06 | 0.92 | 1.14 | 0.686 |
| Model 4b: Model 3 + *Health* | 0.97 | 0.05 | 0.87 | 1.08 | 0.582 |
| Model 4c: Model 3 + *Fully Adjusted* [d] | 0.96 | 0.06 | 0.86 | 1.08 | 0.487 |

**Notes:** The *low-risk* group is the reference; HR = hazard ratio; SE = standard error; CI = confidence interval; *p* = significance value.

a *Demographic and genetic variables*: age; sex; 10 principal components (PCs); C-reactive Protein (CRP) polygenic score (PGS); White Blood Cell Counts (WBCC) PGS; Insulin Growth Factor-1 (IGF-1) PGS; Anxiety PGS; Depression PGS; Schizophrenia PGS; Insomnia PGS; Pain PGS.

b All variables: age; sex; 10 PCs; CRP PGS; WBCC PGS; IGF-1 PGS; PGS; Anxiety PGS; Depression PGS; Schizophrenia PGS; Insomnia PGS; Pain PGS; education; wealth; smoking status; alcohol consumption; mobility.

c Additional variables: medication, physical activity; BMI.

d Additional variables: health (i.e., chronic lung disease; coronary heart disease; abnormal heart rhythm; heart murmur; congestive heart failure; angina; hypertension; diabetes; cancer; Parkinson's; Alzheimer's; dementia; asthma; arthritis; osteoporosis; psychiatric disorder).

***Table S9x.*** **Longitudinal associations between white blood cell counts and hospitalisation for diseases of the digestive system (*n*=3,188)**

| Adjustments | WBCC | | | | |
|---|---|---|---|---|---|
| | HR | SE | 95% CI | | *p* |
| **Digestive Disorders** | | | | | |
| Model 1: *Unadjusted* | 1.27 | 0.15 | 1.00 | 1.60 | 0.046 |
| Model 2: *Model 1 + demographics & genetics* [a] | 1.15 | 0.14 | 0.90 | 1.46 | 0.262 |
| Model 3: *Model 2 + Fully Adjusted* [b] | 1.01 | 0.13 | 0.78 | 1.29 | 0.962 |
| Model 2a: *Demographics* | 1.14 | 0.14 | 0.90 | 1.45 | 0.272 |
| Model 2b: *Genetics* | 1.23 | 0.15 | 0.98 | 1.56 | 0.079 |
| Model 3a: Model 3 + *Medication* | 1.00 | 0.13 | 0.78 | 1.29 | 0.992 |
| Model 3b: Model 3 + *Physical Activity* | 0.99 | 0.13 | 0.77 | 1.28 | 0.961 |
| Model 3c: Model 3 + *Body Mass Index* | 1.00 | 0.13 | 0.77 | 1.28 | 0.969 |
| Model 3d: Model 3 + *Fully Adjusted* [c] | 0.98 | 0.13 | 0.76 | 1.27 | 0.885 |
| Model 4a: Model 2 + *Health* | 1.11 | 0.14 | 0.87 | 1.41 | 0.395 |
| Model 4b: Model 3 + *Health* | 0.99 | 0.13 | 0.77 | 1.27 | 0.913 |
| Model 4c: Model 3 + *Fully Adjusted* [d] | 0.96 | 0.13 | 0.75 | 1.25 | 0.781 |

**Notes:** The *low-risk* group is the reference; HR = hazard ratio; SE = standard error; CI = confidence interval; *p* = significance value.

a *Demographic and genetic variables*: age; sex; 10 principal components (PCs); C-reactive Protein (CRP) polygenic score (PGS); White Blood Cell Counts (WBCC) PGS; Insulin Growth Factor-1 (IGF-1) PGS; Anxiety PGS; Depression PGS; Schizophrenia PGS; Insomnia PGS; Pain PGS.

b All variables: age; sex; 10 PCs; CRP PGS; WBCC PGS; IGF-1 PGS; PGS; Anxiety PGS; Depression PGS; Schizophrenia PGS; Insomnia PGS; Pain PGS; education; wealth; smoking status; alcohol consumption; mobility.

c Additional variables: medication, physical activity; BMI.

d Additional variables: health (i.e., chronic lung disease; coronary heart disease; abnormal heart rhythm; heart murmur; congestive heart failure; angina; hypertension; diabetes; cancer; Parkinson's; Alzheimer's; dementia; asthma; arthritis; osteoporosis; psychiatric disorder).

***Table S9y.*** **Longitudinal associations between insulin growth factor-1 and hospitalisation for diseases of the digestive system (*n*=3,188)**

| Adjustments | IGF-1 | | | | |
|---|---|---|---|---|---|
| | HR | SE | 95% CI | | *p* |
| **Digestive Disorders** | | | | | |
| Model 1: *Unadjusted* | 0.72 | 0.07 | 0.61 | 0.86 | <0.001 |
| Model 2: *Model 1 + demographics & genetics* [a] | 1.15 | 0.14 | 0.90 | 1.46 | 0.262 |
| Model 3: *Model 2 + Fully Adjusted* [b] | 1.01 | 0.13 | 0.78 | 1.29 | 0.962 |
| Model 2a: *Demographics* | 1.14 | 0.14 | 0.90 | 1.45 | 0.272 |
| Model 2b: *Genetics* | 1.23 | 0.15 | 0.98 | 1.56 | 0.079 |
| Model 3a: Model 3 + *Medication* | 1.00 | 0.13 | 0.78 | 1.29 | 0.992 |
| Model 3b: Model 3 + *Physical Activity* | 0.99 | 0.13 | 0.77 | 1.28 | 0.961 |
| Model 3c: Model 3 + *Body Mass Index* | 1.00 | 0.13 | 0.77 | 1.28 | 0.969 |
| Model 3d: Model 3 + *Fully Adjusted* [c] | 0.98 | 0.13 | 0.76 | 1.27 | 0.885 |
| Model 4a: Model 2 + *Health* | 1.11 | 0.14 | 0.87 | 1.41 | 0.395 |
| Model 4b: Model 3 + *Health* | 0.99 | 0.13 | 0.77 | 1.27 | 0.913 |
| Model 4c: Model 3 + *Fully Adjusted* [d] | 0.96 | 0.13 | 0.75 | 1.25 | 0.781 |

**Notes:** The *low-risk* group is the reference; HR = hazard ratio; SE = standard error; CI = confidence interval; *p* = significance value.

a *Demographic and genetic variables*: age; sex; 10 principal components (PCs); C-reactive Protein (CRP) polygenic score (PGS); White Blood Cell Counts (WBCC) PGS; Insulin Growth Factor-1 (IGF-1) PGS; Anxiety PGS; Depression PGS; Schizophrenia PGS; Insomnia PGS; Pain PGS.

b All variables: age; sex; 10 PCs; CRP PGS; WBCC PGS; IGF-1 PGS; PGS; Anxiety PGS; Depression PGS; Schizophrenia PGS; Insomnia PGS; Pain PGS; education; wealth; smoking status; alcohol consumption; mobility.

c Additional variables: medication, physical activity; BMI.

d Additional variables: health (i.e., chronic lung disease; coronary heart disease; abnormal heart rhythm; heart murmur; congestive heart failure; angina; hypertension; diabetes; cancer; Parkinson's; Alzheimer's; dementia; asthma; arthritis; osteoporosis; psychiatric disorder).

*Table S9z.* **Longitudinal associations between immune and neuroendocrine profiles and hospitalisation for endocrine, nutritional and metabolic diseases (*n*=3,749)**

| Adjustments | Immune and Neuroendocrine Profiles | | | | |
|---|---|---|---|---|---|
| | HR | SE | 95% CI | | *p* |
| ***Moderate-risk* Profile \| Endocrine Disorders** | | | | | |
| Model 1: *Unadjusted* | 1.57 | 0.10 | 1.39 | 1.78 | <0.001 |
| Model 2: *Model 1 + demographics & genetics* [a] | 1.41 | 0.09 | 1.24 | 1.60 | <0.001 |
| Model 3: *Model 2 + Fully Adjusted* [b] | 1.26 | 0.08 | 1.11 | 1.43 | <0.001 |
| Model 2a: *Demographics* | 1.41 | 0.09 | 1.25 | 1.60 | <0.001 |
| Model 2b: *Genetics* | 1.57 | 0.10 | 1.39 | 1.78 | <0.001 |
| Model 3a: Model 3 + *Medication* | 1.26 | 0.08 | 1.11 | 1.43 | <0.001 |
| Model 3b: Model 3 + *Physical Activity* | 1.24 | 0.08 | 1.09 | 1.41 | 0.001 |
| Model 3c: Model 3 + *Body Mass Index* | 1.14 | 0.08 | 1.00 | 1.30 | 0.044 |
| Model 3d: Model 3 + *Fully Adjusted* [c] | 1.13 | 0.08 | 0.99 | 1.29 | 0.070 |
| Model 4a: Model 2 + *Health* | 1.41 | 0.09 | 1.24 | 1.60 | <0.001 |
| Model 4b: Model 3 + *Health* | 1.25 | 0.08 | 1.10 | 1.42 | 0.001 |
| Model 4c: Model 3 + *Fully Adjusted* [d] | 1.13 | 0.08 | 0.99 | 1.28 | 0.077 |
| ***High-risk* Profile \| Endocrine Disorders** | | | | | |
| Model 1: *Unadjusted* | 1.81 | 0.16 | 1.52 | 2.16 | <0.001 |
| Model 2: *Model 1 + demographics & genetics* [a] | 1.62 | 0.15 | 1.36 | 1.93 | <0.001 |
| Model 3: *Model 2 + Fully Adjusted* [b] | 1.38 | 0.13 | 1.16 | 1.66 | <0.001 |
| Model 2a: *Demographics* | 1.62 | 0.15 | 1.35 | 1.93 | <0.001 |
| Model 2b: *Genetics* | 1.81 | 0.16 | 1.52 | 2.16 | <0.001 |
| Model 3a: Model 3 + *Medication* | 1.38 | 0.13 | 1.16 | 1.66 | <0.001 |
| Model 3b: Model 3 + *Physical Activity* | 1.33 | 0.12 | 1.11 | 1.59 | 0.002 |
| Model 3c: Model 3 + *Body Mass Index* | 1.23 | 0.12 | 1.03 | 1.48 | 0.025 |
| Model 3d: Model 3 + *Fully Adjusted* [c] | 1.19 | 0.11 | 0.99 | 1.43 | 0.066 |
| Model 4a: Model 2 + *Health* | 1.62 | 0.15 | 1.36 | 1.93 | <0.001 |
| Model 4b: Model 3 + *Health* | 1.38 | 0.13 | 1.15 | 1.65 | <0.001 |
| Model 4c: Model 3 + *Fully Adjusted* [d] | 1.19 | 0.11 | 0.99 | 1.43 | 0.070 |

**Notes:** The *low-risk* group is the reference; HR = hazard ratio; SE = standard error; CI = confidence interval; *p* = significance value.

a *Demographic and genetic variables*: age; sex; 10 principal components (PCs); C-reactive Protein (CRP) polygenic score (PGS); White Blood Cell Counts (WBCC) PGS; Insulin Growth Factor-1 (IGF-1) PGS; Anxiety PGS; Depression PGS; Schizophrenia PGS; Insomnia PGS; Pain PGS.

b All variables: age; sex; 10 PCs; CRP PGS; WBCC PGS; IGF-1 PGS; PGS; Anxiety PGS; Depression PGS; Schizophrenia PGS; Insomnia PGS; Pain PGS; education; wealth; smoking status; alcohol consumption; mobility.

c Additional variables: medication, physical activity; BMI.

d Additional variables: health (i.e., chronic lung disease; coronary heart disease; abnormal heart rhythm; heart murmur; congestive heart failure; angina; hypertension; diabetes; cancer; Parkinson's; Alzheimer's; dementia; asthma; arthritis; osteoporosis; psychiatric disorder).

***Table S9aa.*** **Longitudinal associations between C-reactive protein and hospitalisation for endocrine, nutritional and metabolic diseases (*n*=3,749)**

| Adjustments | CRP | | | | |
|---|---|---|---|---|---|
| | HR | SE | 95% CI | | *p* |
| **Endocrine Disorders** | | | | | |
| Model 1: *Unadjusted* | 1.44 | 0.06 | 1.33 | 1.56 | <0.001 |
| Model 2: *Model 1 + demographics & genetics* [a] | 1.35 | 0.06 | 1.24 | 1.47 | <0.001 |
| Model 3: *Model 2 + Fully Adjusted* [b] | 1.24 | 0.05 | 1.13 | 1.35 | <0.001 |
| Model 2a: *Demographics* | 1.35 | 0.06 | 1.24 | 1.46 | <0.001 |
| Model 2b: *Genetics* | 1.44 | 0.06 | 1.33 | 1.56 | <0.001 |
| Model 3a: Model 3 + *Medication* | 1.24 | 0.05 | 1.13 | 1.35 | <0.001 |
| Model 3b: Model 3 + *Physical Activity* | 1.21 | 0.05 | 1.11 | 1.32 | <0.001 |
| Model 3c: Model 3 + *Body Mass Index* | 1.15 | 0.05 | 1.05 | 1.26 | 0.003 |
| Model 3d: Model 3 + *Fully Adjusted* [c] | 1.13 | 0.05 | 1.03 | 1.23 | 0.012 |
| Model 4a: Model 2 + *Health* | 1.35 | 0.06 | 1.24 | 1.47 | <0.001 |
| Model 4b: Model 3 + *Health* | 1.23 | 0.05 | 1.13 | 1.34 | <0.001 |
| Model 4c: Model 3 + *Fully Adjusted* [d] | 1.12 | 0.05 | 1.03 | 1.23 | 0.013 |

**Notes:** The *low-risk* group is the reference; HR = hazard ratio; SE = standard error; CI = confidence interval; *p* = significance value.

a *Demographic and genetic variables*: age; sex; 10 principal components (PCs); C-reactive Protein (CRP) polygenic score (PGS); White Blood Cell Counts (WBCC) PGS; Insulin Growth Factor-1 (IGF-1) PGS; Anxiety PGS; Depression PGS; Schizophrenia PGS; Insomnia PGS; Pain PGS.

b All variables: age; sex; 10 PCs; CRP PGS; WBCC PGS; IGF-1 PGS; PGS; Anxiety PGS; Depression PGS; Schizophrenia PGS; Insomnia PGS; Pain PGS; education; wealth; smoking status; alcohol consumption; mobility.

c Additional variables: medication, physical activity; BMI.

d Additional variables: health (i.e., chronic lung disease; coronary heart disease; abnormal heart rhythm; heart murmur; congestive heart failure; angina; hypertension; diabetes; cancer; Parkinson's; Alzheimer's; dementia; asthma; arthritis; osteoporosis; psychiatric disorder).

***Table S9ab.*** **Longitudinal associations between fibrinogen and hospitalisation for endocrine, nutritional and metabolic diseases (*n*=3,749)**

| **Adjustments** | **Fb** | | | | |
|---|---|---|---|---|---|
| | **HR** | **SE** | **95% CI** | | ***p*** |
| **Endocrine Disorders** | | | | | |
| Model 1: *Unadjusted* | 1.47 | 0.07 | 1.33 | 1.62 | <0.001 |
| Model 2: *Model 1 + demographics & genetics* [a] | 1.36 | 0.07 | 1.23 | 1.51 | <0.001 |
| Model 3: *Model 2 + Fully Adjusted* [b] | 1.23 | 0.07 | 1.10 | 1.36 | <0.001 |
| Model 2a: *Demographics* | 1.35 | 0.07 | 1.22 | 1.50 | <0.001 |
| Model 2b: *Genetics* | 1.47 | 0.08 | 1.33 | 1.62 | <0.001 |
| Model 3a: Model 3 + *Medication* | 1.23 | 0.07 | 1.10 | 1.36 | <0.001 |
| Model 3b: Model 3 + *Physical Activity* | 1.20 | 0.07 | 1.08 | 1.34 | 0.001 |
| Model 3c: Model 3 + *Body Mass Index* | 1.16 | 0.06 | 1.04 | 1.30 | 0.006 |
| Model 3d: Model 3 + *Fully Adjusted* [c] | 1.15 | 0.06 | 1.03 | 1.28 | 0.013 |
| Model 4a: Model 2 + *Health* | 1.35 | 0.07 | 1.22 | 1.50 | <0.001 |
| Model 4b: Model 3 + *Health* | 1.22 | 0.07 | 1.10 | 1.36 | <0.001 |
| Model 4c: Model 3 + *Fully Adjusted* [d] | 1.15 | 0.06 | 1.03 | 1.28 | 0.013 |

**Notes:** The *low-risk* group is the reference; HR = hazard ratio; SE = standard error; CI = confidence interval; *p* = significance value.

a *Demographic and genetic variables*: age; sex; 10 principal components (PCs); C-reactive Protein (CRP) polygenic score (PGS); White Blood Cell Counts (WBCC) PGS; Insulin Growth Factor-1 (IGF-1) PGS; Anxiety PGS; Depression PGS; Schizophrenia PGS; Insomnia PGS; Pain PGS.

b All variables: age; sex; 10 PCs; CRP PGS; WBCC PGS; IGF-1 PGS; PGS; Anxiety PGS; Depression PGS; Schizophrenia PGS; Insomnia PGS; Pain PGS; education; wealth; smoking status; alcohol consumption; mobility.

c Additional variables: medication, physical activity; BMI.

d Additional variables: health (i.e., chronic lung disease; coronary heart disease; abnormal heart rhythm; heart murmur; congestive heart failure; angina; hypertension; diabetes; cancer; Parkinson's; Alzheimer's; dementia; asthma; arthritis; osteoporosis; psychiatric disorder).

***Table S9ac.*** **Longitudinal associations between white blood cell counts and hospitalisation for endocrine, nutritional and metabolic diseases (*n*=3,749)**

| Adjustments | WBCC | | | | |
|---|---|---|---|---|---|
| | HR | SE | 95% CI | | *p* |
| **Endocrine Disorders** | | | | | |
| Model 1: *Unadjusted* | 2.11 | 0.25 | 1.68 | 2.66 | <0.001 |
| Model 2: *Model 1 + demographics & genetics* [a] | 2.02 | 0.24 | 1.60 | 2.56 | <0.001 |
| Model 3: *Model 2 + Fully Adjusted* [b] | 1.54 | 0.19 | 1.20 | 1.97 | 0.001 |
| Model 2a: *Demographics* | 2.00 | 0.24 | 1.58 | 2.53 | <0.001 |
| Model 2b: *Genetics* | 2.11 | 0.25 | 1.68 | 2.66 | <0.001 |
| Model 3a: Model 3 + *Medication* | 1.54 | 0.19 | 1.20 | 1.97 | 0.001 |
| Model 3b: Model 3 + *Physical Activity* | 1.50 | 0.19 | 1.17 | 1.92 | 0.001 |
| Model 3c: Model 3 + *Body Mass Index* | 1.36 | 0.18 | 1.06 | 1.75 | 0.015 |
| Model 3d: Model 3 + *Fully Adjusted* [c] | 1.34 | 0.17 | 1.04 | 1.72 | 0.023 |
| Model 4a: Model 2 + *Health* | 1.98 | 0.24 | 1.56 | 2.51 | <0.001 |
| Model 4b: Model 3 + *Health* | 1.53 | 0.19 | 1.19 | 1.95 | 0.001 |
| Model 4c: Model 3 + *Fully Adjusted* [d] | 1.33 | 0.17 | 1.04 | 1.71 | 0.025 |

**Notes:** The *low-risk* group is the reference; HR = hazard ratio; SE = standard error; CI = confidence interval; *p* = significance value.

a *Demographic and genetic variables*: age; sex; 10 principal components (PCs); C-reactive Protein (CRP) polygenic score (PGS); White Blood Cell Counts (WBCC) PGS; Insulin Growth Factor-1 (IGF-1) PGS; Anxiety PGS; Depression PGS; Schizophrenia PGS; Insomnia PGS; Pain PGS.

b All variables: age; sex; 10 PCs; CRP PGS; WBCC PGS; IGF-1 PGS; PGS; Anxiety PGS; Depression PGS; Schizophrenia PGS; Insomnia PGS; Pain PGS; education; wealth; smoking status; alcohol consumption; mobility.

c Additional variables: medication, physical activity; BMI.

d Additional variables: health (i.e., chronic lung disease; coronary heart disease; abnormal heart rhythm; heart murmur; congestive heart failure; angina; hypertension; diabetes; cancer; Parkinson's; Alzheimer's; dementia; asthma; arthritis; osteoporosis; psychiatric disorder).

***Table S9ad.*** **Longitudinal associations between insulin growth factor-1 and hospitalisation for endocrine, nutritional and metabolic diseases (*n*=3,749)**

| Adjustments | IGF-1 | | | | |
|---|---|---|---|---|---|
| | HR | SE | 95% CI | | *p* |
| **Endocrine Disorders** | | | | | |
| Model 1: *Unadjusted* | 0.65 | 0.06 | 0.54 | 0.77 | <0.001 |
| Model 2: *Model 1 + demographics & genetics* [a] | 0.85 | 0.08 | 0.71 | 1.02 | 0.075 |
| Model 3: *Model 2 + Fully Adjusted* [b] | 0.87 | 0.08 | 0.73 | 1.03 | 0.106 |
| Model 2a: *Demographics* | 0.85 | 0.08 | 0.71 | 1.02 | 0.073 |
| Model 2b: *Genetics* | 0.65 | 0.06 | 0.54 | 0.77 | <0.001 |
| Model 3a: Model 3 + *Medication* | 0.87 | 0.08 | 0.73 | 1.03 | 0.105 |
| Model 3b: Model 3 + *Physical Activity* | 0.87 | 0.08 | 0.73 | 1.04 | 0.132 |
| Model 3c: Model 3 + *Body Mass Index* | 0.87 | 0.08 | 0.73 | 1.04 | 0.123 |
| Model 3d: Model 3 + *Fully Adjusted* [c] | 0.88 | 0.08 | 0.74 | 1.05 | 0.159 |
| Model 4a: Model 2 + *Health* | 0.85 | 0.08 | 0.71 | 1.01 | 0.068 |
| Model 4b: Model 3 + *Health* | 0.86 | 0.08 | 0.72 | 1.03 | 0.100 |
| Model 4c: Model 3 + *Fully Adjusted* [d] | 0.88 | 0.08 | 0.74 | 1.05 | 0.153 |

**Notes:** The *low-risk* group is the reference; HR = hazard ratio; SE = standard error; CI = confidence interval; *p* = significance value.

a *Demographic and genetic variables*: age; sex; 10 principal components (PCs); C-reactive Protein (CRP) polygenic score (PGS); White Blood Cell Counts (WBCC) PGS; Insulin Growth Factor-1 (IGF-1) PGS; Anxiety PGS; Depression PGS; Schizophrenia PGS; Insomnia PGS; Pain PGS.

b All variables: age; sex; 10 PCs; CRP PGS; WBCC PGS; IGF-1 PGS; PGS; Anxiety PGS; Depression PGS; Schizophrenia PGS; Insomnia PGS; Pain PGS; education; wealth; smoking status; alcohol consumption; mobility.

c Additional variables: medication, physical activity; BMI.

d Additional variables: health (i.e., chronic lung disease; coronary heart disease; abnormal heart rhythm; heart murmur; congestive heart failure; angina; hypertension; diabetes; cancer; Parkinson's; Alzheimer's; dementia; asthma; arthritis; osteoporosis; psychiatric disorder).

***Table S9ae.*** **Longitudinal associations between immune and neuroendocrine profiles and hospitalisation for diseases of the genitourinary system (*n*=3,524)**

| Adjustments | Immune and Neuroendocrine Profiles | | | | |
|---|---|---|---|---|---|
| | HR | SE | 95% CI | | *p* |
| ***Moderate-risk*** **Profile \| Genitourinary Disorders** | | | | | |
| Model 1: *Unadjusted* | 1.45 | 0.10 | 1.26 | 1.67 | <0.001 |
| Model 2: *Model 1 + demographics & genetics* [a] | 1.28 | 0.09 | 1.11 | 1.48 | 0.001 |
| Model 3: *Model 2 + Fully Adjusted* [b] | 1.18 | 0.09 | 1.02 | 1.36 | 0.027 |
| Model 2a: *Demographics* | 1.27 | 0.09 | 1.10 | 1.46 | 0.001 |
| Model 2b: *Genetics* | 1.45 | 0.10 | 1.26 | 1.67 | <0.001 |
| Model 3a: Model 3 + *Medication* | 1.18 | 0.09 | 1.02 | 1.36 | 0.028 |
| Model 3b: Model 3 + *Physical Activity* | 1.16 | 0.09 | 1.01 | 1.34 | 0.042 |
| Model 3c: Model 3 + *Body Mass Index* | 1.12 | 0.09 | 0.97 | 1.30 | 0.131 |
| Model 3d: Model 3 + *Fully Adjusted* [c] | 1.11 | 0.09 | 0.96 | 1.29 | 0.163 |
| Model 4a: Model 2 + *Health* | 1.27 | 0.09 | 1.10 | 1.46 | 0.001 |
| Model 4b: Model 3 + *Health* | 1.17 | 0.09 | 1.01 | 1.36 | 0.032 |
| Model 4c: Model 3 + *Fully Adjusted* [d] | 1.11 | 0.09 | 0.95 | 1.29 | 0.180 |
| ***High-risk*** **Profile \| Genitourinary Disorders** | | | | | |
| Model 1: *Unadjusted* | 1.68 | 0.17 | 1.37 | 2.05 | <0.001 |
| Model 2: *Model 1 + demographics & genetics* [a] | 1.50 | 0.16 | 1.22 | 1.84 | <0.001 |
| Model 3: *Model 2 + Fully Adjusted* [b] | 1.35 | 0.14 | 1.10 | 1.65 | 0.004 |
| Model 2a: *Demographics* | 1.49 | 0.15 | 1.22 | 1.83 | <0.001 |
| Model 2b: *Genetics* | 1.66 | 0.17 | 1.36 | 2.03 | <0.001 |
| Model 3a: Model 3 + *Medication* | 1.35 | 0.14 | 1.10 | 1.66 | 0.004 |
| Model 3b: Model 3 + *Physical Activity* | 1.29 | 0.14 | 1.05 | 1.59 | 0.014 |
| Model 3c: Model 3 + *Body Mass Index* | 1.28 | 0.14 | 1.04 | 1.58 | 0.022 |
| Model 3d: Model 3 + *Fully Adjusted* [c] | 1.24 | 0.13 | 1.00 | 1.53 | 0.048 |
| Model 4a: Model 2 + *Health* | 1.49 | 0.15 | 1.21 | 1.82 | <0.001 |
| Model 4b: Model 3 + *Health* | 1.34 | 0.14 | 1.09 | 1.65 | 0.005 |
| Model 4c: Model 3 + *Fully Adjusted* [d] | 1.23 | 0.13 | 1.00 | 1.52 | 0.050 |

**Notes:** The *low-risk* group is the reference; HR = hazard ratio; SE = standard error; CI = confidence interval; *p* = significance value.

a *Demographic and genetic variables*: age; sex; 10 principal components (PCs); C-reactive Protein (CRP) polygenic score (PGS); White Blood Cell Counts (WBCC) PGS; Insulin Growth Factor-1 (IGF-1) PGS; Anxiety PGS; Depression PGS; Schizophrenia PGS; Insomnia PGS; Pain PGS.

b All variables: age; sex; 10 PCs; CRP PGS; WBCC PGS; IGF-1 PGS; PGS; Anxiety PGS; Depression PGS; Schizophrenia PGS; Insomnia PGS; Pain PGS; education; wealth; smoking status; alcohol consumption; mobility.

c Additional variables: medication, physical activity; BMI.

d Additional variables: health (i.e., chronic lung disease; coronary heart disease; abnormal heart rhythm; heart murmur; congestive heart failure; angina; hypertension; diabetes; cancer; Parkinson's; Alzheimer's; dementia; asthma; arthritis; osteoporosis; psychiatric disorder).

***Table S9af.*** **Longitudinal associations between C-reactive protein and hospitalisation for diseases of the genitourinary system (*n*=3,524)**

| Adjustments | CRP | | | | |
|---|---|---|---|---|---|
| | HR | SE | 95% CI | | *p* |
| **Genitourinary Disorders** | | | | | |
| Model 1: *Unadjusted* | 1.35 | 0.06 | 1.23 | 1.48 | <0.001 |
| Model 2: *Model 1 + demographics & genetics* [a] | 1.26 | 0.06 | 1.14 | 1.39 | <0.001 |
| Model 3: *Model 2 + Fully Adjusted* [b] | 1.17 | 0.06 | 1.06 | 1.30 | 0.002 |
| Model 2a: *Demographics* | 1.25 | 0.06 | 1.14 | 1.38 | <0.001 |
| Model 2b: *Genetics* | 1.34 | 0.06 | 1.22 | 1.48 | <0.001 |
| Model 3a: Model 3 + *Medication* | 1.18 | 0.06 | 1.06 | 1.30 | 0.001 |
| Model 3b: Model 3 + *Physical Activity* | 1.15 | 0.06 | 1.04 | 1.27 | 0.007 |
| Model 3c: Model 3 + *Body Mass Index* | 1.13 | 0.06 | 1.02 | 1.26 | 0.018 |
| Model 3d: Model 3 + *Fully Adjusted* [c] | 1.11 | 0.06 | 1.00 | 1.24 | 0.044 |
| Model 4a: Model 2 + *Health* | 1.25 | 0.06 | 1.13 | 1.38 | <0.001 |
| Model 4b: Model 3 + *Health* | 1.17 | 0.06 | 1.06 | 1.29 | 0.002 |
| Model 4c: Model 3 + *Fully Adjusted* [d] | 1.11 | 0.06 | 1.00 | 1.23 | 0.049 |

**Notes:** The *low-risk* group is the reference; HR = hazard ratio; SE = standard error; CI = confidence interval; *p* = significance value.

a *Demographic and genetic variables*: age; sex; 10 principal components (PCs); C-reactive Protein (CRP) polygenic score (PGS); White Blood Cell Counts (WBCC) PGS; Insulin Growth Factor-1 (IGF-1) PGS; Anxiety PGS; Depression PGS; Schizophrenia PGS; Insomnia PGS; Pain PGS.

b All variables: age; sex; 10 PCs; CRP PGS; WBCC PGS; IGF-1 PGS; PGS; Anxiety PGS; Depression PGS; Schizophrenia PGS; Insomnia PGS; Pain PGS; education; wealth; smoking status; alcohol consumption; mobility.

c Additional variables: medication, physical activity; BMI.

d Additional variables: health (i.e., chronic lung disease; coronary heart disease; abnormal heart rhythm; heart murmur; congestive heart failure; angina; hypertension; diabetes; cancer; Parkinson's; Alzheimer's; dementia; asthma; arthritis; osteoporosis; psychiatric disorder).

***Table S9ag.*** **Longitudinal associations between fibrinogen and hospitalisation for diseases of the genitourinary system (*n*=3,524)**

| Adjustments | Fb | | | | |
|---|---|---|---|---|---|
| | HR | SE | 95% CI | | *p* |
| **Genitourinary Disorders** | | | | | |
| Model 1: *Unadjusted* | 1.37 | 0.08 | 1.22 | 1.53 | <0.001 |
| Model 2: *Model 1 + demographics & genetics* [a] | 1.26 | 0.08 | 1.12 | 1.42 | <0.001 |
| Model 3: *Model 2 + Fully Adjusted* [b] | 1.18 | 0.07 | 1.04 | 1.33 | 0.009 |
| Model 2a: *Demographics* | 1.26 | 0.08 | 1.12 | 1.42 | <0.001 |
| Model 2b: *Genetics* | 1.37 | 0.08 | 1.22 | 1.54 | <0.001 |
| Model 3a: Model 3 + *Medication* | 1.18 | 0.07 | 1.04 | 1.33 | 0.009 |
| Model 3b: Model 3 + *Physical Activity* | 1.16 | 0.07 | 1.03 | 1.31 | 0.017 |
| Model 3c: Model 3 + *Body Mass Index* | 1.15 | 0.07 | 1.02 | 1.30 | 0.029 |
| Model 3d: Model 3 + *Fully Adjusted* [c] | 1.14 | 0.07 | 1.01 | 1.29 | 0.041 |
| Model 4a: Model 2 + *Health* | 1.26 | 0.08 | 1.12 | 1.42 | <0.001 |
| Model 4b: Model 3 + *Health* | 1.18 | 0.07 | 1.04 | 1.33 | 0.009 |
| Model 4c: Model 3 + *Fully Adjusted* [d] | 1.14 | 0.07 | 1.01 | 1.29 | 0.041 |

**Notes:** The *low-risk* group is the reference; HR = hazard ratio; SE = standard error; CI = confidence interval; *p* = significance value.

a *Demographic and genetic variables*: age; sex; 10 principal components (PCs); C-reactive Protein (CRP) polygenic score (PGS); White Blood Cell Counts (WBCC) PGS; Insulin Growth Factor-1 (IGF-1) PGS; Anxiety PGS; Depression PGS; Schizophrenia PGS; Insomnia PGS; Pain PGS.

b All variables: age; sex; 10 PCs; CRP PGS; WBCC PGS; IGF-1 PGS; PGS; Anxiety PGS; Depression PGS; Schizophrenia PGS; Insomnia PGS; Pain PGS; education; wealth; smoking status; alcohol consumption; mobility.

c Additional variables: medication, physical activity; BMI.

d Additional variables: health (i.e., chronic lung disease; coronary heart disease; abnormal heart rhythm; heart murmur; congestive heart failure; angina; hypertension; diabetes; cancer; Parkinson's; Alzheimer's; dementia; asthma; arthritis; osteoporosis; psychiatric disorder).

***Table S9ah.*** **Longitudinal associations between white blood cell counts and hospitalisation for diseases of the genitourinary system (*n*=3,524)**

| Adjustments | WBCC | | | | |
|---|---|---|---|---|---|
| | HR | SE | 95% CI | | *p* |
| **Genitourinary Disorders** | | | | | |
| Model 1: *Unadjusted* | 1.92 | 0.26 | 1.47 | 2.49 | <0.001 |
| Model 2: *Model 1 + demographics & genetics* [a] | 1.75 | 0.25 | 1.33 | 2.30 | <0.001 |
| Model 3: *Model 2 + Fully Adjusted* [b] | 1.54 | 0.22 | 1.16 | 2.05 | 0.003 |
| Model 2a: *Demographics* | 1.71 | 0.24 | 1.31 | 2.24 | <0.001 |
| Model 2b: *Genetics* | 1.93 | 0.26 | 1.48 | 2.52 | <0.001 |
| Model 3a: Model 3 + *Medication* | 1.54 | 0.22 | 1.16 | 2.05 | 0.003 |
| Model 3b: Model 3 + *Physical Activity* | 1.49 | 0.22 | 1.12 | 1.98 | 0.006 |
| Model 3c: Model 3 + *Body Mass Index* | 1.46 | 0.22 | 1.10 | 1.95 | 0.010 |
| Model 3d: Model 3 + *Fully Adjusted* [c] | 1.43 | 0.21 | 1.07 | 1.90 | 0.016 |
| Model 4a: Model 2 + *Health* | 1.72 | 0.24 | 1.31 | 2.26 | <0.001 |
| Model 4b: Model 3 + *Health* | 1.52 | 0.22 | 1.15 | 2.03 | 0.004 |
| Model 4c: Model 3 + *Fully Adjusted* [d] | 1.42 | 0.21 | 1.06 | 1.89 | 0.018 |

**Notes:** The *low-risk* group is the reference; HR = hazard ratio; SE = standard error; CI = confidence interval; *p* = significance value.

a *Demographic and genetic variables*: age; sex; 10 principal components (PCs); C-reactive Protein (CRP) polygenic score (PGS); White Blood Cell Counts (WBCC) PGS; Insulin Growth Factor-1 (IGF-1) PGS; Anxiety PGS; Depression PGS; Schizophrenia PGS; Insomnia PGS; Pain PGS.

b All variables: age; sex; 10 PCs; CRP PGS; WBCC PGS; IGF-1 PGS; PGS; Anxiety PGS; Depression PGS; Schizophrenia PGS; Insomnia PGS; Pain PGS; education; wealth; smoking status; alcohol consumption; mobility.

c Additional variables: medication, physical activity; BMI.

d Additional variables: health (i.e., chronic lung disease; coronary heart disease; abnormal heart rhythm; heart murmur; congestive heart failure; angina; hypertension; diabetes; cancer; Parkinson's; Alzheimer's; dementia; asthma; arthritis; osteoporosis; psychiatric disorder).

***Table S9ai.*** **Longitudinal associations between insulin growth factor-1 and hospitalisation for diseases of the genitourinary system (*n*=3,524)**

| **Adjustments** | **IGF-1** | | | | |
|---|---|---|---|---|---|
| | **HR** | **SE** | **95% CI** | | ***p*** |
| **Genitourinary Disorders** | | | | | |
| Model 1: *Unadjusted* | 0.71 | 0.07 | 0.58 | 0.86 | 0.001 |
| Model 2: *Model 1 + demographics & genetics* [a] | 1.03 | 0.11 | 0.84 | 1.26 | 0.789 |
| Model 3: *Model 2 + Fully Adjusted* [b] | 1.04 | 0.11 | 0.85 | 1.27 | 0.732 |
| Model 2a: *Demographics* | 1.03 | 0.11 | 0.84 | 1.27 | 0.760 |
| Model 2b: *Genetics* | 0.70 | 0.07 | 0.58 | 0.86 | 0.001 |
| Model 3a: Model 3 + *Medication* | 1.04 | 0.11 | 0.85 | 1.27 | 0.734 |
| Model 3b: Model 3 + *Physical Activity* | 1.05 | 0.11 | 0.86 | 1.29 | 0.617 |
| Model 3c: Model 3 + *Body Mass Index* | 1.05 | 0.11 | 0.85 | 1.28 | 0.668 |
| Model 3d: Model 3 + *Fully Adjusted* [c] | 1.06 | 0.11 | 0.87 | 1.30 | 0.558 |
| Model 4a: Model 2 + *Health* | 1.03 | 0.11 | 0.84 | 1.26 | 0.789 |
| Model 4b: Model 3 + *Health* | 1.04 | 0.11 | 0.85 | 1.27 | 0.733 |
| Model 4c: Model 3 + *Fully Adjusted* [d] | 1.06 | 0.11 | 0.87 | 1.30 | 0.557 |

**Notes:** The *low-risk* group is the reference; HR = hazard ratio; SE = standard error; CI = confidence interval; *p* = significance value.

a *Demographic and genetic variables*: age; sex; 10 principal components (PCs); C-reactive Protein (CRP) polygenic score (PGS); White Blood Cell Counts (WBCC) PGS; Insulin Growth Factor-1 (IGF-1) PGS; Anxiety PGS; Depression PGS; Schizophrenia PGS; Insomnia PGS; Pain PGS.

b All variables: age; sex; 10 PCs; CRP PGS; WBCC PGS; IGF-1 PGS; PGS; Anxiety PGS; Depression PGS; Schizophrenia PGS; Insomnia PGS; Pain PGS; education; wealth; smoking status; alcohol consumption; mobility.

c Additional variables: medication, physical activity; BMI.

d Additional variables: health (i.e., chronic lung disease; coronary heart disease; abnormal heart rhythm; heart murmur; congestive heart failure; angina; hypertension; diabetes; cancer; Parkinson's; Alzheimer's; dementia; asthma; arthritis; osteoporosis; psychiatric disorder).

***Table S9aj.*** **Longitudinal associations between immune and neuroendocrine profiles and hospitalisation for infectious and parasitic diseases (*n*=4,118)**

| Adjustments | Immune and Neuroendocrine Profiles | | | | |
|---|---|---|---|---|---|
| | HR | SE | 95% CI | | *p* |
| ***Moderate-risk* Profile \| Infections** | | | | | |
| Model 1: *Unadjusted* | 1.29 | 0.12 | 1.07 | 1.54 | 0.007 |
| Model 2: *Model 1 + demographics & genetics* [a] | 1.11 | 0.10 | 0.92 | 1.33 | 0.287 |
| Model 3: *Model 2 + Fully Adjusted* [b] | 1.02 | 0.10 | 0.85 | 1.23 | 0.810 |
| Model 2a: *Demographics* | 1.12 | 0.11 | 0.93 | 1.35 | 0.225 |
| Model 2b: *Genetics* | 1.27 | 0.12 | 1.06 | 1.53 | 0.011 |
| Model 3a: Model 3 + *Medication* | 1.02 | 0.10 | 0.85 | 1.23 | 0.824 |
| Model 3b: Model 3 + *Physical Activity* | 1.00 | 0.10 | 0.83 | 1.21 | 0.978 |
| Model 3c: Model 3 + *Body Mass Index* | 1.04 | 0.10 | 0.85 | 1.26 | 0.717 |
| Model 3d: Model 3 + *Fully Adjusted* [c] | 1.02 | 0.10 | 0.84 | 1.24 | 0.842 |
| Model 4a: Model 2 + *Health* | 1.11 | 0.10 | 0.92 | 1.33 | 0.287 |
| Model 4b: Model 3 + *Health* | 1.02 | 0.10 | 0.84 | 1.23 | 0.853 |
| Model 4c: Model 3 + *Fully Adjusted* [d] | 1.02 | 0.10 | 0.84 | 1.23 | 0.877 |
| ***High-risk* Profile \| Infections** | | | | | |
| Model 1: *Unadjusted* | 1.78 | 0.22 | 1.39 | 2.27 | <0.001 |
| Model 2: *Model 1 + demographics & genetics* [a] | 1.55 | 0.20 | 1.21 | 1.98 | 0.001 |
| Model 3: *Model 2 + Fully Adjusted* [b] | 1.38 | 0.18 | 1.08 | 1.78 | 0.011 |
| Model 2a: *Demographics* | 1.57 | 0.20 | 1.23 | 2.01 | <0.001 |
| Model 2b: *Genetics* | 1.77 | 0.22 | 1.39 | 2.26 | <0.001 |
| Model 3a: Model 3 + *Medication* | 1.39 | 0.18 | 1.08 | 1.78 | 0.011 |
| Model 3b: Model 3 + *Physical Activity* | 1.31 | 0.17 | 1.02 | 1.69 | 0.034 |
| Model 3c: Model 3 + *Body Mass Index* | 1.40 | 0.18 | 1.09 | 1.81 | 0.010 |
| Model 3d: Model 3 + *Fully Adjusted* [c] | 1.34 | 0.18 | 1.04 | 1.74 | 0.025 |
| Model 4a: Model 2 + *Health* | 1.55 | 0.20 | 1.21 | 1.98 | 0.001 |
| Model 4b: Model 3 + *Health* | 1.38 | 0.18 | 1.07 | 1.77 | 0.012 |
| Model 4c: Model 3 + *Fully Adjusted* [d] | 1.34 | 0.18 | 1.04 | 1.74 | 0.025 |

**Notes:** The *low-risk* group is the reference; HR = hazard ratio; SE = standard error; CI = confidence interval; *p* = significance value.

a *Demographic and genetic variables*: age; sex; 10 principal components (PCs); C-reactive Protein (CRP) polygenic score (PGS); White Blood Cell Counts (WBCC) PGS; Insulin Growth Factor-1 (IGF-1) PGS; Anxiety PGS; Depression PGS; Schizophrenia PGS; Insomnia PGS; Pain PGS.

b All variables: age; sex; 10 PCs; CRP PGS; WBCC PGS; IGF-1 PGS; PGS; Anxiety PGS; Depression PGS; Schizophrenia PGS; Insomnia PGS; Pain PGS; education; wealth; smoking status; alcohol consumption; mobility.

c Additional variables: medication, physical activity; BMI.

d Additional variables: health (i.e., chronic lung disease; coronary heart disease; abnormal heart rhythm; heart murmur; congestive heart failure; angina; hypertension; diabetes; cancer; Parkinson's; Alzheimer's; dementia; asthma; arthritis; osteoporosis; psychiatric disorder).

***Table S9ak.*** **Longitudinal associations between C-reactive protein and hospitalisation for infectious and parasitic diseases (*n*=4,118)**

| Adjustments | CRP | | | | |
|---|---|---|---|---|---|
| | HR | SE | 95% CI | | *p* |
| **Infections** | | | | | |
| Model 1: *Unadjusted* | 1.31 | 0.08 | 1.17 | 1.48 | <0.001 |
| Model 2: *Model 1 + demographics & genetics* [a] | 1.20 | 0.08 | 1.06 | 1.36 | 0.004 |
| Model 3: *Model 2 + Fully Adjusted* [b] | 1.12 | 0.07 | 0.99 | 1.27 | 0.076 |
| Model 2a: *Demographics* | 1.21 | 0.08 | 1.07 | 1.37 | 0.002 |
| Model 2b: *Genetics* | 1.31 | 0.08 | 1.16 | 1.47 | <0.001 |
| Model 3a: Model 3 + *Medication* | 1.12 | 0.07 | 0.99 | 1.28 | 0.074 |
| Model 3b: Model 3 + *Physical Activity* | 1.09 | 0.07 | 0.96 | 1.24 | 0.184 |
| Model 3c: Model 3 + *Body Mass Index* | 1.14 | 0.08 | 1.00 | 1.30 | 0.059 |
| Model 3d: Model 3 + *Fully Adjusted* [c] | 1.11 | 0.08 | 0.97 | 1.27 | 0.127 |
| Model 4a: Model 2 + *Health* | 1.20 | 0.08 | 1.06 | 1.36 | 0.004 |
| Model 4b: Model 3 + *Health* | 1.12 | 0.07 | 0.98 | 1.27 | 0.087 |
| Model 4c: Model 3 + *Fully Adjusted* [d] | 1.11 | 0.08 | 0.97 | 1.26 | 0.140 |

**Notes:** The *low-risk* group is the reference; HR = hazard ratio; SE = standard error; CI = confidence interval; *p* = significance value.

a *Demographic and genetic variables*: age; sex; 10 principal components (PCs); C-reactive Protein (CRP) polygenic score (PGS); White Blood Cell Counts (WBCC) PGS; Insulin Growth Factor-1 (IGF-1) PGS; Anxiety PGS; Depression PGS; Schizophrenia PGS; Insomnia PGS; Pain PGS.

b All variables: age; sex; 10 PCs; CRP PGS; WBCC PGS; IGF-1 PGS; PGS; Anxiety PGS; Depression PGS; Schizophrenia PGS; Insomnia PGS; Pain PGS; education; wealth; smoking status; alcohol consumption; mobility.

c Additional variables: medication, physical activity; BMI.

d Additional variables: health (i.e., chronic lung disease; coronary heart disease; abnormal heart rhythm; heart murmur; congestive heart failure; angina; hypertension; diabetes; cancer; Parkinson's; Alzheimer's; dementia; asthma; arthritis; osteoporosis; psychiatric disorder).

***Table S9al.*** **Longitudinal associations between fibrinogen and hospitalisation for infectious and parasitic diseases (*n*=4,118)**

| **Adjustments** | **Fb** | | | | |
|---|---|---|---|---|---|
| | **HR** | **SE** | **95% CI** | | ***p*** |
| **Infections** | | | | | |
| Model 1: *Unadjusted* | 1.34 | 0.10 | 1.16 | 1.55 | <0.001 |
| Model 2: *Model 1 + demographics & genetics* [a] | 1.22 | 0.10 | 1.05 | 1.42 | 0.011 |
| Model 3: *Model 2 + Fully Adjusted* [b] | 1.13 | 0.09 | 0.97 | 1.32 | 0.111 |
| Model 2a: *Demographics* | 1.23 | 0.10 | 1.06 | 1.43 | 0.008 |
| Model 2b: *Genetics* | 1.34 | 0.10 | 1.15 | 1.55 | <0.001 |
| Model 3a: Model 3 + *Medication* | 1.14 | 0.09 | 0.97 | 1.33 | 0.110 |
| Model 3b: Model 3 + *Physical Activity* | 1.11 | 0.09 | 0.96 | 1.30 | 0.170 |
| Model 3c: Model 3 + *Body Mass Index* | 1.14 | 0.09 | 0.97 | 1.33 | 0.104 |
| Model 3d: Model 3 + *Fully Adjusted* [c] | 1.12 | 0.09 | 0.96 | 1.31 | 0.149 |
| Model 4a: Model 2 + *Health* | 1.21 | 0.09 | 1.04 | 1.41 | 0.013 |
| Model 4b: Model 3 + *Health* | 1.14 | 0.09 | 0.97 | 1.33 | 0.107 |
| Model 4c: Model 3 + *Fully Adjusted* [d] | 1.12 | 0.09 | 0.96 | 1.31 | 0.144 |

**Notes:** The *low-risk* group is the reference; HR = hazard ratio; SE = standard error; CI = confidence interval; *p* = significance value.

a *Demographic and genetic variables*: age; sex; 10 principal components (PCs); C-reactive Protein (CRP) polygenic score (PGS); White Blood Cell Counts (WBCC) PGS; Insulin Growth Factor-1 (IGF-1) PGS; Anxiety PGS; Depression PGS; Schizophrenia PGS; Insomnia PGS; Pain PGS.

b All variables: age; sex; 10 PCs; CRP PGS; WBCC PGS; IGF-1 PGS; PGS; Anxiety PGS; Depression PGS; Schizophrenia PGS; Insomnia PGS; Pain PGS; education; wealth; smoking status; alcohol consumption; mobility.

c Additional variables: medication, physical activity; BMI.

d Additional variables: health (i.e., chronic lung disease; coronary heart disease; abnormal heart rhythm; heart murmur; congestive heart failure; angina; hypertension; diabetes; cancer; Parkinson's; Alzheimer's; dementia; asthma; arthritis; osteoporosis; psychiatric disorder).

***Table S9am.*** **Longitudinal associations between white blood cell counts and hospitalisation for infectious and parasitic diseases (*n*=4,118)**

| Adjustments | WBCC | | | | |
|---|---|---|---|---|---|
| | HR | SE | 95% CI | | *p* |
| **Infections** | | | | | |
| Model 1: *Unadjusted* | 1.77 | 0.31 | 1.26 | 2.48 | 0.001 |
| Model 2: *Model 1 + demographics & genetics* [a] | 1.63 | 0.29 | 1.14 | 2.31 | 0.007 |
| Model 3: *Model 2 + Fully Adjusted* [b] | 1.37 | 0.26 | 0.95 | 1.97 | 0.093 |
| Model 2a: *Demographics* | 1.58 | 0.28 | 1.12 | 2.23 | 0.010 |
| Model 2b: *Genetics* | 1.78 | 0.31 | 1.27 | 2.51 | 0.001 |
| Model 3a: Model 3 + *Medication* | 1.37 | 0.26 | 0.95 | 1.98 | 0.092 |
| Model 3b: Model 3 + *Physical Activity* | 1.33 | 0.25 | 0.92 | 1.91 | 0.127 |
| Model 3c: Model 3 + *Body Mass Index* | 1.38 | 0.26 | 0.95 | 2.00 | 0.087 |
| Model 3d: Model 3 + *Fully Adjusted* [c] | 1.35 | 0.25 | 0.94 | 1.96 | 0.108 |
| Model 4a: Model 2 + *Health* | 1.59 | 0.29 | 1.12 | 2.26 | 0.010 |
| Model 4b: Model 3 + *Health* | 1.36 | 0.25 | 0.94 | 1.96 | 0.102 |
| Model 4c: Model 3 + *Fully Adjusted* [d] | 1.35 | 0.25 | 0.93 | 1.95 | 0.114 |

**Notes:** The *low-risk* group is the reference; HR = hazard ratio; SE = standard error; CI = confidence interval; *p* = significance value.

a *Demographic and genetic variables*: age; sex; 10 principal components (PCs); C-reactive Protein (CRP) polygenic score (PGS); White Blood Cell Counts (WBCC) PGS; Insulin Growth Factor-1 (IGF-1) PGS; Anxiety PGS; Depression PGS; Schizophrenia PGS; Insomnia PGS; Pain PGS.

b All variables: age; sex; 10 PCs; CRP PGS; WBCC PGS; IGF-1 PGS; PGS; Anxiety PGS; Depression PGS; Schizophrenia PGS; Insomnia PGS; Pain PGS; education; wealth; smoking status; alcohol consumption; mobility.

c Additional variables: medication, physical activity; BMI.

d Additional variables: health (i.e., chronic lung disease; coronary heart disease; abnormal heart rhythm; heart murmur; congestive heart failure; angina; hypertension; diabetes; cancer; Parkinson's; Alzheimer's; dementia; asthma; arthritis; osteoporosis; psychiatric disorder).

***Table S9an.*** **Longitudinal associations between insulin growth factor-1 and hospitalisation for infectious and parasitic diseases (*n*=4,118)**

| Adjustments | IGF-1 | | | | |
|---|---|---|---|---|---|
| | HR | SE | 95% CI | | *p* |
| **Infections** | | | | | |
| Model 1: *Unadjusted* | 0.70 | 0.09 | 0.54 | 0.90 | 0.005 |
| Model 2: *Model 1 + demographics & genetics* [a] | 1.02 | 0.14 | 0.78 | 1.32 | 0.914 |
| Model 3: *Model 2 + Fully Adjusted* [b] | 1.04 | 0.14 | 0.80 | 1.35 | 0.761 |
| Model 2a: *Demographics* | 0.99 | 0.13 | 0.77 | 1.29 | 0.963 |
| Model 2b: *Genetics* | 0.70 | 0.09 | 0.55 | 0.91 | 0.007 |
| Model 3a: Model 3 + *Medication* | 1.04 | 0.14 | 0.80 | 1.35 | 0.763 |
| Model 3b: Model 3 + *Physical Activity* | 1.05 | 0.14 | 0.82 | 1.36 | 0.688 |
| Model 3c: Model 3 + *Body Mass Index* | 1.04 | 0.14 | 0.80 | 1.35 | 0.763 |
| Model 3d: Model 3 + *Fully Adjusted* [c] | 1.05 | 0.14 | 0.81 | 1.36 | 0.699 |
| Model 4a: Model 2 + *Health* | 1.02 | 0.14 | 0.78 | 1.32 | 0.891 |
| Model 4b: Model 3 + *Health* | 1.04 | 0.14 | 0.81 | 1.35 | 0.747 |
| Model 4c: Model 3 + *Fully Adjusted* [d] | 1.06 | 0.14 | 0.82 | 1.36 | 0.683 |

**Notes:** The *low-risk* group is the reference; HR = hazard ratio; SE = standard error; CI = confidence interval; *p* = significance value.

a *Demographic and genetic variables*: age; sex; 10 principal components (PCs); C-reactive Protein (CRP) polygenic score (PGS); White Blood Cell Counts (WBCC) PGS; Insulin Growth Factor-1 (IGF-1) PGS; Anxiety PGS; Depression PGS; Schizophrenia PGS; Insomnia PGS; Pain PGS.

b All variables: age; sex; 10 PCs; CRP PGS; WBCC PGS; IGF-1 PGS; PGS; Anxiety PGS; Depression PGS; Schizophrenia PGS; Insomnia PGS; Pain PGS; education; wealth; smoking status; alcohol consumption; mobility.

c Additional variables: medication, physical activity; BMI.

d Additional variables: health (i.e., chronic lung disease; coronary heart disease; abnormal heart rhythm; heart murmur; congestive heart failure; angina; hypertension; diabetes; cancer; Parkinson's; Alzheimer's; dementia; asthma; arthritis; osteoporosis; psychiatric disorder).

*Table S9ao.* **Longitudinal associations between immune and neuroendocrine profiles and hospitalisation for diseases of the musculoskeletal system and connective tissue (*n*=3,524)**

| Adjustments | Immune and Neuroendocrine Profiles | | | | |
|---|---|---|---|---|---|
| | HR | SE | 95% CI | | *p* |
| ***Moderate-risk* Profile \| Musculoskeletal Disorders** | | | | | |
| Model 1: *Unadjusted* | 1.36 | 0.09 | 1.20 | 1.55 | <0.001 |
| Model 2: *Model 1 + demographics & genetics* [a] | 1.22 | 0.08 | 1.07 | 1.39 | 0.003 |
| Model 3: *Model 2 + Fully Adjusted* [b] | 1.13 | 0.08 | 0.99 | 1.30 | 0.063 |
| Model 2a: *Demographics* | 1.20 | 0.08 | 1.05 | 1.36 | 0.007 |
| Model 2b: *Genetics* | 1.36 | 0.09 | 1.20 | 1.55 | <0.001 |
| Model 3a: Model 3 + *Medication* | 1.14 | 0.08 | 0.99 | 1.30 | 0.062 |
| Model 3b: Model 3 + *Physical Activity* | 1.12 | 0.08 | 0.98 | 1.28 | 0.087 |
| Model 3c: Model 3 + *Body Mass Index* | 1.11 | 0.08 | 0.97 | 1.27 | 0.136 |
| Model 3d: Model 3 + *Fully Adjusted* [c] | 1.10 | 0.08 | 0.96 | 1.26 | 0.167 |
| Model 4a: Model 2 + *Health* | 1.19 | 0.08 | 1.05 | 1.36 | 0.008 |
| Model 4b: Model 3 + *Health* | 1.12 | 0.08 | 0.98 | 1.28 | 0.092 |
| Model 4c: Model 3 + *Fully Adjusted* [d] | 1.09 | 0.08 | 0.95 | 1.26 | 0.201 |
| ***High-risk* Profile \| Musculoskeletal Disorders** | | | | | |
| Model 1: *Unadjusted* | 1.49 | 0.14 | 1.24 | 1.80 | <0.001 |
| Model 2: *Model 1 + demographics & genetics* [a] | 1.41 | 0.14 | 1.17 | 1.70 | <0.001 |
| Model 3: *Model 2 + Fully Adjusted* [b] | 1.28 | 0.13 | 1.06 | 1.55 | 0.012 |
| Model 2a: *Demographics* | 1.39 | 0.13 | 1.15 | 1.68 | 0.001 |
| Model 2b: *Genetics* | 1.57 | 0.15 | 1.30 | 1.89 | <0.001 |
| Model 3a: Model 3 + *Medication* | 1.28 | 0.13 | 1.06 | 1.55 | 0.012 |
| Model 3b: Model 3 + *Physical Activity* | 1.25 | 0.12 | 1.03 | 1.51 | 0.025 |
| Model 3c: Model 3 + *Body Mass Index* | 1.25 | 0.13 | 1.03 | 1.52 | 0.027 |
| Model 3d: Model 3 + *Fully Adjusted* [c] | 1.22 | 0.12 | 1.00 | 1.48 | 0.052 |
| Model 4a: Model 2 + *Health* | 1.37 | 0.13 | 1.13 | 1.65 | 0.001 |
| Model 4b: Model 3 + *Health* | 1.26 | 0.12 | 1.04 | 1.52 | 0.020 |
| Model 4c: Model 3 + *Fully Adjusted* [d] | 1.20 | 0.12 | 0.99 | 1.46 | 0.069 |

**Notes:** The *low-risk* group is the reference; HR = hazard ratio; SE = standard error; CI = confidence interval; *p* = significance value.

a *Demographic and genetic variables*: age; sex; 10 principal components (PCs); C-reactive Protein (CRP) polygenic score (PGS); White Blood Cell Counts (WBCC) PGS; Insulin Growth Factor-1 (IGF-1) PGS; Anxiety PGS; Depression PGS; Schizophrenia PGS; Insomnia PGS; Pain PGS.

b All variables: age; sex; 10 PCs; CRP PGS; WBCC PGS; IGF-1 PGS; PGS; Anxiety PGS; Depression PGS; Schizophrenia PGS; Insomnia PGS; Pain PGS; education; wealth; smoking status; alcohol consumption; mobility.

c Additional variables: medication, physical activity; BMI.

d Additional variables: health (i.e., chronic lung disease; coronary heart disease; abnormal heart rhythm; heart murmur; congestive heart failure; angina; hypertension; diabetes; cancer; Parkinson's; Alzheimer's; dementia; asthma; arthritis; osteoporosis; psychiatric disorder).

***Table S9ap.*** **Longitudinal associations between C-reactive protein and hospitalisation for diseases of the musculoskeletal system and connective tissue (*n*=3,524)**

| Adjustments | CRP | | | | |
|---|---|---|---|---|---|
| | HR | SE | 95% CI | | *p* |
| **Musculoskeletal Disorders** | | | | | |
| Model 1: *Unadjusted* | 1.23 | 0.06 | 1.13 | 1.35 | <0.001 |
| Model 2: *Model 1 + demographics & genetics* [a] | 1.26 | 0.06 | 1.15 | 1.37 | <0.001 |
| Model 3: *Model 2 + Fully Adjusted* [b] | 1.18 | 0.06 | 1.08 | 1.29 | <0.001 |
| Model 2a: *Demographics* | 1.24 | 0.06 | 1.13 | 1.35 | <0.001 |
| Model 2b: *Genetics* | 1.34 | 0.06 | 1.23 | 1.47 | <0.001 |
| Model 3a: Model 3 + *Medication* | 1.18 | 0.06 | 1.08 | 1.29 | <0.001 |
| Model 3b: Model 3 + *Physical Activity* | 1.17 | 0.05 | 1.06 | 1.28 | 0.001 |
| Model 3c: Model 3 + *Body Mass Index* | 1.17 | 0.06 | 1.06 | 1.28 | 0.002 |
| Model 3d: Model 3 + *Fully Adjusted* [c] | 1.15 | 0.06 | 1.05 | 1.27 | 0.004 |
| Model 4a: Model 2 + *Health* | 1.23 | 0.06 | 1.13 | 1.35 | <0.001 |
| Model 4b: Model 3 + *Health* | 1.17 | 0.06 | 1.07 | 1.28 | 0.001 |
| Model 4c: Model 3 + *Fully Adjusted* [d] | 1.14 | 0.06 | 1.04 | 1.26 | 0.007 |

**Notes:** The *low-risk* group is the reference; HR = hazard ratio; SE = standard error; CI = confidence interval; *p* = significance value.

a *Demographic and genetic variables*: age; sex; 10 principal components (PCs); C-reactive Protein (CRP) polygenic score (PGS); White Blood Cell Counts (WBCC) PGS; Insulin Growth Factor-1 (IGF-1) PGS; Anxiety PGS; Depression PGS; Schizophrenia PGS; Insomnia PGS; Pain PGS.

b All variables: age; sex; 10 PCs; CRP PGS; WBCC PGS; IGF-1 PGS; PGS; Anxiety PGS; Depression PGS; Schizophrenia PGS; Insomnia PGS; Pain PGS; education; wealth; smoking status; alcohol consumption; mobility.

c Additional variables: medication, physical activity; BMI.

d Additional variables: health (i.e., chronic lung disease; coronary heart disease; abnormal heart rhythm; heart murmur; congestive heart failure; angina; hypertension; diabetes; cancer; Parkinson's; Alzheimer's; dementia; asthma; arthritis; osteoporosis; psychiatric disorder).

*Table S9aq.* **Longitudinal associations between fibrinogen and hospitalisation for diseases of the musculoskeletal system and connective tissue (*n*=3,524)**

| Adjustments | Fb | | | | |
|---|---|---|---|---|---|
| | HR | SE | 95% CI | | *p* |
| **Musculoskeletal Disorders** | | | | | |
| Model 1: *Unadjusted* | 1.24 | 0.07 | 1.12 | 1.38 | <0.001 |
| Model 2: *Model 1 + demographics & genetics* [a] | 1.12 | 0.06 | 1.01 | 1.26 | 0.040 |
| Model 3: *Model 2 + Fully Adjusted* [b] | 1.07 | 0.06 | 0.95 | 1.19 | 0.273 |
| Model 2a: *Demographics* | 1.12 | 0.06 | 1.00 | 1.25 | 0.042 |
| Model 2b: *Genetics* | 1.23 | 0.07 | 1.10 | 1.37 | <0.001 |
| Model 3a: Model 3 + *Medication* | 1.07 | 0.06 | 0.95 | 1.19 | 0.273 |
| Model 3b: Model 3 + *Physical Activity* | 1.06 | 0.06 | 0.95 | 1.18 | 0.335 |
| Model 3c: Model 3 + *Body Mass Index* | 1.05 | 0.06 | 0.94 | 1.18 | 0.415 |
| Model 3d: Model 3 + *Fully Adjusted* [c] | 1.04 | 0.06 | 0.93 | 1.17 | 0.479 |
| Model 4a: Model 2 + *Health* | 1.11 | 0.06 | 0.99 | 1.24 | 0.070 |
| Model 4b: Model 3 + *Health* | 1.06 | 0.06 | 0.95 | 1.19 | 0.308 |
| Model 4c: Model 3 + *Fully Adjusted* [d] | 1.04 | 0.06 | 0.93 | 1.16 | 0.505 |

**Notes:** The *low-risk* group is the reference; HR = hazard ratio; SE = standard error; CI = confidence interval; *p* = significance value.

a *Demographic and genetic variables*: age; sex; 10 principal components (PCs); C-reactive Protein (CRP) polygenic score (PGS); White Blood Cell Counts (WBCC) PGS; Insulin Growth Factor-1 (IGF-1) PGS; Anxiety PGS; Depression PGS; Schizophrenia PGS; Insomnia PGS; Pain PGS.

b All variables: age; sex; 10 PCs; CRP PGS; WBCC PGS; IGF-1 PGS; PGS; Anxiety PGS; Depression PGS; Schizophrenia PGS; Insomnia PGS; Pain PGS; education; wealth; smoking status; alcohol consumption; mobility.

c Additional variables: medication, physical activity; BMI.

d Additional variables: health (i.e., chronic lung disease; coronary heart disease; abnormal heart rhythm; heart murmur; congestive heart failure; angina; hypertension; diabetes; cancer; Parkinson's; Alzheimer's; dementia; asthma; arthritis; osteoporosis; psychiatric disorder).

***Table S9ar.*** **Longitudinal associations between white blood cell counts and hospitalisation for diseases of the musculoskeletal system and connective tissue (*n*=3,524)**

| Adjustments | WBCC | | | | |
|---|---|---|---|---|---|
| | **HR** | **SE** | **95% CI** | | ***p*** |
| **Musculoskeletal Disorders** | | | | | |
| Model 1: *Unadjusted* | 1.29 | 0.16 | 1.01 | 1.64 | 0.039 |
| Model 2: *Model 1 + demographics & genetics* [a] | 1.29 | 0.16 | 1.01 | 1.66 | 0.042 |
| Model 3: *Model 2 + Fully Adjusted* [b] | 1.19 | 0.16 | 0.92 | 1.54 | 0.197 |
| Model 2a: *Demographics* | 1.26 | 0.16 | 0.99 | 1.61 | 0.066 |
| Model 2b: *Genetics* | 1.28 | 0.16 | 1.00 | 1.63 | 0.048 |
| Model 3a: Model 3 + *Medication* | 1.19 | 0.16 | 0.92 | 1.54 | 0.196 |
| Model 3b: Model 3 + *Physical Activity* | 1.16 | 0.15 | 0.89 | 1.50 | 0.266 |
| Model 3c: Model 3 + *Body Mass Index* | 1.15 | 0.15 | 0.88 | 1.49 | 0.308 |
| Model 3d: Model 3 + *Fully Adjusted* [c] | 1.12 | 0.15 | 0.86 | 1.46 | 0.384 |
| Model 4a: Model 2 + *Health* | 1.25 | 0.16 | 0.98 | 1.60 | 0.076 |
| Model 4b: Model 3 + *Health* | 1.16 | 0.15 | 0.89 | 1.50 | 0.271 |
| Model 4c: Model 3 + *Fully Adjusted* [d] | 1.11 | 0.15 | 0.85 | 1.44 | 0.453 |

**Notes:** The *low-risk* group is the reference; HR = hazard ratio; SE = standard error; CI = confidence interval; *p* = significance value.

a *Demographic and genetic variables*: age; sex; 10 principal components (PCs); C-reactive Protein (CRP) polygenic score (PGS); White Blood Cell Counts (WBCC) PGS; Insulin Growth Factor-1 (IGF-1) PGS; Anxiety PGS; Depression PGS; Schizophrenia PGS; Insomnia PGS; Pain PGS.

b All variables: age; sex; 10 PCs; CRP PGS; WBCC PGS; IGF-1 PGS; PGS; Anxiety PGS; Depression PGS; Schizophrenia PGS; Insomnia PGS; Pain PGS; education; wealth; smoking status; alcohol consumption; mobility.

c Additional variables: medication, physical activity; BMI.

d Additional variables: health (i.e., chronic lung disease; coronary heart disease; abnormal heart rhythm; heart murmur; congestive heart failure; angina; hypertension; diabetes; cancer; Parkinson's; Alzheimer's; dementia; asthma; arthritis; osteoporosis; psychiatric disorder).

***Table S9as.*** **Longitudinal associations between insulin growth factor-1 and hospitalisation for diseases of the musculoskeletal system and connective tissue (*n*=3,524)**

| Adjustments | IGF-1 | | | | |
|---|---|---|---|---|---|
| | HR | SE | 95% CI | | *p* |
| **Musculoskeletal Disorders** | | | | | |
| Model 1: *Unadjusted* | 0.69 | 0.06 | 0.57 | 0.82 | <0.001 |
| Model 2: *Model 1 + demographics & genetics* [a] | 0.94 | 0.09 | 0.78 | 1.14 | 0.554 |
| Model 3: *Model 2 + Fully Adjusted* [b] | 0.95 | 0.09 | 0.79 | 1.15 | 0.617 |
| Model 2a: *Demographics* | 0.95 | 0.09 | 0.79 | 1.14 | 0.570 |
| Model 2b: *Genetics* | 0.69 | 0.07 | 0.57 | 0.83 | <0.001 |
| Model 3a: Model 3 + *Medication* | 0.95 | 0.09 | 0.79 | 1.15 | 0.617 |
| Model 3b: Model 3 + *Physical Activity* | 0.95 | 0.09 | 0.79 | 1.15 | 0.609 |
| Model 3c: Model 3 + *Body Mass Index* | 0.96 | 0.09 | 0.79 | 1.15 | 0.633 |
| Model 3d: Model 3 + *Fully Adjusted* [c] | 0.96 | 0.09 | 0.79 | 1.15 | 0.633 |
| Model 4a: Model 2 + *Health* | 0.95 | 0.09 | 0.79 | 1.15 | 0.582 |
| Model 4b: Model 3 + *Health* | 0.95 | 0.09 | 0.79 | 1.15 | 0.623 |
| Model 4c: Model 3 + *Fully Adjusted* [d] | 0.96 | 0.09 | 0.79 | 1.15 | 0.632 |

**Notes:** The *low-risk* group is the reference; HR = hazard ratio; SE = standard error; CI = confidence interval; *p* = significance value.

a *Demographic and genetic variables*: age; sex; 10 principal components (PCs); C-reactive Protein (CRP) polygenic score (PGS); White Blood Cell Counts (WBCC) PGS; Insulin Growth Factor-1 (IGF-1) PGS; Anxiety PGS; Depression PGS; Schizophrenia PGS; Insomnia PGS; Pain PGS.

b All variables: age; sex; 10 PCs; CRP PGS; WBCC PGS; IGF-1 PGS; PGS; Anxiety PGS; Depression PGS; Schizophrenia PGS; Insomnia PGS; Pain PGS; education; wealth; smoking status; alcohol consumption; mobility.

c Additional variables: medication, physical activity; BMI.

d Additional variables: health (i.e., chronic lung disease; coronary heart disease; abnormal heart rhythm; heart murmur; congestive heart failure; angina; hypertension; diabetes; cancer; Parkinson's; Alzheimer's; dementia; asthma; arthritis; osteoporosis; psychiatric disorder).

*Table S9at.* **Longitudinal associations between immune and neuroendocrine profiles and hospitalisation for diseases of the nervous system (*n*=4,057)**

| Adjustments | Immune and Neuroendocrine Profiles | | | | |
|---|---|---|---|---|---|
| | HR | SE | 95% CI | | *p* |
| ***Moderate-risk* Profile \| Nervous Disorders** | | | | | |
| Model 1: *Unadjusted* | 1.37 | 0.13 | 1.15 | 1.65 | 0.001 |
| Model 2: *Model 1 + demographics & genetics* [a] | 1.19 | 0.11 | 0.99 | 1.43 | 0.067 |
| Model 3: *Model 2 + Fully Adjusted* [b] | 1.12 | 0.11 | 0.93 | 1.35 | 0.220 |
| Model 2a: *Demographics* | 1.21 | 0.11 | 1.01 | 1.45 | 0.043 |
| Model 2b: *Genetics* | 1.36 | 0.13 | 1.13 | 1.62 | 0.001 |
| Model 3a: Model 3 + *Medication* | 1.12 | 0.11 | 0.93 | 1.35 | 0.220 |
| Model 3b: Model 3 + *Physical Activity* | 1.09 | 0.10 | 0.91 | 1.32 | 0.346 |
| Model 3c: Model 3 + *Body Mass Index* | 1.08 | 0.11 | 0.89 | 1.31 | 0.428 |
| Model 3d: Model 3 + *Fully Adjusted* [c] | 1.06 | 0.10 | 0.88 | 1.29 | 0.549 |
| Model 4a: Model 2 + *Health* | 1.19 | 0.11 | 0.99 | 1.43 | 0.067 |
| Model 4b: Model 3 + *Health* | 1.11 | 0.11 | 0.92 | 1.34 | 0.257 |
| Model 4c: Model 3 + *Fully Adjusted* [d] | 1.05 | 0.10 | 0.87 | 1.28 | 0.594 |
| ***High-risk* Profile \| Nervous Disorders** | | | | | |
| Model 1: *Unadjusted* | 1.24 | 0.18 | 0.94 | 1.64 | 0.125 |
| Model 2: *Model 1 + demographics & genetics* [a] | 1.06 | 0.15 | 0.80 | 1.41 | 0.666 |
| Model 3: *Model 2 + Fully Adjusted* [b] | 0.97 | 0.14 | 0.73 | 1.28 | 0.808 |
| Model 2a: *Demographics* | 1.08 | 0.15 | 0.82 | 1.43 | 0.592 |
| Model 2b: *Genetics* | 1.22 | 0.17 | 0.92 | 1.61 | 0.165 |
| Model 3a: Model 3 + *Medication* | 0.97 | 0.14 | 0.73 | 1.29 | 0.830 |
| Model 3b: Model 3 + *Physical Activity* | 0.90 | 0.13 | 0.68 | 1.20 | 0.486 |
| Model 3c: Model 3 + *Body Mass Index* | 0.93 | 0.14 | 0.69 | 1.24 | 0.598 |
| Model 3d: Model 3 + *Fully Adjusted* [c] | 0.88 | 0.13 | 0.66 | 1.17 | 0.376 |
| Model 4a: Model 2 + *Health* | 1.06 | 0.15 | 0.80 | 1.41 | 0.666 |
| Model 4b: Model 3 + *Health* | 0.95 | 0.14 | 0.71 | 1.26 | 0.721 |
| Model 4c: Model 3 + *Fully Adjusted* [d] | 0.87 | 0.13 | 0.65 | 1.16 | 0.332 |

**Notes:** The *low-risk* group is the reference; HR = hazard ratio; SE = standard error; CI = confidence interval; *p* = significance value.

a *Demographic and genetic variables*: age; sex; 10 principal components (PCs); C-reactive Protein (CRP) polygenic score (PGS); White Blood Cell Counts (WBCC) PGS; Insulin Growth Factor-1 (IGF-1) PGS; Anxiety PGS; Depression PGS; Schizophrenia PGS; Insomnia PGS; Pain PGS.

b All variables: age; sex; 10 PCs; CRP PGS; WBCC PGS; IGF-1 PGS; PGS; Anxiety PGS; Depression PGS; Schizophrenia PGS; Insomnia PGS; Pain PGS; education; wealth; smoking status; alcohol consumption; mobility.

c Additional variables: medication, physical activity; BMI.

d Additional variables: health (i.e., chronic lung disease; coronary heart disease; abnormal heart rhythm; heart murmur; congestive heart failure; angina; hypertension; diabetes; cancer; Parkinson's; Alzheimer's; dementia; asthma; arthritis; osteoporosis; psychiatric disorder).

***Table S9au.*** **Longitudinal associations between C-reactive protein and hospitalisation for diseases of the nervous system (*n*=4,057)**

| Adjustments | CRP | | | | |
|---|---|---|---|---|---|
| | HR | SE | 95% CI | | *p* |
| **Nervous Disorders** | | | | | |
| Model 1: *Unadjusted* | 1.20 | 0.08 | 1.06 | 1.36 | 0.004 |
| Model 2: *Model 1 + demographics & genetics* [a] | 1.09 | 0.07 | 0.95 | 1.23 | 0.213 |
| Model 3: *Model 2 + Fully Adjusted* [b] | 1.03 | 0.07 | 0.91 | 1.18 | 0.630 |
| Model 2a: *Demographics* | 1.10 | 0.07 | 0.97 | 1.25 | 0.150 |
| Model 2b: *Genetics* | 1.19 | 0.08 | 1.05 | 1.34 | 0.007 |
| Model 3a: Model 3 + *Medication* | 1.03 | 0.07 | 0.91 | 1.18 | 0.610 |
| Model 3b: Model 3 + *Physical Activity* | 1.00 | 0.07 | 0.88 | 1.14 | 0.976 |
| Model 3c: Model 3 + *Body Mass Index* | 1.00 | 0.07 | 0.87 | 1.15 | 0.996 |
| Model 3d: Model 3 + *Fully Adjusted* [c] | 0.97 | 0.07 | 0.85 | 1.12 | 0.703 |
| Model 4a: Model 2 + *Health* | 1.09 | 0.07 | 0.95 | 1.23 | 0.213 |
| Model 4b: Model 3 + *Health* | 1.02 | 0.07 | 0.90 | 1.16 | 0.752 |
| Model 4c: Model 3 + *Fully Adjusted* [d] | 0.97 | 0.07 | 0.84 | 1.11 | 0.609 |

**Notes:** The *low-risk* group is the reference; HR = hazard ratio; SE = standard error; CI = confidence interval; *p* = significance value.

a *Demographic and genetic variables*: age; sex; 10 principal components (PCs); C-reactive Protein (CRP) polygenic score (PGS); White Blood Cell Counts (WBCC) PGS; Insulin Growth Factor-1 (IGF-1) PGS; Anxiety PGS; Depression PGS; Schizophrenia PGS; Insomnia PGS; Pain PGS.

b All variables: age; sex; 10 PCs; CRP PGS; WBCC PGS; IGF-1 PGS; PGS; Anxiety PGS; Depression PGS; Schizophrenia PGS; Insomnia PGS; Pain PGS; education; wealth; smoking status; alcohol consumption; mobility.

c Additional variables: medication, physical activity; BMI.

d Additional variables: health (i.e., chronic lung disease; coronary heart disease; abnormal heart rhythm; heart murmur; congestive heart failure; angina; hypertension; diabetes; cancer; Parkinson's; Alzheimer's; dementia; asthma; arthritis; osteoporosis; psychiatric disorder).

***Table S9av.*** **Longitudinal associations between fibrinogen and hospitalisation for diseases of the nervous system (*n*=4,057)**

| Adjustments | Fb | | | | |
|---|---|---|---|---|---|
| | **HR** | **SE** | **95% CI** | | ***p*** |
| **Nervous Disorders** | | | | | |
| Model 1: *Unadjusted* | 1.20 | 0.09 | 1.03 | 1.39 | 0.018 |
| Model 2: *Model 1 + demographics & genetics* [a] | 1.07 | 0.09 | 0.92 | 1.26 | 0.377 |
| Model 3: *Model 2 + Fully Adjusted* [b] | 0.99 | 0.08 | 0.84 | 1.16 | 0.866 |
| Model 2a: *Demographics* | 1.08 | 0.09 | 0.92 | 1.26 | 0.346 |
| Model 2b: *Genetics* | 1.18 | 0.09 | 1.02 | 1.38 | 0.029 |
| Model 3a: Model 3 + *Medication* | 0.99 | 0.08 | 0.84 | 1.16 | 0.880 |
| Model 3b: Model 3 + *Physical Activity* | 0.97 | 0.08 | 0.82 | 1.13 | 0.676 |
| Model 3c: Model 3 + *Body Mass Index* | 0.97 | 0.08 | 0.82 | 1.14 | 0.678 |
| Model 3d: Model 3 + *Fully Adjusted* [c] | 0.95 | 0.08 | 0.81 | 1.12 | 0.560 |
| Model 4a: Model 2 + *Health* | 1.06 | 0.09 | 0.91 | 1.24 | 0.448 |
| Model 4b: Model 3 + *Health* | 0.98 | 0.08 | 0.84 | 1.15 | 0.828 |
| Model 4c: Model 3 + *Fully Adjusted* [d] | 0.95 | 0.08 | 0.81 | 1.12 | 0.534 |

**Notes:** The *low-risk* group is the reference; HR = hazard ratio; SE = standard error; CI = confidence interval; *p* = significance value.

a *Demographic and genetic variables*: age; sex; 10 principal components (PCs); C-reactive Protein (CRP) polygenic score (PGS); White Blood Cell Counts (WBCC) PGS; Insulin Growth Factor-1 (IGF-1) PGS; Anxiety PGS; Depression PGS; Schizophrenia PGS; Insomnia PGS; Pain PGS.

b All variables: age; sex; 10 PCs; CRP PGS; WBCC PGS; IGF-1 PGS; PGS; Anxiety PGS; Depression PGS; Schizophrenia PGS; Insomnia PGS; Pain PGS; education; wealth; smoking status; alcohol consumption; mobility.

c Additional variables: medication, physical activity; BMI.

d Additional variables: health (i.e., chronic lung disease; coronary heart disease; abnormal heart rhythm; heart murmur; congestive heart failure; angina; hypertension; diabetes; cancer; Parkinson's; Alzheimer's; dementia; asthma; arthritis; osteoporosis; psychiatric disorder).

***Table S9aw.*** **Longitudinal associations between white blood cell counts and hospitalisation for diseases of the nervous system (*n*=4,057)**

| Adjustments | WBCC | | | | |
|---|---|---|---|---|---|
| | **HR** | **SE** | **95% CI** | | ***p*** |
| **Nervous Disorders** | | | | | |
| Model 1: *Unadjusted* | 1.49 | 0.26 | 1.06 | 2.10 | 0.023 |
| Model 2: *Model 1 + demographics & genetics* [a] | 1.34 | 0.24 | 0.94 | 1.91 | 0.109 |
| Model 3: *Model 2 + Fully Adjusted* [b] | 1.08 | 0.20 | 0.74 | 1.56 | 0.703 |
| Model 2a: *Demographics* | 1.26 | 0.23 | 0.89 | 1.80 | 0.195 |
| Model 2b: *Genetics* | 1.52 | 0.27 | 1.08 | 2.15 | 0.018 |
| Model 3a: Model 3 + *Medication* | 1.08 | 0.21 | 0.74 | 1.56 | 0.692 |
| Model 3b: Model 3 + *Physical Activity* | 1.03 | 0.20 | 0.71 | 1.49 | 0.896 |
| Model 3c: Model 3 + *Body Mass Index* | 1.03 | 0.20 | 0.71 | 1.50 | 0.873 |
| Model 3d: Model 3 + *Fully Adjusted* [c] | 0.99 | 0.19 | 0.68 | 1.45 | 0.975 |
| Model 4a: Model 2 + *Health* | 1.30 | 0.24 | 0.91 | 1.87 | 0.147 |
| Model 4b: Model 3 + *Health* | 1.06 | 0.20 | 0.73 | 1.53 | 0.778 |
| Model 4c: Model 3 + *Fully Adjusted* [d] | 0.98 | 0.19 | 0.67 | 1.43 | 0.917 |

**Notes:** The *low-risk* group is the reference; HR = hazard ratio; SE = standard error; CI = confidence interval; *p* = significance value.

a *Demographic and genetic variables*: age; sex; 10 principal components (PCs); C-reactive Protein (CRP) polygenic score (PGS); White Blood Cell Counts (WBCC) PGS; Insulin Growth Factor-1 (IGF-1) PGS; Anxiety PGS; Depression PGS; Schizophrenia PGS; Insomnia PGS; Pain PGS.

b All variables: age; sex; 10 PCs; CRP PGS; WBCC PGS; IGF-1 PGS; PGS; Anxiety PGS; Depression PGS; Schizophrenia PGS; Insomnia PGS; Pain PGS; education; wealth; smoking status; alcohol consumption; mobility.

c Additional variables: medication, physical activity; BMI.

d Additional variables: health (i.e., chronic lung disease; coronary heart disease; abnormal heart rhythm; heart murmur; congestive heart failure; angina; hypertension; diabetes; cancer; Parkinson's; Alzheimer's; dementia; asthma; arthritis; osteoporosis; psychiatric disorder).

***Table S9ax.*** **Longitudinal associations between insulin growth factor-1 and hospitalisation for diseases of the nervous system (*n*=4,057)**

| Adjustments | IGF-1 | | | | |
|---|---|---|---|---|---|
| | HR | SE | 95% CI | | *p* |
| **Nervous Disorders** | | | | | |
| Model 1: *Unadjusted* | 0.64 | 0.08 | 0.49 | 0.82 | 0.001 |
| Model 2: *Model 1 + demographics & genetics* [a] | 0.84 | 0.11 | 0.64 | 1.09 | 0.185 |
| Model 3: *Model 2 + Fully Adjusted* [b] | 0.84 | 0.11 | 0.64 | 1.09 | 0.178 |
| Model 2a: *Demographics* | 0.84 | 0.11 | 0.65 | 1.10 | 0.201 |
| Model 2b: *Genetics* | 0.64 | 0.08 | 0.50 | 0.83 | 0.001 |
| Model 3a: Model 3 + *Medication* | 0.83 | 0.11 | 0.64 | 1.08 | 0.172 |
| Model 3b: Model 3 + *Physical Activity* | 0.85 | 0.11 | 0.65 | 1.10 | 0.205 |
| Model 3c: Model 3 + *Body Mass Index* | 0.84 | 0.11 | 0.65 | 1.09 | 0.188 |
| Model 3d: Model 3 + *Fully Adjusted* [c] | 0.85 | 0.11 | 0.65 | 1.10 | 0.208 |
| Model 4a: Model 2 + *Health* | 0.85 | 0.11 | 0.65 | 1.10 | 0.208 |
| Model 4b: Model 3 + *Health* | 0.84 | 0.11 | 0.65 | 1.09 | 0.194 |
| Model 4c: Model 3 + *Fully Adjusted* [d] | 0.85 | 0.11 | 0.66 | 1.10 | 0.224 |

**Notes:** The *low-risk* group is the reference; HR = hazard ratio; SE = standard error; CI = confidence interval; *p* = significance value.

a *Demographic and genetic variables*: age; sex; 10 principal components (PCs); C-reactive Protein (CRP) polygenic score (PGS); White Blood Cell Counts (WBCC) PGS; Insulin Growth Factor-1 (IGF-1) PGS; Anxiety PGS; Depression PGS; Schizophrenia PGS; Insomnia PGS; Pain PGS.

b All variables: age; sex; 10 PCs; CRP PGS; WBCC PGS; IGF-1 PGS; PGS; Anxiety PGS; Depression PGS; Schizophrenia PGS; Insomnia PGS; Pain PGS; education; wealth; smoking status; alcohol consumption; mobility.

c Additional variables: medication, physical activity; BMI.

d Additional variables: health (i.e., chronic lung disease; coronary heart disease; abnormal heart rhythm; heart murmur; congestive heart failure; angina; hypertension; diabetes; cancer; Parkinson's; Alzheimer's; dementia; asthma; arthritis; osteoporosis; psychiatric disorder).

***Table S9ay.*** **Longitudinal associations between immune and neuroendocrine profiles and hospitalisation for diseases of the respiratory system (*n*=3,846)**

| Adjustments | Immune and Neuroendocrine Profiles | | | | |
|---|---|---|---|---|---|
| | HR | SE | 95% CI | | *p* |
| ***Moderate-risk* Profile \| Respiratory Disorders** | | | | | |
| Model 1: *Unadjusted* | 1.59 | 0.12 | 1.38 | 1.84 | <0.001 |
| Model 2: *Model 1 + demographics & genetics* [a] | 1.41 | 0.11 | 1.22 | 1.63 | <0.001 |
| Model 3: *Model 2 + Fully Adjusted* [b] | 1.26 | 0.10 | 1.09 | 1.47 | 0.002 |
| Model 2a: *Demographics* | 1.40 | 0.10 | 1.21 | 1.62 | <0.001 |
| Model 2b: *Genetics* | 1.60 | 0.12 | 1.39 | 1.85 | <0.001 |
| Model 3a: Model 3 + *Medication* | 1.26 | 0.10 | 1.08 | 1.46 | 0.003 |
| Model 3b: Model 3 + *Physical Activity* | 1.25 | 0.10 | 1.08 | 1.45 | 0.004 |
| Model 3c: Model 3 + *Body Mass Index* | 1.29 | 0.10 | 1.11 | 1.51 | 0.001 |
| Model 3d: Model 3 + *Fully Adjusted* [c] | 1.28 | 0.10 | 1.09 | 1.49 | 0.002 |
| Model 4a: Model 2 + *Health* | 1.41 | 0.11 | 1.22 | 1.63 | <0.001 |
| Model 4b: Model 3 + *Health* | 1.25 | 0.10 | 1.08 | 1.45 | 0.003 |
| Model 4c: Model 3 + *Fully Adjusted* [d] | 1.26 | 0.10 | 1.08 | 1.47 | 0.003 |
| ***High-risk* Profile \| Respiratory Disorders** | | | | | |
| Model 1: *Unadjusted* | 2.64 | 0.25 | 2.20 | 3.18 | <0.001 |
| Model 2: *Model 1 + demographics & genetics* [a] | 2.34 | 0.22 | 1.94 | 2.82 | <0.001 |
| Model 3: *Model 2 + Fully Adjusted* [b] | 1.99 | 0.19 | 1.64 | 2.40 | <0.001 |
| Model 2a: *Demographics* | 2.34 | 0.22 | 1.95 | 2.82 | <0.001 |
| Model 2b: *Genetics* | 2.64 | 0.25 | 2.19 | 3.18 | <0.001 |
| Model 3a: Model 3 + *Medication* | 1.99 | 0.19 | 1.65 | 2.41 | <0.001 |
| Model 3b: Model 3 + *Physical Activity* | 1.94 | 0.19 | 1.61 | 2.35 | <0.001 |
| Model 3c: Model 3 + *Body Mass Index* | 2.04 | 0.20 | 1.68 | 2.48 | <0.001 |
| Model 3d: Model 3 + *Fully Adjusted* [c] | 2.01 | 0.20 | 1.65 | 2.44 | <0.001 |
| Model 4a: Model 2 + *Health* | 2.34 | 0.22 | 1.94 | 2.82 | <0.001 |
| Model 4b: Model 3 + *Health* | 1.98 | 0.19 | 1.64 | 2.40 | <0.001 |
| Model 4c: Model 3 + *Fully Adjusted* [d] | 2.01 | 0.20 | 1.66 | 2.44 | <0.001 |

**Notes:** The *low-risk* group is the reference; HR = hazard ratio; SE = standard error; CI = confidence interval; *p* = significance value.

a *Demographic and genetic variables*: age; sex; 10 principal components (PCs); C-reactive Protein (CRP) polygenic score (PGS); White Blood Cell Counts (WBCC) PGS; Insulin Growth Factor-1 (IGF-1) PGS; Anxiety PGS; Depression PGS; Schizophrenia PGS; Insomnia PGS; Pain PGS.

b All variables: age; sex; 10 PCs; CRP PGS; WBCC PGS; IGF-1 PGS; PGS; Anxiety PGS; Depression PGS; Schizophrenia PGS; Insomnia PGS; Pain PGS; education; wealth; smoking status; alcohol consumption; mobility.

c Additional variables: medication, physical activity; BMI.

d Additional variables: health (i.e., chronic lung disease; coronary heart disease; abnormal heart rhythm; heart murmur; congestive heart failure; angina; hypertension; diabetes; cancer; Parkinson's; Alzheimer's; dementia; asthma; arthritis; osteoporosis; psychiatric disorder).

***Table S9az.*** **Longitudinal associations between C-reactive protein and hospitalisation for diseases of the respiratory system (*n*=3,846)**

| Adjustments | CRP | | | | |
|---|---|---|---|---|---|
| | HR | SE | 95% CI | | *p* |
| **Respiratory Disorders** | | | | | |
| Model 1: *Unadjusted* | 1.67 | 0.08 | 1.52 | 1.83 | <0.001 |
| Model 2: *Model 1 + demographics & genetics* [a] | 1.56 | 0.08 | 1.42 | 1.72 | <0.001 |
| Model 3: *Model 2 + Fully Adjusted* [b] | 1.43 | 0.07 | 1.30 | 1.58 | <0.001 |
| Model 2a: *Demographics* | 1.56 | 0.08 | 1.42 | 1.71 | <0.001 |
| Model 2b: *Genetics* | 1.68 | 0.08 | 1.53 | 1.84 | <0.001 |
| Model 3a: Model 3 + *Medication* | 1.44 | 0.07 | 1.30 | 1.58 | <0.001 |
| Model 3b: Model 3 + *Physical Activity* | 1.42 | 0.07 | 1.29 | 1.56 | <0.001 |
| Model 3c: Model 3 + *Body Mass Index* | 1.47 | 0.08 | 1.33 | 1.62 | <0.001 |
| Model 3d: Model 3 + *Fully Adjusted* [c] | 1.45 | 0.08 | 1.31 | 1.61 | <0.001 |
| Model 4a: Model 2 + *Health* | 1.56 | 0.08 | 1.42 | 1.72 | <0.001 |
| Model 4b: Model 3 + *Health* | 1.43 | 0.07 | 1.30 | 1.58 | <0.001 |
| Model 4c: Model 3 + *Fully Adjusted* [d] | 1.45 | 0.08 | 1.31 | 1.61 | <0.001 |

**Notes:** The *low-risk* group is the reference; HR = hazard ratio; SE = standard error; CI = confidence interval; *p* = significance value.

a *Demographic and genetic variables*: age; sex; 10 principal components (PCs); C-reactive Protein (CRP) polygenic score (PGS); White Blood Cell Counts (WBCC) PGS; Insulin Growth Factor-1 (IGF-1) PGS; Anxiety PGS; Depression PGS; Schizophrenia PGS; Insomnia PGS; Pain PGS.

b All variables: age; sex; 10 PCs; CRP PGS; WBCC PGS; IGF-1 PGS; PGS; Anxiety PGS; Depression PGS; Schizophrenia PGS; Insomnia PGS; Pain PGS; education; wealth; smoking status; alcohol consumption; mobility.

c Additional variables: medication, physical activity; BMI.

d Additional variables: health (i.e., chronic lung disease; coronary heart disease; abnormal heart rhythm; heart murmur; congestive heart failure; angina; hypertension; diabetes; cancer; Parkinson's; Alzheimer's; dementia; asthma; arthritis; osteoporosis; psychiatric disorder).

***Table S9ba.*** **Longitudinal associations between fibrinogen and hospitalisation for diseases of the respiratory system (*n*=3,846)**

| Adjustments | Fb | | | | |
|---|---|---|---|---|---|
| | HR | SE | 95% CI | | *p* |
| **Respiratory Disorders** | | | | | |
| Model 1: *Unadjusted* | 1.64 | 0.10 | 1.47 | 1.84 | <0.001 |
| Model 2: *Model 1 + demographics & genetics* [a] | 1.48 | 0.09 | 1.31 | 1.66 | <0.001 |
| Model 3: *Model 2 + Fully Adjusted* [b] | 1.30 | 0.08 | 1.15 | 1.46 | <0.001 |
| Model 2a: *Demographics* | 1.52 | 0.09 | 1.35 | 1.71 | <0.001 |
| Model 2b: *Genetics* | 1.62 | 0.09 | 1.44 | 1.81 | <0.001 |
| Model 3a: Model 3 + *Medication* | 1.29 | 0.08 | 1.15 | 1.46 | <0.001 |
| Model 3b: Model 3 + *Physical Activity* | 1.29 | 0.08 | 1.14 | 1.45 | <0.001 |
| Model 3c: Model 3 + *Body Mass Index* | 1.30 | 0.08 | 1.15 | 1.47 | <0.001 |
| Model 3d: Model 3 + *Fully Adjusted* [c] | 1.29 | 0.08 | 1.15 | 1.46 | <0.001 |
| Model 4a: Model 2 + *Health* | 1.48 | 0.09 | 1.32 | 1.67 | <0.001 |
| Model 4b: Model 3 + *Health* | 1.31 | 0.08 | 1.16 | 1.48 | <0.001 |
| Model 4c: Model 3 + *Fully Adjusted* [d] | 1.31 | 0.08 | 1.16 | 1.48 | <0.001 |

**Notes:** The *low-risk* group is the reference; HR = hazard ratio; SE = standard error; CI = confidence interval; *p* = significance value.

a *Demographic and genetic variables*: age; sex; 10 principal components (PCs); C-reactive Protein (CRP) polygenic score (PGS); White Blood Cell Counts (WBCC) PGS; Insulin Growth Factor-1 (IGF-1) PGS; Anxiety PGS; Depression PGS; Schizophrenia PGS; Insomnia PGS; Pain PGS.

b All variables: age; sex; 10 PCs; CRP PGS; WBCC PGS; IGF-1 PGS; PGS; Anxiety PGS; Depression PGS; Schizophrenia PGS; Insomnia PGS; Pain PGS; education; wealth; smoking status; alcohol consumption; mobility.

c Additional variables: medication, physical activity; BMI.

d Additional variables: health (i.e., chronic lung disease; coronary heart disease; abnormal heart rhythm; heart murmur; congestive heart failure; angina; hypertension; diabetes; cancer; Parkinson's; Alzheimer's; dementia; asthma; arthritis; osteoporosis; psychiatric disorder).

***Table S9bb.*** **Longitudinal associations between white blood cell counts and hospitalisation for diseases of the respiratory system (*n*=3,846)**

| Adjustments | WBCC | | | | |
|---|---|---|---|---|---|
| | HR | SE | 95% CI | | *p* |
| **Respiratory Disorders** | | | | | |
| Model 1: *Unadjusted* | 3.18 | 0.42 | 2.45 | 4.13 | <0.001 |
| Model 2: *Model 1 + demographics & genetics* [a] | 3.02 | 0.41 | 2.32 | 3.94 | <0.001 |
| Model 3: *Model 2 + Fully Adjusted* [b] | 2.18 | 0.31 | 1.65 | 2.87 | <0.001 |
| Model 2a: *Demographics* | 2.82 | 0.38 | 2.17 | 3.67 | <0.001 |
| Model 2b: *Genetics* | 3.24 | 0.43 | 2.50 | 4.20 | <0.001 |
| Model 3a: Model 3 + *Medication* | 2.16 | 0.30 | 1.64 | 2.85 | <0.001 |
| Model 3b: Model 3 + *Physical Activity* | 2.13 | 0.30 | 1.62 | 2.80 | <0.001 |
| Model 3c: Model 3 + *Body Mass Index* | 2.19 | 0.31 | 1.66 | 2.89 | <0.001 |
| Model 3d: Model 3 + *Fully Adjusted* [c] | 2.14 | 0.30 | 1.62 | 2.81 | <0.001 |
| Model 4a: Model 2 + *Health* | 2.88 | 0.39 | 2.21 | 3.74 | <0.001 |
| Model 4b: Model 3 + *Health* | 2.10 | 0.29 | 1.60 | 2.76 | <0.001 |
| Model 4c: Model 3 + *Fully Adjusted* [d] | 2.07 | 0.29 | 1.58 | 2.72 | <0.001 |

**Notes:** The *low-risk* group is the reference; HR = hazard ratio; SE = standard error; CI = confidence interval; *p* = significance value.

a *Demographic and genetic variables*: age; sex; 10 principal components (PCs); C-reactive Protein (CRP) polygenic score (PGS); White Blood Cell Counts (WBCC) PGS; Insulin Growth Factor-1 (IGF-1) PGS; Anxiety PGS; Depression PGS; Schizophrenia PGS; Insomnia PGS; Pain PGS.

b All variables: age; sex; 10 PCs; CRP PGS; WBCC PGS; IGF-1 PGS; PGS; Anxiety PGS; Depression PGS; Schizophrenia PGS; Insomnia PGS; Pain PGS; education; wealth; smoking status; alcohol consumption; mobility.

c Additional variables: medication, physical activity; BMI.

d Additional variables: health (i.e., chronic lung disease; coronary heart disease; abnormal heart rhythm; heart murmur; congestive heart failure; angina; hypertension; diabetes; cancer; Parkinson's; Alzheimer's; dementia; asthma; arthritis; osteoporosis; psychiatric disorder).

***Table S9bc.*** **Longitudinal associations between insulin growth factor-1 and hospitalisation for diseases of the respiratory system (*n*=3,846)**

| **Adjustments** | **IGF-1** | | | | |
|---|---|---|---|---|---|
| | **HR** | **SE** | **95% CI** | | ***p*** |
| **Respiratory Disorders** | | | | | |
| Model 1: *Unadjusted* | 0.53 | 0.05 | 0.43 | 0.64 | <0.001 |
| Model 2: *Model 1 + demographics & genetics* [a] | 0.73 | 0.08 | 0.60 | 0.89 | 0.002 |
| Model 3: *Model 2 + Fully Adjusted* [b] | 0.73 | 0.08 | 0.60 | 0.90 | 0.002 |
| Model 2a: *Demographics* | 0.75 | 0.08 | 0.62 | 0.92 | 0.006 |
| Model 2b: *Genetics* | 0.52 | 0.05 | 0.43 | 0.63 | <0.001 |
| Model 3a: Model 3 + *Medication* | 0.73 | 0.08 | 0.60 | 0.90 | 0.002 |
| Model 3b: Model 3 + *Physical Activity* | 0.74 | 0.08 | 0.61 | 0.90 | 0.003 |
| Model 3c: Model 3 + *Body Mass Index* | 0.73 | 0.08 | 0.60 | 0.90 | 0.002 |
| Model 3d: Model 3 + *Fully Adjusted* [c] | 0.74 | 0.08 | 0.61 | 0.91 | 0.003 |
| Model 4a: Model 2 + *Health* | 0.74 | 0.08 | 0.60 | 0.90 | 0.003 |
| Model 4b: Model 3 + *Health* | 0.74 | 0.08 | 0.60 | 0.90 | 0.003 |
| Model 4c: Model 3 + *Fully Adjusted* [d] | 0.75 | 0.08 | 0.61 | 0.91 | 0.004 |

**Notes:** The *low-risk* group is the reference; HR = hazard ratio; SE = standard error; CI = confidence interval; *p* = significance value.

a *Demographic and genetic variables*: age; sex; 10 principal components (PCs); C-reactive Protein (CRP) polygenic score (PGS); White Blood Cell Counts (WBCC) PGS; Insulin Growth Factor-1 (IGF-1) PGS; Anxiety PGS; Depression PGS; Schizophrenia PGS; Insomnia PGS; Pain PGS.

b All variables: age; sex; 10 PCs; CRP PGS; WBCC PGS; IGF-1 PGS; PGS; Anxiety PGS; Depression PGS; Schizophrenia PGS; Insomnia PGS; Pain PGS; education; wealth; smoking status; alcohol consumption; mobility.

c Additional variables: medication, physical activity; BMI.

d Additional variables: health (i.e., chronic lung disease; coronary heart disease; abnormal heart rhythm; heart murmur; congestive heart failure; angina; hypertension; diabetes; cancer; Parkinson's; Alzheimer's; dementia; asthma; arthritis; osteoporosis; psychiatric disorder).

***Table S9bd.*** **Longitudinal associations between immune and neuroendocrine profiles and hospitalisation for diseases of the skin and subcutaneous tissue (*n*=4,045)**

| **Adjustments** | **Immune and Neuroendocrine Profiles** | | | | |
|---|---|---|---|---|---|
| | **HR** | **SE** | **95% CI** | | ***p*** |
| ***Moderate-risk* Profile \| Skin Disorders** | | | | | |
| Model 1: *Unadjusted* | 1.22 | 0.12 | 1.00 | 1.48 | 0.048 |
| Model 2: *Model 1 + demographics & genetics* [a] | 1.07 | 0.11 | 0.88 | 1.31 | 0.491 |
| Model 3: *Model 2 + Fully Adjusted* [b] | 1.00 | 0.10 | 0.82 | 1.23 | 0.968 |
| Model 2a: *Demographics* | 1.08 | 0.11 | 0.89 | 1.31 | 0.454 |
| Model 2b: *Genetics* | 1.22 | 0.12 | 1.01 | 1.49 | 0.044 |
| Model 3a: Model 3 + *Medication* | 1.01 | 0.10 | 0.82 | 1.23 | 0.965 |
| Model 3b: Model 3 + *Physical Activity* | 0.99 | 0.10 | 0.81 | 1.22 | 0.946 |
| Model 3c: Model 3 + *Body Mass Index* | 0.97 | 0.10 | 0.79 | 1.19 | 0.773 |
| Model 3d: Model 3 + *Fully Adjusted* [c] | 0.96 | 0.10 | 0.78 | 1.19 | 0.721 |
| Model 4a: Model 2 + *Health* | 1.07 | 0.11 | 0.88 | 1.31 | 0.491 |
| Model 4b: Model 3 + *Health* | 1.00 | 0.10 | 0.82 | 1.23 | 0.988 |
| Model 4c: Model 3 + *Fully Adjusted* [d] | 0.96 | 0.10 | 0.78 | 1.18 | 0.710 |
| ***High-risk* Profile \| Skin Disorders** | | | | | |
| Model 1: *Unadjusted* | 1.49 | 0.21 | 1.13 | 1.96 | 0.005 |
| Model 2: *Model 1 + demographics & genetics* [a] | 1.33 | 0.19 | 1.00 | 1.76 | 0.047 |
| Model 3: *Model 2 + Fully Adjusted* [b] | 1.19 | 0.17 | 0.90 | 1.58 | 0.231 |
| Model 2a: *Demographics* | 1.33 | 0.19 | 1.01 | 1.76 | 0.043 |
| Model 2b: *Genetics* | 1.49 | 0.21 | 1.13 | 1.97 | 0.005 |
| Model 3a: Model 3 + *Medication* | 1.19 | 0.17 | 0.89 | 1.58 | 0.237 |
| Model 3b: Model 3 + *Physical Activity* | 1.17 | 0.17 | 0.88 | 1.55 | 0.292 |
| Model 3c: Model 3 + *Body Mass Index* | 1.15 | 0.17 | 0.86 | 1.53 | 0.361 |
| Model 3d: Model 3 + *Fully Adjusted* [c] | 1.12 | 0.17 | 0.84 | 1.50 | 0.434 |
| Model 4a: Model 2 + *Health* | 1.33 | 0.19 | 1.00 | 1.76 | 0.047 |
| Model 4b: Model 3 + *Health* | 1.19 | 0.17 | 0.89 | 1.58 | 0.236 |
| Model 4c: Model 3 + *Fully Adjusted* [d] | 1.12 | 0.17 | 0.84 | 1.50 | 0.437 |

**Notes:** The *low-risk* group is the reference; HR = hazard ratio; SE = standard error; CI = confidence interval; *p* = significance value.

a *Demographic and genetic variables*: age; sex; 10 principal components (PCs); C-reactive Protein (CRP) polygenic score (PGS); White Blood Cell Counts (WBCC) PGS; Insulin Growth Factor-1 (IGF-1) PGS; Anxiety PGS; Depression PGS; Schizophrenia PGS; Insomnia PGS; Pain PGS.

b All variables: age; sex; 10 PCs; CRP PGS; WBCC PGS; IGF-1 PGS; PGS; Anxiety PGS; Depression PGS; Schizophrenia PGS; Insomnia PGS; Pain PGS; education; wealth; smoking status; alcohol consumption; mobility.

c Additional variables: medication, physical activity; BMI.

d Additional variables: health (i.e., chronic lung disease; coronary heart disease; abnormal heart rhythm; heart murmur; congestive heart failure; angina; hypertension; diabetes; cancer; Parkinson's; Alzheimer's; dementia; asthma; arthritis; osteoporosis; psychiatric disorder).

***Table S9be.*** **Longitudinal associations between C-reactive protein and hospitalisation for diseases of the skin and subcutaneous tissue (*n*=4,045)**

| **Adjustments** | **CRP** | | | | |
|---|---|---|---|---|---|
| | **HR** | **SE** | **95% CI** | | ***p*** |
| **Skin Disorders** | | | | | |
| Model 1: *Unadjusted* | 1.22 | 0.08 | 1.07 | 1.39 | 0.004 |
| Model 2: *Model 1 + demographics & genetics* [a] | 1.13 | 0.08 | 0.99 | 1.30 | 0.076 |
| Model 3: *Model 2 + Fully Adjusted* [b] | 1.07 | 0.08 | 0.93 | 1.23 | 0.355 |
| Model 2a: *Demographics* | 1.13 | 0.08 | 0.99 | 1.30 | 0.076 |
| Model 2b: *Genetics* | 1.22 | 0.08 | 1.07 | 1.40 | 0.003 |
| Model 3a: Model 3 + *Medication* | 1.07 | 0.08 | 0.93 | 1.23 | 0.361 |
| Model 3b: Model 3 + *Physical Activity* | 1.06 | 0.08 | 0.92 | 1.22 | 0.453 |
| Model 3c: Model 3 + *Body Mass Index* | 1.04 | 0.08 | 0.90 | 1.21 | 0.591 |
| Model 3d: Model 3 + *Fully Adjusted* [c] | 1.03 | 0.08 | 0.89 | 1.19 | 0.694 |
| Model 4a: Model 2 + *Health* | 1.13 | 0.08 | 0.99 | 1.30 | 0.076 |
| Model 4b: Model 3 + *Health* | 1.07 | 0.08 | 0.93 | 1.23 | 0.369 |
| Model 4c: Model 3 + *Fully Adjusted* [d] | 1.03 | 0.08 | 0.89 | 1.19 | 0.704 |

**Notes:** The *low-risk* group is the reference; HR = hazard ratio; SE = standard error; CI = confidence interval; *p* = significance value.

a *Demographic and genetic variables*: age; sex; 10 principal components (PCs); C-reactive Protein (CRP) polygenic score (PGS); White Blood Cell Counts (WBCC) PGS; Insulin Growth Factor-1 (IGF-1) PGS; Anxiety PGS; Depression PGS; Schizophrenia PGS; Insomnia PGS; Pain PGS.

b All variables: age; sex; 10 PCs; CRP PGS; WBCC PGS; IGF-1 PGS; PGS; Anxiety PGS; Depression PGS; Schizophrenia PGS; Insomnia PGS; Pain PGS; education; wealth; smoking status; alcohol consumption; mobility.

c Additional variables: medication, physical activity; BMI.

d Additional variables: health (i.e., chronic lung disease; coronary heart disease; abnormal heart rhythm; heart murmur; congestive heart failure; angina; hypertension; diabetes; cancer; Parkinson's; Alzheimer's; dementia; asthma; arthritis; osteoporosis; psychiatric disorder).

***Table S9bf.*** **Longitudinal associations between fibrinogen and hospitalisation for diseases of the skin and subcutaneous tissue (*n*=4,045)**

| **Adjustments** | **Fb** | | | | |
|---|---|---|---|---|---|
| | **HR** | **SE** | **95% CI** | | ***p*** |
| **Skin Disorders** | | | | | |
| Model 1: *Unadjusted* | 1.17 | 0.10 | 1.00 | 1.38 | 0.053 |
| Model 2: *Model 1 + demographics & genetics* [a] | 1.07 | 0.09 | 0.91 | 1.27 | 0.418 |
| Model 3: *Model 2 + Fully Adjusted* [b] | 0.99 | 0.09 | 0.83 | 1.17 | 0.883 |
| Model 2a: *Demographics* | 1.07 | 0.09 | 0.91 | 1.27 | 0.404 |
| Model 2b: *Genetics* | 1.17 | 0.10 | 1.00 | 1.38 | 0.054 |
| Model 3a: Model 3 + *Medication* | 0.99 | 0.09 | 0.83 | 1.17 | 0.888 |
| Model 3b: Model 3 + *Physical Activity* | 0.98 | 0.09 | 0.83 | 1.17 | 0.849 |
| Model 3c: Model 3 + *Body Mass Index* | 0.97 | 0.09 | 0.81 | 1.15 | 0.720 |
| Model 3d: Model 3 + *Fully Adjusted* [c] | 0.97 | 0.09 | 0.82 | 1.15 | 0.716 |
| Model 4a: Model 2 + *Health* | 1.07 | 0.09 | 0.90 | 1.27 | 0.432 |
| Model 4b: Model 3 + *Health* | 0.99 | 0.09 | 0.83 | 1.17 | 0.884 |
| Model 4c: Model 3 + *Fully Adjusted* [d] | 0.97 | 0.09 | 0.82 | 1.15 | 0.718 |

**Notes:** The *low-risk* group is the reference; HR = hazard ratio; SE = standard error; CI = confidence interval; *p* = significance value.

a *Demographic and genetic variables*: age; sex; 10 principal components (PCs); C-reactive Protein (CRP) polygenic score (PGS); White Blood Cell Counts (WBCC) PGS; Insulin Growth Factor-1 (IGF-1) PGS; Anxiety PGS; Depression PGS; Schizophrenia PGS; Insomnia PGS; Pain PGS.

b All variables: age; sex; 10 PCs; CRP PGS; WBCC PGS; IGF-1 PGS; PGS; Anxiety PGS; Depression PGS; Schizophrenia PGS; Insomnia PGS; Pain PGS; education; wealth; smoking status; alcohol consumption; mobility.

c Additional variables: medication, physical activity; BMI.

d Additional variables: health (i.e., chronic lung disease; coronary heart disease; abnormal heart rhythm; heart murmur; congestive heart failure; angina; hypertension; diabetes; cancer; Parkinson's; Alzheimer's; dementia; asthma; arthritis; osteoporosis; psychiatric disorder).

***Table S9bg.*** **Longitudinal associations between white blood cell counts and hospitalisation for diseases of the skin and subcutaneous tissue (*n*=4,045)**

| **Adjustments** | **WBCC** | | | | |
|---|---|---|---|---|---|
| | **HR** | **SE** | **95% CI** | | ***p*** |
| **Skin Disorders** | | | | | |
| Model 1: *Unadjusted* | 1.71 | 0.32 | 1.19 | 2.46 | 0.004 |
| Model 2: *Model 1 + demographics & genetics* [a] | 1.55 | 0.30 | 1.06 | 2.27 | 0.024 |
| Model 3: *Model 2 + Fully Adjusted* [b] | 1.25 | 0.25 | 0.84 | 1.85 | 0.275 |
| Model 2a: *Demographics* | 1.51 | 0.29 | 1.04 | 2.20 | 0.031 |
| Model 2b: *Genetics* | 1.75 | 0.33 | 1.21 | 2.52 | 0.003 |
| Model 3a: Model 3 + *Medication* | 1.24 | 0.25 | 0.84 | 1.85 | 0.279 |
| Model 3b: Model 3 + *Physical Activity* | 1.23 | 0.25 | 0.83 | 1.82 | 0.309 |
| Model 3c: Model 3 + *Body Mass Index* | 1.20 | 0.25 | 0.81 | 1.79 | 0.363 |
| Model 3d: Model 3 + *Fully Adjusted* [c] | 1.19 | 0.24 | 0.80 | 1.77 | 0.399 |
| Model 4a: Model 2 + *Health* | 1.53 | 0.30 | 1.05 | 2.24 | 0.028 |
| Model 4b: Model 3 + *Health* | 1.24 | 0.25 | 0.84 | 1.84 | 0.285 |
| Model 4c: Model 3 + *Fully Adjusted* [d] | 1.18 | 0.24 | 0.80 | 1.77 | 0.406 |

**Notes:** The *low-risk* group is the reference; HR = hazard ratio; SE = standard error; CI = confidence interval; *p* = significance value.

a *Demographic and genetic variables*: age; sex; 10 principal components (PCs); C-reactive Protein (CRP) polygenic score (PGS); White Blood Cell Counts (WBCC) PGS; Insulin Growth Factor-1 (IGF-1) PGS; Anxiety PGS; Depression PGS; Schizophrenia PGS; Insomnia PGS; Pain PGS.

b All variables: age; sex; 10 PCs; CRP PGS; WBCC PGS; IGF-1 PGS; PGS; Anxiety PGS; Depression PGS; Schizophrenia PGS; Insomnia PGS; Pain PGS; education; wealth; smoking status; alcohol consumption; mobility.

c Additional variables: medication, physical activity; BMI.

d Additional variables: health (i.e., chronic lung disease; coronary heart disease; abnormal heart rhythm; heart murmur; congestive heart failure; angina; hypertension; diabetes; cancer; Parkinson's; Alzheimer's; dementia; asthma; arthritis; osteoporosis; psychiatric disorder).

***Table S9bh.*** **Longitudinal associations between insulin growth factor-1 and hospitalisation for diseases of the skin and subcutaneous tissue (*n*=4,045)**

| **Adjustments** | **IGF-1** | | | | |
|---|---|---|---|---|---|
| | **HR** | **SE** | **95% CI** | | ***p*** |
| **Skin Disorders** | | | | | |
| Model 1: *Unadjusted* | 0.71 | 0.10 | 0.54 | 0.93 | 0.012 |
| Model 2: *Model 1 + demographics & genetics* [a] | 0.94 | 0.14 | 0.71 | 1.24 | 0.665 |
| Model 3: *Model 2 + Fully Adjusted* [b] | 0.95 | 0.13 | 0.72 | 1.25 | 0.711 |
| Model 2a: *Demographics* | 0.95 | 0.14 | 0.72 | 1.25 | 0.705 |
| Model 2b: *Genetics* | 0.70 | 0.10 | 0.53 | 0.91 | 0.009 |
| Model 3a: Model 3 + *Medication* | 0.95 | 0.13 | 0.72 | 1.25 | 0.718 |
| Model 3b: Model 3 + *Physical Activity* | 0.95 | 0.13 | 0.72 | 1.26 | 0.736 |
| Model 3c: Model 3 + *Body Mass Index* | 0.96 | 0.13 | 0.73 | 1.26 | 0.761 |
| Model 3d: Model 3 + *Fully Adjusted* [c] | 0.96 | 0.13 | 0.73 | 1.26 | 0.761 |
| Model 4a: Model 2 + *Health* | 0.94 | 0.13 | 0.71 | 1.24 | 0.664 |
| Model 4b: Model 3 + *Health* | 0.95 | 0.13 | 0.72 | 1.25 | 0.710 |
| Model 4c: Model 3 + *Fully Adjusted* [d] | 0.96 | 0.13 | 0.73 | 1.26 | 0.761 |

**Notes:** The *low-risk* group is the reference; HR = hazard ratio; SE = standard error; CI = confidence interval; *p* = significance value.

a *Demographic and genetic variables*: age; sex; 10 principal components (PCs); C-reactive Protein (CRP) polygenic score (PGS); White Blood Cell Counts (WBCC) PGS; Insulin Growth Factor-1 (IGF-1) PGS; Anxiety PGS; Depression PGS; Schizophrenia PGS; Insomnia PGS; Pain PGS.

b All variables: age; sex; 10 PCs; CRP PGS; WBCC PGS; IGF-1 PGS; PGS; Anxiety PGS; Depression PGS; Schizophrenia PGS; Insomnia PGS; Pain PGS; education; wealth; smoking status; alcohol consumption; mobility.

c Additional variables: medication, physical activity; BMI.

d Additional variables: health (i.e., chronic lung disease; coronary heart disease; abnormal heart rhythm; heart murmur; congestive heart failure; angina; hypertension; diabetes; cancer; Parkinson's; Alzheimer's; dementia; asthma; arthritis; osteoporosis; psychiatric disorder).
